# Home-based transcranial direct current stimulation RCT in major depression

**DOI:** 10.1101/2023.11.27.23299059

**Authors:** Rachel D. Woodham, Sudhakar Selvaraj, Nahed Lajmi, Harriet Hobday, Gabrielle Sheehan, Ali-Reza Ghazi-Noori, Peter J. Lagerberg, Maheen Rizvi, Sarah S. Kwon, Paulette Orhii, David Maislin, Lucia Hernandez, Rodrigo Machado-Vieira, Jair C. Soares, Allan H. Young, Cynthia H.Y. Fu

**Author notes:** Author for correspondence: Professor CHY Fu, University of East London, School of Psychology, Arthur Edwards Building, Water Lane, London E15 4LZ, UK.

## Abstract

**Background:** Transcranial direct current stimulation (tDCS) has been proposed as a novel treatment in major depressive disorder (MDD). However, efficacy and safety of home-based tDCS treatment have not been investigated.

**Methods:** Fully remote, multisite, double-blind, placebo-controlled, randomized superiority trial of home-based tDCS treatment in MDD was conducted in UK and USA. Participants were adults 18 years or older, having MDD diagnosis based on DSM-5 criteria, in current depressive episode of at least moderate severity, measured by score >=16 on 17-item Hamilton Depression Rating Scale (HDRS), without treatment resistant depression history. Protocol was 10-week blinded phase: 5 tDCS sessions per week for 3 weeks then 3 sessions per week for 7 weeks; followed by 10-week open label phase. tDCS montage was bifrontal, 30-minute sessions, active tDCS 2 mA, and sham tDCS 0 mA with brief ramp up and down to mimic active device. Primary outcome was HDRS change at week 10 in modified intention-to-treat analysis.

**Results:** 174 MDD participants were randomized: active (n=87; mean age 37.1 ± 11.1 years) and sham (n=87; mean age 38.3 ± 10.9 years) treatment. Significant improvement in HDRS was observed in active (9.4 ± 6.25 points) relative to sham treatment (7.1 ± 6.10 points) (95% CI 0.5 to 4.0, *p* = 0.012), with no differences in discontinuation rates between active (n=13) and sham (n=12).

**Conclusions:** Home-based tDCS with remote supervision is a potential first line treatment for MDD that is acceptable and safe. Consideration of continuing effective safety monitoring is required.

Trial registration number NCT05202119

## Introduction

Major depressive disorder (MDD) is common, is the leading cause of disability worldwide, and the most significant precursor in suicide.^1^ First line treatments are antidepressant medications and psychotherapy. However, a lack of remission is observed in over a third of MDD individuals to antidepressant medication as well as psychotherapy despite full treatment trials.^2,3^

Transcranial direct current stimulation (tDCS) is a form of non-invasive brain stimulation that applies a weak (0.5-2 mA) direct current via scalp electrodes.^4^ tDCS modulates cortical tissue excitability but does not directly trigger action potential in neuronal cells in contrast to repetitive transcranial magnetic stimulation (rTMS).^5^ tDCS is applied through a flexible cap or band worn over the forehead. The anode electrode is typically placed over left dorsolateral prefrontal cortex (DLPFC) and cathode over right DLPFC, suborbital or frontotemporal region.^5^ An individual patient data meta-analysis reported significantly greater clinical response (30.9% vs. 18.9%; number needed to treat (NNT) 9) and remission (19.9% vs. 11.7%; NNT 13) for active relative to sham tDCS. ^6^ tDCS is safe and well tolerated with no significant differences in attrition and adverse events between active and sham stimulation groups.^4^ However, a course of tDCS treatment requires daily sessions for several weeks.^5,6^ As it is portable and safe, tDCS could be provided at home,^4^ and open-label trials indicate high acceptability and feasibility.^7–9^

Our home-based tDCS treatment protocol with real-time remote supervision by video conference has demonstrated high feasibility, acceptability and safety.^9^ In the present trial, MDD participants were randomly allocated to either active or sham tDCS. The primary objective was efficacy of 10-week course of home-based, self-administered tDCS.

## Methods

### Trial design

All participants provided written informed consent. Ethical approval was provided by South Central-Hampshire B Research Ethics Committee, UK, and WIRB-Copernicus Group International Review Board, USA. Multisite, double-blind, placebo-controlled, randomized, superiority trial of home-based tDCS in MDD (ClinicalTrials.gov NCT05202119) was conducted in England and Wales, UK, and Texas, USA, at University of East London and University of Texas Health Science Center at Houston, respectively.

Recruitment was from May 12, 2022 to March 10, 2023. Potential participants were recruited through Flow Neuroscience website, email lists and online marketing. Participants were directed to online pre-screening form hosted by contract research organization (CRO), followed by pre-screening CRO telephone call, and then screening interview with site researchers. All assessments were conducted by Microsoft Teams video conference. Final open-label follow up was conducted on August 23, 2023.

The trial consisted of a 10-week blinded treatment phase followed by 10-week open label phase. The blinded phase consisted of random assignment to sham or active tDCS treatment in 1:1 ratio, performed independently at each site. Block randomization was used with permuted block sizes of 4 and 6, conducted by the trial server and stored in dedicated database. The Sponsor, Flow Neuroscience, provided tDCS devices (Flow FL-100). The Sponsor had no role in data analysis, interpretation of data, decision to publish, or manuscript preparation.

### Participants

Participants were adults >=18 years, with MDD in current depressive episode based on Diagnostic and Statistical Manual of Mental Disorders, Fifth Edition (DSM-5) criteria^10^ by structured assessment, Mini-International Neuropsychiatric Interview (MINI; Version 7.0.2).^11^ Inclusion criteria included: at least a moderate severity of depressive symptoms, as measured by score >=16 on 17-item Hamilton Depression Rating Scale (HDRS);^12^ being treatment free, or taking stable antidepressant medication, or in psychotherapy, for at least 6 weeks prior to enrolment, and agreeable to maintaining same treatment throughout the trial; under care of GP or psychiatrist. Exclusion criteria included: treatment resistant depression, defined as inadequate clinical response to two or more trials of antidepressant medication at an adequate dose and duration; significant suicide risk based on Columbia Suicide Severity Rating Scale (C-SSRS) Triage and Risk Identification Screener;^13^ comorbid psychiatric disorder; and taking medications that affect cortical excitability (e.g., benzodiazepines, epileptics). Full criteria are presented in Supplementary Appendix.

### Interventions

tDCS device is a headset placed over the forehead with two pre-positioned conductive rubber electrodes, each 23cm^2^. Anode is positioned over F3 and cathode over F4 on international 10/20 EEG system. Active tDCS stimulation is 2 mA direct current stimulation for 30 minutes with gradual ramp up over 120 seconds at the start and ramp down over 15 seconds at the end of each session. Sham tDCS stimulation is an initial ramp up from 0 to 1 mA over 30 seconds then ramp down to 0 mA over 15 seconds and repeated at session end to provide a tingling sensation which mimics active stimulation.

Blinded phase consisted of 5 tDCS sessions per week for 3 weeks followed by 3 tDCS sessions per week for 7 weeks. At week 10, participants and researchers were informed of treatment arm allocation. Open label phase consisted of active tDCS sessions for all participants. Participants in the initial active tDCS treatment arm were offered 3 sessions per week for 10 weeks, and participants in initial sham tDCS treatment arm were offered the active tDCS stimulation schedule, 5 sessions per week for 3 weeks then 3 sessions per week for 7 weeks.

tDCS stimulation was provided in study device app, and researchers had access to remote monitoring with real-time data use. Initial stimulation was supervised by video conference. Participants were asked to have video and microphone on during the session, were advised to sit or to lie down, and were able to engage in other tasks.

### Blinding

Participants and researchers were unaware of trial-group assignments. We sought to have the same researcher present for same MDD participant for trial duration. A second researcher joined clinical reviews for independent ratings. Adequacy of blinding was assessed by asking about allocation (active or sham) and certainty from 1 (very uncertain) to 5 (very certain).

### Clinical and Quality of Life Assessments

Assessments were performed by trained researchers and reviewed by consultant psychiatrists. Self-report measures were completed by participants in study app or online. Source records, electronic case report forms and data checking promoted outcome data quality. Assessments for depressive severity, suicidal ideation and manic symptoms were conducted at baseline and weeks 1, 4, 7, 10 and 20. Depressive severity was measured by clinician-rated scales, 17-item Hamilton Depression Rating Scale (HDRS),^12^ Montgomery-Åsberg Depression Rating Scale (MADRS),^14^ and self-report scale, Montgomery-Åsberg Depression Rating Scale - self-report (MADRS-s);^15^ suicide ideation, C-SSRS;^13^ mania symptoms, Young Mania Rating Scale (YMRS).^16^ At baseline, weeks 10 and 20, anxiety symptoms and quality of life were assessed by Hamilton Anxiety Rating Scale (HAMA),^17^ and EQ-5D-3L,^18–20^ which has five dimensions: mobility, self-care, usual activities, pain and discomfort, and anxiety and depression; with three severity levels.

### Primary and Secondary Outcomes in Clinical and Quality of Life Measures

Primary efficacy outcome was estimated mean group difference in depressive severity as measured by HDRS at week 10 compared to baseline in active and sham treatment arms. Secondary outcomes were all at week 10: clinical response, >=50% reduction from baseline in HDRS, MADRS and MADRS-s; clinical remission, HDRS score <=7, MADRS score <=10, and MADRS-s score <=12; clinician-rated depressive severity, MADRS; self-report, MADRS-s; and quality of life, EQ-5D-3L.

Exploratory outcomes included correlation between adherence to stimulation and HDRS, MADRS decrease in active treatment arm at week 10; changes in anxiety symptoms from baseline to week 10; and presence of hypomanic/manic symptoms at week 10.

Neuropsychological functioning was assessed by Rey Auditory Verbal Learning Test (RAVLT)^21^ for memory and verbal learning and Symbol Digit Modalities Test (SDMT)^22^ for psychomotor speed and visuospatial attention at baseline, weeks 10 and 20. Order and versions were counterbalanced. Assessments were mailed to participants, completed by pen and paper during session, and recorded by screenshot.

Treatment acceptability was assessed by our treatment acceptability questionnaire (TAQ)^9^ at baseline, weeks 10 and 20. Full description of exploratory outcomes is presented in Supplementary Appendix.

### Adverse events

Adverse events were assessed at each visit, and participants were able to contact the research team by a dedicated cell number at any time. tDCS Adverse Events Questionnaire (AEQ)^23^ was administered at weeks 10 and 20.

### Statistical Analysis

Sample size calculation was based on Brunoni et al,^24^ with two-sample t-test for mean difference with 80% power and one-sided Type 1 error 0.025, resulting in a sample size of 176 MDD participants. To increase power to 87.6%, sample size was increased to 216. Assuming 20% attrition rate, total sample size was 270 participants. Interim analysis was performed when 90 MDD participants completed week 10, which included both futility assessment and sample size re-estimation.^25^

Intent to treat (ITT) analysis consisted of all randomized participants and classified according to intended treatment. Participants excluded prior to randomization were considered screen failures. Modified intent-to-treat (mITT) analysis set included ITT participants who received at least 1 tDCS session (active or sham) and excluded participants randomized in error.

Primary effectiveness outcome was estimated mean group difference in HDRS scores in participants randomized to active and sham treatments using a mixed model for repeated measures (MMRM). The model included the baseline value for HDRS-17, usage of antidepressant use, psychotherapy treatment, age, and sex. Missing data was categorized by the reason for missingness (missing at random or not) and differentially imputed based on that classification. If *p*-value were less than one-sided *p* = 0.025, then endpoint would be declared positive. Secondary outcomes were: HDRS clinical response and remission, EQ-5D-3L change, and change in ratings, response and remission in MADRS and MADRS-s.

Standard deviations are provided based Cochran’s^26^ conversion of SE to SD weighted by sample size. Type 1 error was controlled by only testing the 3 named secondary endpoints after meeting the primary endpoint; nominal *p*-values are provided for all other evaluations.

## Results

### Patients

Based on blinded interim analysis, recruitment was ended early. 174 MDD participants were enrolled, randomised to active (n=87) and sham (n=87) treatment. 153 participants completed the protocol-specified number of sessions. One participant who was randomised did not continue and did not receive any treatment, therefore mITT sample was 173 participants. There were no significant differences in withdrawal rates between groups.

### Primary Outcome

In the primary hypothesis, significant improvement was observed in change in HDRS depressive severity from baseline to week 10 in active tDCS treatment arm, HDRS decrease 9.4 + 6.25 points (estimated week 10 HDRS, mean 9.6 + 6.02), as compared to sham tDCS treatment arm, HDRS decrease 7.1 + 6.10 points (estimated week 10 HDRS, mean 11.7 + 5.96) (95% CI 0.5 to 4.0, *p* = 0.012) (Figure 1).

**Figure 1.**
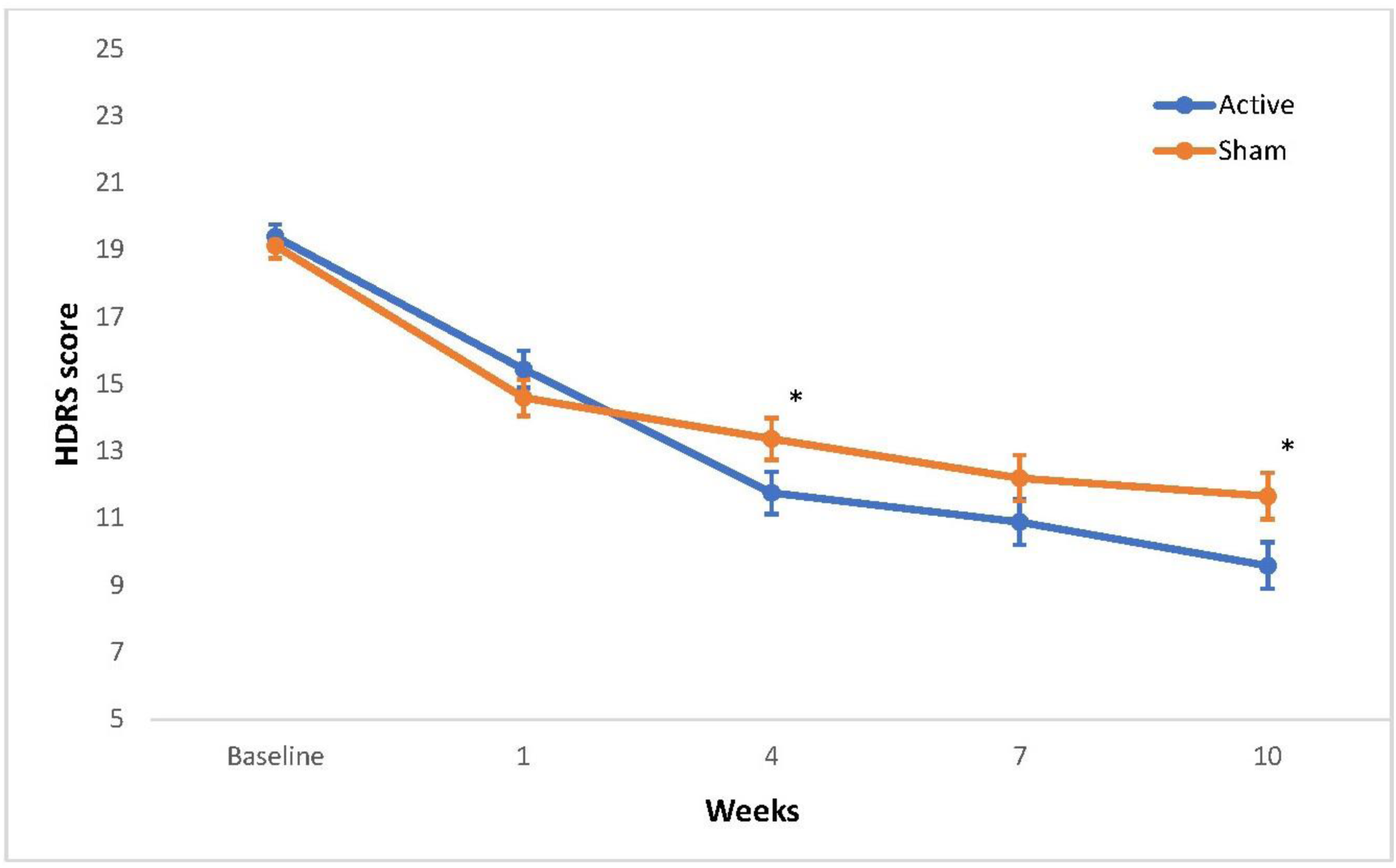
Change in depressive severity ratings over time. Shown are the estimated mean 17-item Hamilton Rating Scale for Depression (HDRS) rating scores from baseline to week 10 in the modified intention-to-treat analysis sample (n=173) for the active tDCS and sham tDCS treatment arms. Error bars represent ± 1 standard error (SE). HDRS scores range from 0 to 52 with higher values indicating more severe depressive symptoms. A significant improvement was observed in the change in HDRS ratings from baseline to week 10 in the active tDCS treatment arm, HDRS decrease 9.4 ± 6.25 (SD) (mean week 10 HDRS 9.6 ± 0.7 (SE)), as compared to sham tDCS treatment arm, HDRS decrease 7.1 ± 6.10 (SD) (mean week 10 HDRS 11.7 ± 0.7 (SE)) (95% CI 0.5 to 4.0, *p* = 0.012). The difference in change scores was also significant at week 4 (*p* = 0.03) with a greater score decrease in the active treatment arm. * = *p* <0.05.

### Secondary Outcomes

Based on HDRS ratings, active tDCS treatment arm showed a significantly greater clinical response of 54.4% relative to sham response of 26.9% (*p* = 0.001) (Post hoc Odds Ratio (OR) 3.25 (lower bound (LB) 1.57, upper bound (UB) 6.74), and the active treatment arm showed significantly greater remission rate 44.9% relative to sham arm 21.8% (*p* = 0.004) (Post hoc OR 2.93 (LB 1.41, UB 6.09).

Based on MADRS ratings, active tDCS treatment arm showed a significant improvement from baseline to week 10, mean improvement 11.3 ± 8.81 relative to sham treatment 7.7 ± 8.47 (*p* = 0.006). In clinical response, active treatment arm showed a significantly greater response of 63.0% relative to sham response of 31.6% (*p* < 0.001) (Post hoc OR 3.70 (LB 1.82, UB 7.52)). In clinical remission, active treatment arm showed a significantly greater remission rate 57.5% relative to sham 29.4% (*p* = 0.002) (Post hoc OR 3.26 (LB 1.53, UB 6.94).

Based on self-report MADRS-s ratings, active tDCS treatment arm showed significant improvement from baseline to week 10, mean improvement 9.9 ± 8.94, relative to sham, improvement 6.2 ± 9.13 (*p* = 0.009). In clinical response, active treatment arm showed a significantly greater response of 49.1% relative to sham response of 24.0% (*p* = 0.004) (Post hoc OR 3.06 (LB 1.43, UB 6.56)). In clinical remission, active treatment arm showed significantly greater remission rate 53.8% relative to sham 23.4% (*p* = 0.002) (Post hoc OR 3.83 (LB 1.61, UB 9.13) (Table 2).

**Table 1.** Demographic and clinical characteristics of patients at baseline.

| **Characteristic** | **Active**<br>**(N=87)** | **Sham**<br>**(N=87)** |
| --- | --- | --- |
| Age | 37.1 ± 11.1 | 38.3 ± 10.9 |
| Gender |  |  |
| Female | 54 (62) | 66 (76) |
| Race |  |  |
| Asian | 9 (10) | 2 (2) |
| Black or African American | 3 (3) | 1 (1) |
| Native Hawaiian or Other | 0 (0) | 0 (0) |
| White | 72 (83) | 73 (84) |
| Other | 3 (3) | 11 (13) |
| Missing | 0 (0) | 0 (0) |
| Educational Level |  |  |
| Less than High School/Secondary School | 1 (1) | 0 (0) |
| Some College | 18 (21) | 19 (22) |
| Diploma | 9 (10) | 7 (8) |
| Bachelor's or Professional Degree | 37 (43) | 37 (43) |
| Master's or Doctoral Degree | 22 (25) | 23 (26) |
| Prefer not to answer/Missing | 0 (0) | 1 (1) |
| Age of onset | 22.1 ± 9.7 | 22.4 ± 8.8 |
| Previous number of episodes | 4.1 ± 5.3 | 4.8 ± 5.7 |
| Previous number of suicide attempts | 0.10 ± 0.34 | 0.16 ± 0.43 |
| First episode MDD | 18 (21) | 10 (11) |
| Clinical ratings |  |  |
| HDRS | 19.2 ± 2.8 | 18.9 ± 2.6 |
| MADRS | 24.7 ± 4.7 | 23.9 ± 5.5 |
| MADRS-s | 26.8 ± 6.9 | 25.7 ± 6.3 |
| HAMA | 15.4 ± 4.6 | 14.3 ± 4.6 |
| YMRS | 2.1 ± 1.7 | 1.9 ± 1.6 |
| RAVLT | 57.9 ± 11.2 | 58.5 ± 13.4 |
| SDMT | 52.3 ± 10.1 | 50.4 ± 10.1 |
| Antidepressant medication during trial | 56 (64) | 53 (61) |
| Selective serotonin reuptake inhibitor | 40 (46) | 35 (40) |
| Non-selective monoamine reuptake inhibitor | 1 (1) | 3 (3) |
| Other antidepressant medications | 18 (21) | 17 (20) |
| Individual psychotherapy during trial | 12 (14) | 14 (16) |
| No antidepressant or psychotherapy during trial | 25 (29) | 32 (37) |

Categorical variables are presented as number of participants with percentage in parentheses for gender, race, educational level, first episode MDD, antidepressant medication and individual medications, individual psychotherapy during trial and No antidepressant or psychotherapy during trial. Mean values are presented with '±' standard deviation values. Diploma, a certificate that signifies a certain level of education and practical experience. HDRS, Hamilton Depression Rating Scale; MADRS, Montgomery-Åsberg Depression Rating Scale; MADRS-s, Montgomery-Åsberg Depression Rating Scale-self report; HAMA, Hamilton Anxiety Rating Scale; YMRS, Young Mania Rating Scale; RAVLT, Rey Auditory Verbal Learning Test; SDMT, Symbol Digit Modalities Test. SDMT active, n=85, SDMT sham, n=85. Age of onset, active n=86, sham n=86. HDRS scores range from 0 to 52, MADRS scores range from 0 to 60, MADRS-s scores range from 0 to 54, with higher scores indicating more depression. RAVLT scores range from 0 to 75. SDMT scores range from 0 to 110. A significant difference between groups was found for race, $p = 0.012$ . There were no significant differences any other characteristics

**Table 2.**
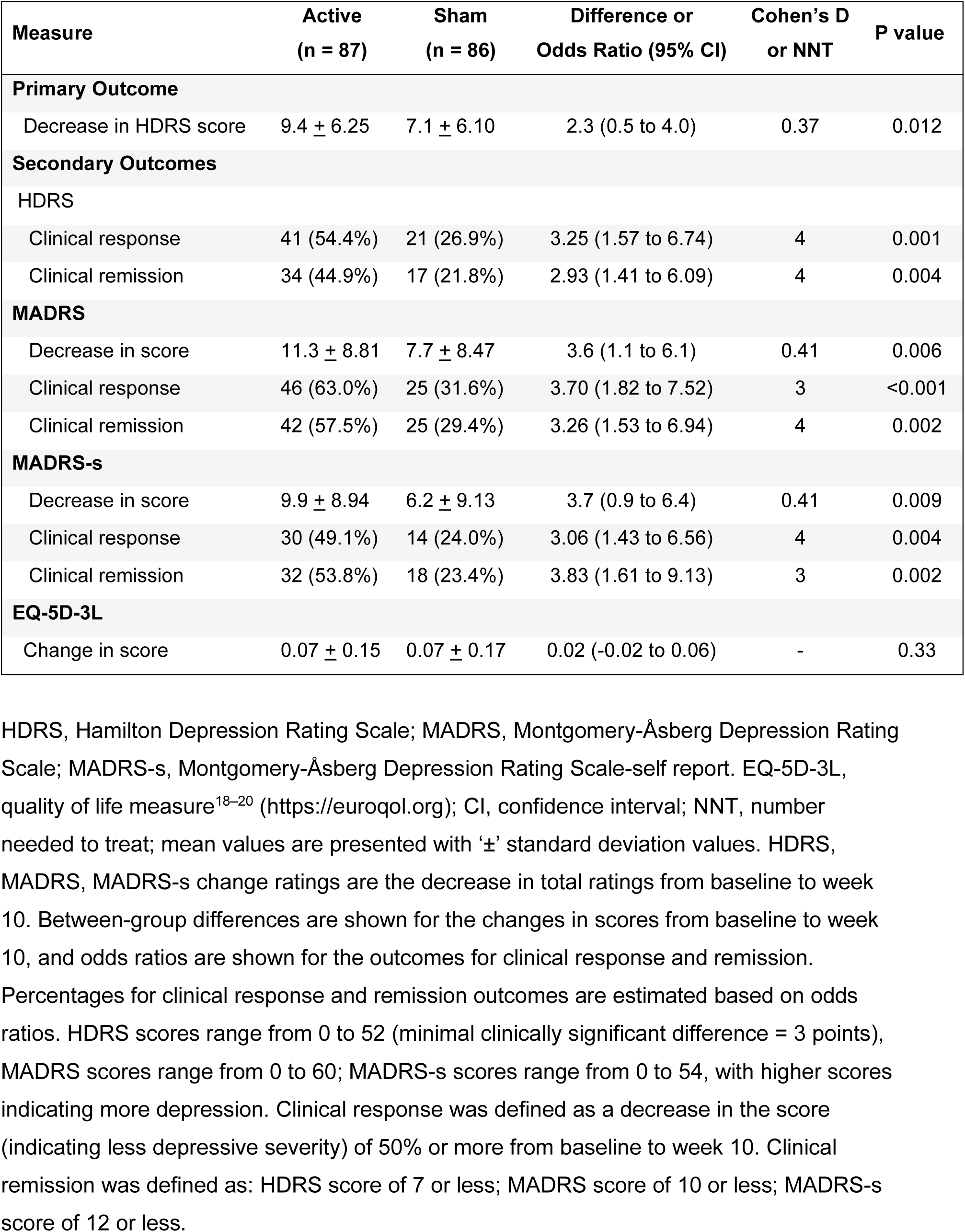
Primary and secondary outcomes: changes in depressive severity as measured by HDRS, MADRS and MADRS-s and quality of life as measured by EQ-5D-3L following a 10-week course of active or sham tDCS sessions.

There was no significant difference in quality of life between treatment arms as measured by EQ-5D-3L (*p* = 0.33).

### Exploratory Outcomes

In anxiety symptoms, there were no significant differences between active, mean HAMA improvement 6.6 ± 6.1 (mean HAMA 8.2 ± 5.7), relative to sham treatment arm 4.9 ± 5.9 (mean HAMA 9.3 ± 4.9) (*p* = 0.08). In hypomanic symptoms, YMRS mean score was 1.3 ± 1.4 in active treatment arm at week 10 and 1.8 ± 1.7 in sham treatment arm, which was statistically significant (*p* = 0.03).

In active treatment arm, 78% participants thought they were receiving active tDCS, and in the sham treatment arm, 59% participants thought they were receiving active tDCS.

In neuropsychological assessments, there were no significant differences in RAVLT or SDMT between treatment arms.

### Adverse events and safety

At week 10, there were increased reports of skin redness (*p* < 0.001), skin irritation (difference 6.9% (1.9% to 14.5%) *p* = 0.03) and trouble concentrating (*p* =0.03) in active relative to sham, and no differences in headache, neck pain, scalp pain, itching, burning sensation, sleepiness, or acute mood changes between treatment arms. Two participants in the active group reported burns at the left electrode site from using sponges which had dried out. Both burns healed, and neither developed into skin lesions. Both participants had informed the research team at the next study visit and were advised that they could take a break until the burn had healed. There were no serious adverse events related to the device, and no participants developed mania or hypomania.

## Discussion

In this international, sham-controlled RCT, active tDCS stimulation was associated with significantly greater improvements in depressive symptoms, greater clinical response rates, and greater remission rates relative to sham stimulation at 10 weeks. Improvements were evident for participants who were medication-free as well as participants who were taking regular antidepressant medication. The effects were evident at week 10, supporting a recent IPD analysis which found that tDCS effect sizes continue to increase up to 10 weeks as compared to sham stimulation.^27^

The findings support clinic-based studies, in which active tDCS shows greater efficacy than sham tDCS in MDD, particularly in first episode and recurrent MDD.^5,28–30^ In a recent large trial though, Bukhardt et al.^31^ did not observe any significant effects in a 6-week trial of adjunctive tDCS treatment to antidepressant medication. However, the trial had included participants with a history of poor treatment response to multiple antidepressant medications, and treatment resistant depression is negatively correlated with clinical efficacy^5,28–30^.

The present protocol demonstrated greater efficacy and safety relative to recent home-based tDCS trials.^32,33^ One trial had terminated early, enrolling 11 MDD participants, due to adverse events of skin lesions from an accumulation of skin burns.^32^ Another trial was a single-blind RCT of tDCS augmentation to antidepressant medication, consisting of hybrid clinic- and home-based tDCS sessions, which found improved self-report depressive symptoms but not in clinician-based ratings at 6 weeks.^33^ The present protocol was a fully remote, double-blind, sham-controlled RCT with real-time clinical assessments by video conference.

Safety was monitored using real-time assessments through video conference and the availability of a dedicated study number with 24-hour access to researchers. Electrical burns are an unanticipated side effect, usually resulting from application of tap water to moisten sponges,^34,35^ insufficient moistening with conductive saline solution,^36^ or pre-existing skin lesions. We had two cases of reported skin burn, both due to insufficient sponge moistening, but neither developed into skin lesions and participants continued treatment. There were no serious adverse events related to the device and no incidents of serious suicide risk. Active stimulation though was associated with higher rates of skin redness, irritation and dry skin relative to sham.^23,37^

Limitations include a predominantly white ethnicity in the sample, limited sub-group analysis as antidepressant medication type was not controlled for and impedance and current intensity were not analyzed. As history of hospital admissions and treatment resistant depression were exclusion criteria, the findings may not be generalisable to these groups.

In summary, our protocol of home-based tDCS demonstrated significant clinical efficacy, response and remission in a 10-week course of treatment. MDD participants had at least a moderate severity of depressive symptoms, but treatment resistant depression was not included. Home-based tDCS could be a potential first line treatment for MDD that demonstrates efficacy and safety, but consideration of continuing safety monitoring is required.

**Table 3.**
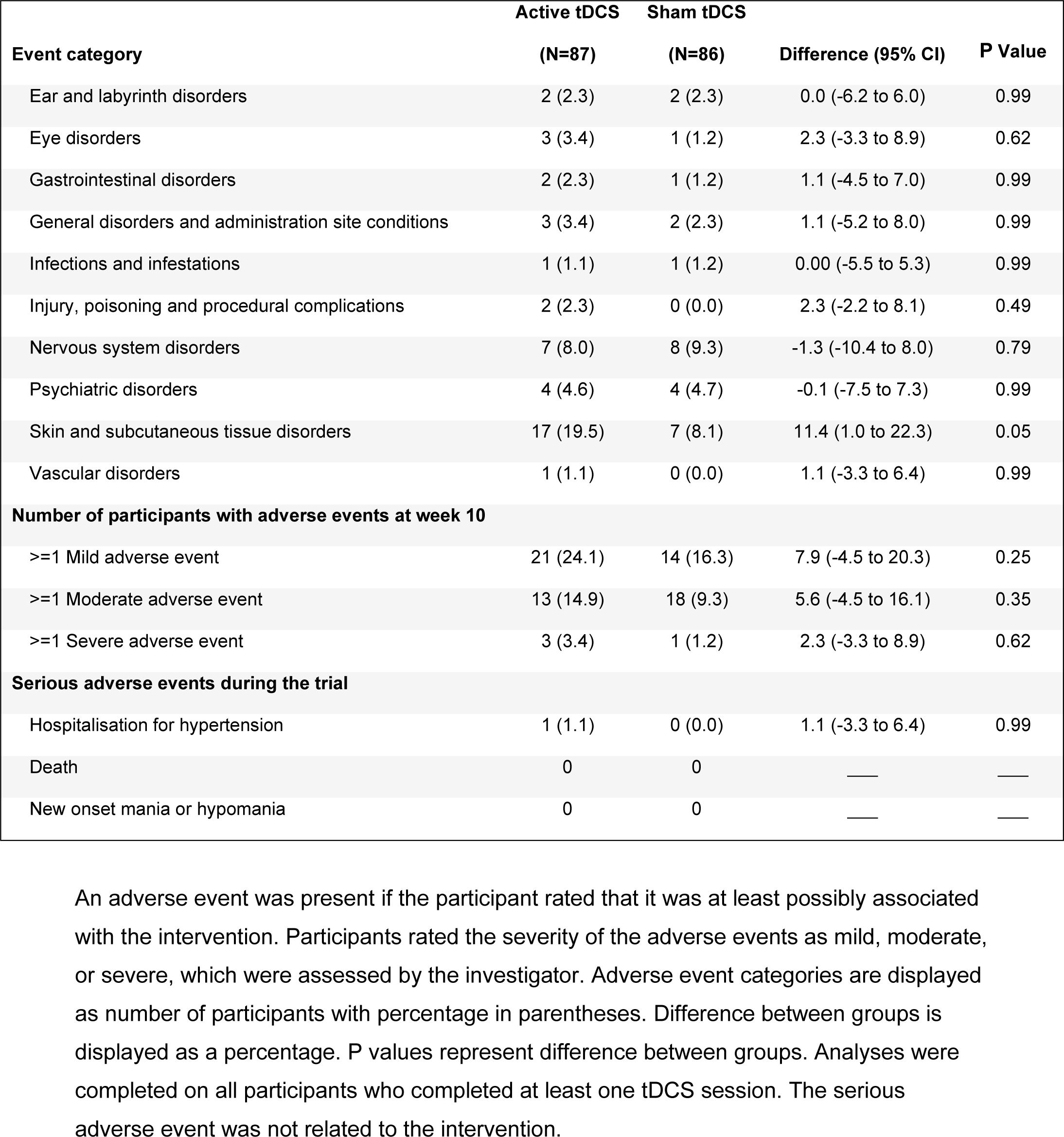
Unanticipated adverse events at 10 weeks.

| Event category | Active tDCS | Sham tDCS | Difference (95% CI) | P Value |
| --- | --- | --- | --- | --- |
|  | (N=87) | (N=86) |  |  |
| Ear and labyrinth disorders | 2 (2.3) | 2 (2.3) | 0.0 (-6.2 to 6.0) | 0.99 |
| Eye disorders | 3 (3.4) | 1 (1.2) | 2.3 (-3.3 to 8.9) | 0.62 |
| Gastrointestinal disorders | 2 (2.3) | 1 (1.2) | 1.1 (-4.5 to 7.0) | 0.99 |
| General disorders and administration site conditions | 3 (3.4) | 2 (2.3) | 1.1 (-5.2 to 8.0) | 0.99 |
| Infections and infestations | 1 (1.1) | 1 (1.2) | 0.00 (-5.5 to 5.3) | 0.99 |
| Injury, poisoning and procedural complications | 2 (2.3) | 0 (0.0) | 2.3 (-2.2 to 8.1) | 0.49 |
| Nervous system disorders | 7 (8.0) | 8 (9.3) | -1.3 (-10.4 to 8.0) | 0.79 |
| Psychiatric disorders | 4 (4.6) | 4 (4.7) | -0.1 (-7.5 to 7.3) | 0.99 |
| Skin and subcutaneous tissue disorders | 17 (19.5) | 7 (8.1) | 11.4 (1.0 to 22.3) | 0.05 |
| Vascular disorders | 1 (1.1) | 0 (0.0) | 1.1 (-3.3 to 6.4) | 0.99 |
| **Number of participants with adverse events at week 10** |  |  |  |  |
| >=1 Mild adverse event | 21 (24.1) | 14 (16.3) | 7.9 (-4.5 to 20.3) | 0.25 |
| >=1 Moderate adverse event | 13 (14.9) | 18 (9.3) | 5.6 (-4.5 to 16.1) | 0.35 |
| >=1 Severe adverse event | 3 (3.4) | 1 (1.2) | 2.3 (-3.3 to 8.9) | 0.62 |
| **Serious adverse events during the trial** |  |  |  |  |
| Hospitalisation for hypertension | 1 (1.1) | 0 (0.0) | 1.1 (-3.3 to 6.4) | 0.99 |
| Death | 0 | 0 | — | — |
| New onset mania or hypomania | 0 | 0 | — | — |

An adverse event was present if the participant rated that it was at least possibly associated with the intervention. Participants rated the severity of the adverse events as mild, moderate, or severe, which were assessed by the investigator. Adverse event categories are displayed as number of participants with percentage in parentheses. Difference between groups is displayed as a percentage. P values represent difference between groups. Analyses were completed on all participants who completed at least one tDCS session. The serious adverse event was not related to the intervention.

**Table 4.** Anticipated adverse events at 10 weeks as measured by the tDCS Adverse Events Questionnaire.^23^.

| Adverse event category | Active (N=87) |  |  |  | Sham (N=86) |  |  |  | P Value |
| --- | --- | --- | --- | --- | --- | --- | --- | --- | --- |
|  | Total | Mild | Moderate | Severe | Total | Mild | Moderate | Severe |  |
| Headache | 36 (42) | 24 (28) | 11 (13) | 1 (1) | 29 (36) | 18 (22) | 9 (11) | 2 (2) | 0.42 |
| Neck Pain | 2 (2) | 0 (0) | 2 (2) | 0 (0) | 4 (5) | 1 (1) | 3 (4) | 0 (0) | 0.43 |
| Scalp pain | 18 (21) | 14 (16) | 3 (4) | 1 (1) | 10 (12) | 7 (9) | 3 (4) | 0 (0) | 0.15 |
| Itching | 43 (51) | 37 (44) | 3 (4) | 3 (4) | 35 (43) | 28 (35) | 7 (9) | 0 (0) | 0.08 |
| Burning sensation | 37 (44) | 32 (38) | 4 (5) | 1 (1) | 31 (38) | 25 (31) | 6 (7) | 0 (0) | 0.42 |
| Skin redness | 54 (64) | 42 (49) | 11 (13) | 1 (1) | 15 (19) | 13 (16) | 2 (2) | 0 (0) | <0.001 |
| Sleepiness | 10 (12) | 5 (6) | 4 (5) | 1 (1) | 12 (15) | 9 (11) | 2 (2) | 1 (1) | 0.65 |
| Trouble concentrating | 12 (14) | 8 (9) | 3 (4) | 1 (1) | 3 (4) | 2 (2) | 1 (1) | 0 (0) | 0.03 |
| Acute mood change | 7 (8) | 3 (4) | 3 (4) | 1 (1) | 6 (7) | 5 (6) | 1 (1) | 0 (0) | 1.00 |

Values are number of participants with percentage in parentheses. An adverse event was present if the participant rated that it was at least remotely possible that it was associated with the intervention. Participants rated the severity of the adverse events as mild, moderate, or severe. P values represent group differences of the total number of events per event category.

## Data Availability

None of the data in the present manuscript will be made available.

## Notes

### Competing Interest Statement

AHY reports declaration of interests: Employed by Kings College London; Honorary Consultant South London and Maudsley NHS Foundation Trust (NHS UK). Editor of Journal of Psychopharmacology and Deputy Editor, BJPsych Open. Paid lectures and advisory boards for the following companies with drugs used in affective and related disorders: Flow Neuroscience, Novartis, Roche, Janssen, Takeda, Noema pharma, Compass, Astrazenaca, Boehringer Ingelheim, Eli Lilly, LivaNova, Lundbeck, Sunovion, Servier, Livanova, Janssen, Allegan, Bionomics, Sumitomo Dainippon Pharma, Sage, Novartis, Neurocentrx. Grant funding (past and present): NIMH (USA); CIHR (Canada); NARSAD (USA); Stanley Medical Research Institute (USA); MRC (UK); Wellcome Trust (UK); Royal College of Physicians (Edin); BMA (UK); UBC-VGH Foundation (Canada); WEDC (Canada); CCS Depression Research Fund (Canada); MSFHR (Canada); NIHR (UK). Janssen (UK) EU Horizon 2020. Principal Investigator on the following trials;1. the Restore-Life VNS registry study funded by LivaNova; 2. ESKETINTRD3004: An Open-label, Long-term, Safety and Efficacy Study of Intranasal Esketamine in Treatment-resistant Depression; 3. The Effects of Psilocybin on Cognitive Function in Healthy Participants; 4. The Safety and Efficacy of Psilocybin in Participants with Treatment-Resistant Depression (P-TRD); 5. A Double-Blind, Randomized, Parallel-Group Study with Quetiapine Extended Release as Comparator to Evaluate the Efficacy and Safety of Seltorexant 20 mg as Adjunctive Therapy to Antidepressants in Adult and Elderly Patients with Major Depressive Disorder with Insomnia Symptoms Who Have Responded Inadequately to Antidepressant Therapy. (Janssen); 6. An Open-label, Long-term, Safety and Efficacy Study of Aticaprant as Adjunctive Therapy in Adult and Elderly Participants with Major Depressive Disorder (MDD). (Janssen); 7. A Randomized, Double-blind, Multicentre, Parallel-group, Placebo-controlled Study to Evaluate the Efficacy, Safety, and Tolerability of Aticaprant 10 mg as Adjunctive Therapy in Adult Participants with Major Depressive Disorder (MDD) with Moderate-to-severe Anhedonia and Inadequate Response to Current Antidepressant Therapy; 8. A Study of Disease Characteristics and Real-life Standard of Care Effectiveness in Patients with Major Depressive Disorder (MDD) With Anhedonia and Inadequate Response to Current Antidepressant Therapy Including an SSRI or SNR. (Janssen). UK Chief Investigator for Compass; COMP006 & COMP007 studies. UK Chief Investigator for Novartis MDD study MIJ821A12201. No shareholdings in pharmaceutical companies
SS received research grant funding from Flow Neuroscience to the University of Texas Health Science Center at Houston for the present manuscript. Additional disclosures are within the past 36 months. Research grant funding payments made to the University of Texas Health Science Center at Houston include: NIMH (1R21MH119441-01A1; 1R21MH129888-01A1), NICHD (1R21HD106779-01A1), SAMHSA (6H79FG000470-01M003). Payment or honoraria for lectures, presentations, speakers bureaus, manuscript writing or educational events for Psychiatry education forum and NIH study section reviewer. Participation on a Data Safety Monitoring Board or Advisory Board for Worldwide Clinical Trials and Inversago and Vicore pharma. Stock or Stock options in Intra-Cellular Therapies as a full time employee and Biogen. Participated in clinical trials as site sub-investigator - University of Texas Health Science Center at Houston for the following: COMPASS Pathways, LivaNova, Janssen and Relmada. Consulting fees from Boxer Capital. Royalties or licenses from Cambridge University Press (no payments have been received).
CHYF received research grant funding from Flow Neuroscience to the University of East London for the present manuscript. Additional disclosures are within the past 36 months. Research grant funding payments made to institutions include: Baszucki Brain Research Milken Institute (0000000029), Institute of Psychoanalysis (IPA0158), NIMH (RO1MH134236), Rosetrees Trust (CF20212104).
JCS disclosures from the past 36 months include grants or contracts from Milken, Compass Pathways and MindMed. Consulting fees from Livanova, Boehringer Ingleheim, Johnson & Johnson, and Sunovian. Participation on a Data Safety Monitoring Board or Advisory Board for Alkermes.
DM works for Biomedical Statistical Consulting. Biomedical Statistical Consulting provides statistical support to MCRA, LLC who received payments from Flow Neuroscience.
LH works for Biomedical Statistical Consulting. Biomedical Statistical Consulting provides statistical support to MCRA, LLC who received payments from Flow Neuroscience.
PL was paid to work on the trial for a period of approximately 3 months as an Assistant Researcher at a level commensurate with that role.
Authors GS, AN, RW, NL, HH, MR, PO, RMV and SK declare that they have no competing interests to declare.

### Clinical Trial

NCT05202119

### Funding Statement

The study was funded by Flow Neuroscience.

### Author Declarations

In UK, South Central-Hampshire B Research Ethics Committee gave ethical approval for this work. In Texas, USA, WIRB-Copernicus Group International Review Board gave ethical approval for this work.

### Summary of Updates

Revision 1: Figure S2 in the supplementary appendix 'Change in Montgomery-Asberg Depression Rating Scale (MADRS) ratings over time' had an incorrect label on the y axis. This has been updated from 'MADRS-s score' to the correct label 'MADRS score'. Revision 2: Figure S9 in the supplementary appendix 'Percentage of participants in active and sham groups who endorsed each response in the acceptability questionnaire at baseline and week 10' had the incorrect labels for the 'Burden' bar chart. 'More effort than usual' and 'Less effort than usual' had been positioned on the wrong sides of the x axis. This has been updated and the 'Burden' bar chart now correctly reflects the responses.

