## Supplementary Appendix for "Home-based transcranial direct current stimulation RCT in major depression"

Home-based transcranial direct current stimulation RCT in major depression

### List of investigators

Rachel D. Woodham, M.Sc. [a], Sudhakar Selvaraj, M.B.B.S., D.Phil. [b, c], Nahed Lajmi, M.Sc. [a], Harriet Hobday, M.Sc. [a], Gabrielle Sheehan, M.Sc. [a], Ali-Reza Ghazi-Noori, M.Sc. [a], Peter J. Lagerberg, M.Sc. [a], Maheen Rizvi, B.Sc. [b], Sarah S. Kwon, B.Sc. [b], Paulette Orhii, B.Sc. [b], David Maislin [d], Lucia Hernadez, Ph.D. [d], Rodrigo Machado-Vieira, M.D., Ph.D. [b], Jair C. Soares, M.D., Ph.D. [b], Allan H. Young MB, ChB., Ph.D. [e, f, g], Cynthia H.Y. Fu, M.D., Ph.D. [a, e]

**Affiliations:**

a) School of Psychology, University of East London, London, UK
b) Center of Excellence on Mood Disorders, Faillace Department of Psychiatry and Behavioral Sciences, McGovern Medical School, University of Texas Health Science Center at Houston, Houston, TX, USA
c) Intra-Cellular Therapies Inc., USA
d) Biomedical Statistical Consulting, Philadelphia, PA, USA
e) Centre for Affective Disorders, Institute of Psychiatry, Psychology and Neuroscience, King’s College London, London, UK
f) National Institute for Health Research Biomedical Research Centre at South London and Maudsley NHS Foundation Trust, King's College London, London, UK
g) South London and Maudsley NHS Foundation Trust, Bethlem Royal Hospital, Beckenham, UK

**Supplementary Appendix**

Home-based transcranial direct current stimulation RCT in major depression

### Inclusion and exclusion criteria

Inclusion criteria:

1. Be >=18 years.
2. Have a diagnosis of Unipolar MDD with a current depressive episode as defined by the diagnostic criteria in the Diagnostic and statistical manual of mental disorders – 5th edition (DSM-V)
3. Have a Hamilton Depression Rating Score (HDRS) of >=16.
4. For 6 weeks prior to enrolment, are either: a. not taking antidepressant medication or: b. are taking a stable antidepressant regimen with a stable medication source and agree to continue the same regimen throughout study participation.
5. If in psychotherapy, have maintained stable psychotherapy for at least 6 weeks prior to enrolment.
6. Have access to a stable internet connection through which the treatment will be received.
7. Have access to a smartphone or other device running Android 5.0+ or iPhone Operating System (iOS) 12+ (e.g., reasonably new iPhone/iPad or Android phone), used to using the device in their everyday life, and can capably use the study application on the device, as determined by the investigator.
8. Are currently living in England/Wales (UK) or Texas (US).
9. Subject is currently under the care of a psychiatrist or a primary care physician, agrees to be evaluated at regular intervals by a psychiatrist or primary care physician for the duration of study participation, and agrees to promptly inform the study staff of any change of psychiatric or mental health providers during study participation.
10. Subject agrees to allow any and all forms of communication between the investigators/study staff and any healthcare provider who currently provides and/or has provided service to the patient/subject within at least two years of study enrolment.
11. Subject agrees to provide the name and verifiable contact information (email and mailing addresses, mobile and land-line phone numbers, as applicable) of at least two persons ≥ age 18 (22 in the US) who reside within a 60-minute drive of the patient's residence and whom the research staff is at liberty to contact, as they deem necessary, for the duration of study participation
12. Be able to give voluntary, written informed consent to participate and have signed an Informed Consent Form specific to this study.
13. Be willing and able to comply with all study procedures.
14. Subject agrees to meet all of the inclusion criteria throughout their participation in the study. Otherwise, the subject will be discontinued from the study.
15. Subject agrees to a Safety/Suicide Risk Management Protocol, which is intended to reduce the reduce the risk of suicide during study participation.

Exclusion criteria:

1. Are in a current state of mania, as determined by the YMRS or psychosis, as determined by the MINI.
2. Are diagnosed with vitamin or hormonal deficiencies that may mimic mood disorders, as determined by the investigator.
3. Are currently receiving any other interventional therapy for MDD other than a stable regimen of antidepressants or psychotherapy as defined in the inclusion criteria.
4. Considered to have treatment resistant depression as defined by inadequate clinical response to 2 or more trials of antidepressants at an adequate dose and duration.
5. Have a history of electroconvulsive therapy (ECT), transcranial magnetic stimulation (TMS), cranial electrotherapy stimulation (CES), transcranial direct current stimulation (tDCS), deep brain stimulation (DBS), or other brain stimulation.

6. Patient answers Yes to Questions 4, 5 or 6 on the Columbia Suicide Severity Rating Scale (C-SSRS) Triage and Risk Identification Screener.
7. Any previous hospitalization for suicidal behavior.
8. Have chronic severe insomnia (< 4 hours of sleep each night), or depression secondary to chronic insomnia or sleep apnea.
9. Have any structural lesion (e.g., any structural neurological condition, or more subcortical lesions than would be expected for age or have had a stroke that affects stimulated area or connected areas) or any other clinically significant abnormality that might affect safety, study participation, or confound interpretation of study results, as determined by the investigator.
10. Have any implant in the brain (e.g., DBS) or neurocranium, or any other active implantable medical device.
11. Have any neurocranial defect.
12. Have a history of epilepsy or seizures (including history of withdrawal / provoked seizures).
13. Have shrapnel or any ferromagnetic material in the head.
14. Have any disorder that would impair the ability to complete the study questionnaires.
15. Have been diagnosed with autism spectrum disorder.
16. Are actively abusing substances (<1 week prior to enrolment).
17. Have a cognitive impairment (including dementia).
18. Have a history of mania or psychosis.
19. Are currently using any medications that affect cortical excitability (e.g., benzodiazepines, epileptics, etc.).
20. Are currently experiencing symptoms of withdrawal from alcohol or benzodiazepines.
21. Have been diagnosed with Parkinsonism or other movement disorder as determined by the investigator to interfere with treatment.
22. Have ever taken esketamine / ketamine for treatment of depression.
23. Have ever been admitted to hospital for depression.
24. Have ever been diagnosed with obsessive-compulsive disorder (OCD) or bipolar type 1 or 2 disorder.
25. Is diagnosed with an active primary anxiety disorder, or PTSD, agoraphobia, anorexia or bulimia, panic or personality disorder with active symptoms.
26. Have a history of psychosurgery for depression.
27. Have any history of myocardial infarction, coronary artery bypass graft (CABG), coronary heart failure (CHF), or history of other cardiac issues.
28. Are currently experiencing or have a history of intractable migraines.
29. Are a chronic tobacco smoker, as defined by smoking by smoking >100 cigarettes (including hand-rolled cigarettes, cigars, cigarillos, etc.) in their life-time and have smoked every day for the last 7 days.
30. If female and of child-bearing potential, currently pregnant or breastfeeding or planning to become pregnant or breastfeed any time during the study.
31. Are currently a prisoner.
32. Are participating concurrently in another clinical investigation or have participated in a clinical investigation within the last 90 days or intend to participate in another clinical investigation during the study, and where the participation in the other investigation might interfere with the results of this trial as deemed by the PI.
33. Have any medical condition or other circumstances, in the judgment of the investigator, that might interfere with the ability to complete follow-up visits and the self-reported MADRS-s in the app.
34. Have any condition which, in the judgment of the Investigator, would preclude adequate evaluation of the device’s safety and performance.
35. A Subject who meets any of the exclusion criteria during study participation will be discontinued from the study.

**Previous exclusion criteria which had been amended following trial commencement (protocol version 7, August 31, 2022):**

Criteria 24 and 25:

This criterion was amended to the current exclusion criteria 24 and 25. Criteria 24 separated out lifetime OCD and bipolar disorder, and criteria 25 reflected current psychiatric disorders, including anorexia and bulimia:

- Have been diagnosed with obsessive-compulsive disorder (OCD), bipolar type 1 or 2 disorder, an active primary anxiety disorder, PTSD, agoraphobia, panic or personality disorder.

Criterion 8:

This criterion was amended to current exclusion criterion number 8 to clarify that depression was secondary to insomnia or sleep apnea:

- Have chronic or current severe insomnia (< 4 hours of sleep each night), or sleep apnea.

**Supplementary Appendix**

Home-based transcranial direct current stimulation RCT in major depression

### Figure S1. Consort flow diagram

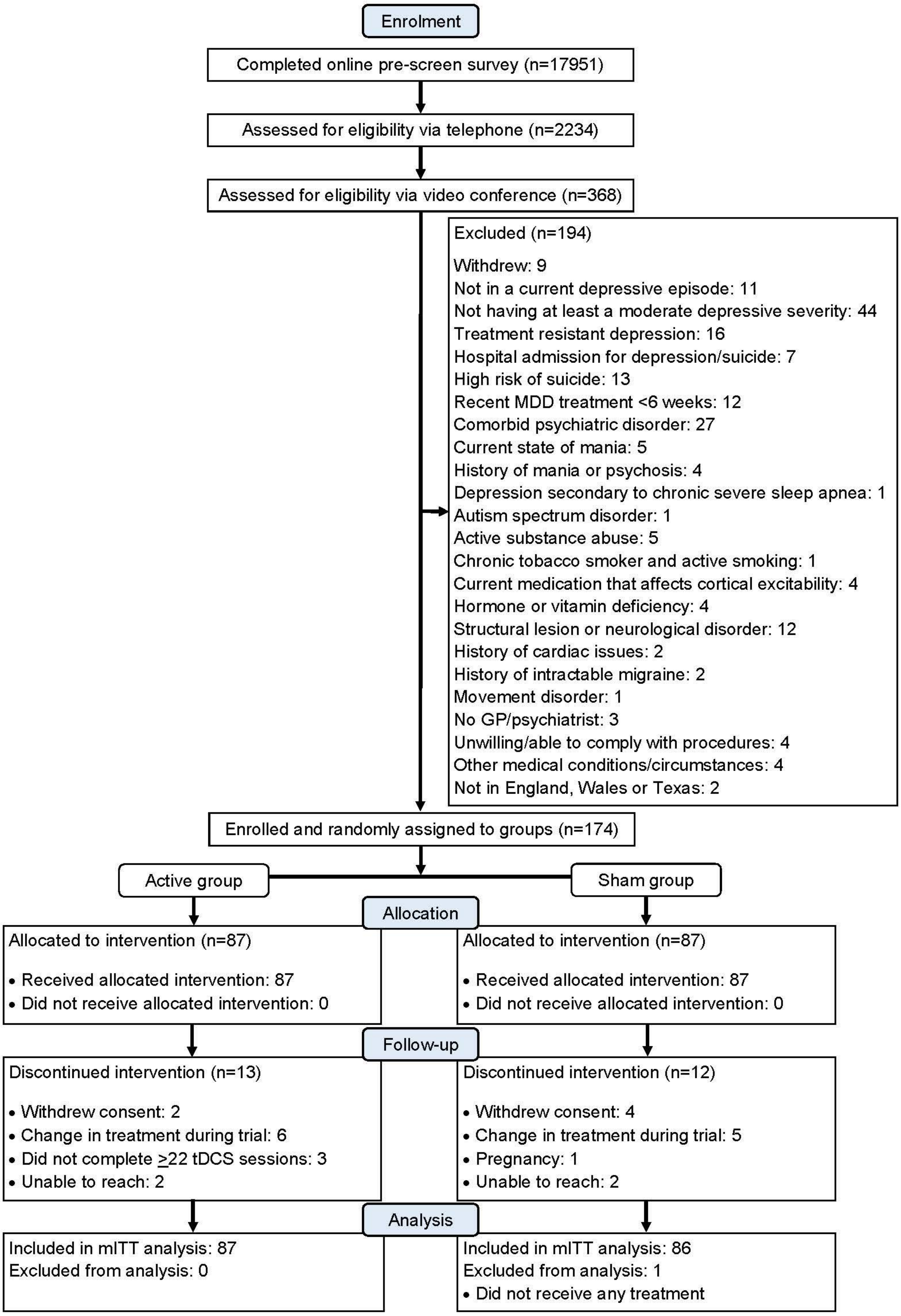

Enrolment, group allocation, follow-up and analysis are presented in the CONSORT flow-diagram. MDD, major depressive disorder; tDCS, transcranial direct current stimulation; mITT, modified intention-to-treat; CONSORT, consolidated standards of reporting trials

**Supplementary Appendix**

Home-based transcranial direct current stimulation RCT in major depression

### Participant recruitment

Pre-screening assessment consisted of two stages. A potential participant would complete an initial online questionnaire and register in the clinical research organisation (CRO), through the landing page. Participants provided their email, telephone, name and zip code, and answered questions related to current antidepressant treatment and medical history. From the initial questionnaire, a CRO coordinator would arrange a telephone call with the potential participant. During the telephone call, the CRO would ask a series of standardised questions and document the answers in the CRO platform. From the telephone assessment, contact details of potentially eligible participants would be referred to the research site.

The US site had two additional recruitment streams using third party companies. One stream followed the same two pre-screening stages and participants would be referred to the research site if they were deemed potentially eligible after the telephone assessment. The second stream used a software search of their patient database based on study inclusion/exclusion criteria. Potentially suitable patients were identified, and the software sent requests for these patients to fill in a form where they expressed interest in participating in the study. If the patient completed the form, they were automatically added as a participant to be interviewed by the research site.

#### Table S1. Recruitment figures and success rates for both sites at each time point

| Outcome (N) | Pre-screen questionnaire | Pre-screen telephone interview | Screening interview |
|---|---|---|---|
| **UK** | | | |
| Eligible | 4954 | 741 | 121 |
| Ineligible | 5211 | 900 | 119 |
| Total | 10165 | 1641 | 240 |
| Step eligible % | **48.74%** | **45.16%** | **50.42%** |
| Eligible % vs all | 48.74% | 7.29% | 1.19% |
| **USA** | | | |
| Eligible | 1582 | 238 | 61 |
| Ineligible | 6204 | 355 | 67 |
| Total | 7786 | 593 | 128 |
| Step eligible % | **20.32%** | **40.13%** | **47.66%** |
| Eligible % vs all | 20.32% | 3.06% | 0.78% |
| **Total both sites** | | | |
| Eligible | 6536 | 979 | 182 |
| Ineligible | 11415 | 1255 | 186 |
| Total | 17951 | 2234 | 368 |
| Step eligible % | **36.41%** | **43.82%** | **49.46%** |
| Eligible % vs all | 36.41% | 5.45% | 1.01% |

8 participants from the UK site who were eligible at the screening interview withdrew before randomisation.

### Safety and tolerability

A safety/suicide risk management protocol was implemented to mitigate suicide risk throughout the study. Prior to participating, all individuals consented to follow the protocol, which involved sharing contact information for at least two adults (age 18 or older in UK, or age 22 or older in USA), residing within an hour's drive, and who could be contacted by the research staff if necessary. The Brown Safety Plan (2012)[1] served as a template. Participants and researchers worked through the plan together to record helpful crisis management strategies and to provide a list of contacts, including the nearest hospital with an emergency department, their GP or psychiatrist contact details, and a suicide prevention helpline.

Columbia-Suicide Severity Rating Scale (C-SSRS)[2] was assessed at each study visit to monitor suicide risk. If a participant had had active suicidal thoughts or any plans, they would be withdrawn from the study, their GP or psychiatrist would be notified, and they would be advised to visit the nearest emergency department or mental health crisis centre. A Hamilton Depression Rating Scale (HDRS)[3] score increase of 30% from baseline would be considered a treatment failure and the participant's GP or psychiatrist would be contacted. The Young Mania Rating Scale (YMRS)[4] was used to assess manic and hypomanic symptoms at each visit.

### Statistical methods

Exploratory endpoints were analyzed through summary statistics as means and standard deviations (SD) or percentages and odds ratios. The two groups were compared through Student's t Test or Fisher Exact Test as appropriate. Spearman correlation alone was used to assess the association between two continuous variables. 95% confidence intervals (CI) were presented.

The percentages of subjects that correctly guessed the arm that they were in were compared through Fisher Exact Test.

Subgroup analyses of the primary and secondary endpoints were conducted through stratification by antidepressant usage at baseline and site.

### Table S2. Demographic and clinical characteristics of patients at baseline (complementary information)

| Characteristic | Active (N=87) | Sham (N=87) |
|---|---|---|
| Age by gender | | |
| Male | 39.9 ± 11.7 | 41.7 ± 11.0 |
| Female | 35.4 ± 10.5 | 37.2 ± 10.8 |
| Ethnicity | | |
| Hispanic or Latino | 7 (8) | 13 (15) |
| Not Hispanic or Latino | 77 (89) | 70 (80) |
| Not reported | 3 (3) | 3 (3) |
| Missing | 0 (0) | 1 (1) |

Ethnicity is presented as number of participants with percentage in parentheses. Mean values are presented with '±' standard deviation values.

### Table S3. Concomitant medications ongoing at randomisation and medical history

| Concomitant medication | On-going at randomization Active | On-going at randomization Sham | Medical history Active | Medical history Sham |
|---|---|---|---|---|
| Proton pump inhibitors | 0 (0) | 1 (1.1) | 0 (0) | 0 (0) |
| Aldosterone antagonists | 0 (0) | 1 (1.1) | 0 (0) | 0 (0) |
| Beta blocking agents, non-selective | 2 (2.3) | 3 (3.4) | 1 (1.1) | 0 (0) |
| Thyroid hormones | 2 (2.3) | 1 (1.1) | 0 (0) | 0 (0) |
| Other muscle relaxants, peripherally acting agents | 0 (0) | 0 (0) | 0 (0) | 0 (0) |
| Other centrally acting agents | 1 (1.1) | 0 (0) | 0 (0) | 0 (0) |
| Phenothiazines with piperazine structure | 0 (0) | 0 (0) | 0 (0) | 0 (0) |
| Diazepines, oxazepines, thiazepines and oxepines | 0 (0) | 1 (1.1) | 0 (0) | 0 (0) |
| Other antipsychotics | 0 (0) | 1 (1.1) | 1 (1.1) | 0 (0) |
| Benzodiazepine derivatives | 0 (0) | 0 (0) | 0 (0) | 0 (0) |
| Diphenylmethane derivatives | 1 (1.1) | 0 (0) | 0 (0) | 0 (0) |
| Azaspirodecanedione derivatives | 1 (1.1) | 0 (0) | 0 (0) | 0 (0) |
| Non-selective monoamine reuptake inhibitors | 1 (1.1) | 3 (3.4) | 0 (0) | 1 (1.1) |
| Selective serotonin reuptake inhibitors | 40 (46) | 35 (40.2) | 13 (14.9) | 17 (19.5) |
| Other antidepressants | 18 (20.7) | 17 (19.5) | 4 (4.6) | 6 (6.9) |
| Centrally acting sympathomimetics | 2 (2.3) | 2 (2.3) | 1 (1.1) | 1 (1.1) |
| Phenothiazine derivatives | 0 (0) | 1 (1.1) | 0 (0) | 0 (0) |

Medical history includes medications that ended in the 6-month period before the randomization date. One participant in the sham group was taking a selective serotonin reuptake inhibitor with unknown start and end dates.

### Interim analysis

An interim analysis was performed when 90 subjects had week 10 data and included both a futility assessment and sample size re-estimation. The futility assessment was based on stochastic curtailing approach by Lachin et al. (2005).[5] The sample size re-estimation will be based on a Promising Zone methodology by Mehta and Pocock (2011)[6] with a Fuzzy design approach introduced by Keenan and Maislin (2014).[7] Separate sample size assessments were performed to evaluate the secondary endpoint of clinical response. Adjustments to the trial for the secondary endpoint were only performed because the primary endpoint met the criteria as dictated by the promising zone criteria. Sample size assessments for the primary and secondary were characterized extensively through simulation analyses and pre-defined methods for controlling operational bias were submitted to FDA for review and agreement prior to the interim analysis be performed. The Interim analysis results had the capacity to modify the trial in two ways for the primary endpoint: 1) Declare the trial futile and stop enrolment, 2) specify a number of subjects between 100 and 270 that is needed for powering the trial. The interim analysis based on the Fuzzy Promising Zone concluded that 6 additional subjects were needed. An analysis was used performed showing that subjects before and after the interim were similar and considered exchangeable.

### Withdrawal rates and reasons for withdrawals

At week 10, 53 participants completed all the 36 planned tDCS sessions (28 in active treatment arm and 25 in sham treatment arm), and 153 participants completed the minimum number of 22 sessions (60% of total number of session) (76 in active treatment arm and 77 in sham treatment arm sham).

25 participants were withdrawn from the study (14.3%), n=13 in active group (14.9%), n=12 in sham group (13.7%) (p = 0.99) during the initial 10-week blinded phase.

### Table S4. Reasons for study withdrawal in the active and sham tDCS groups during the blinded phase of the trial

| Active tDCS | Sham tDCS |
|---|---|
| Change in antidepressant treatment during trial (5) | Change in antidepressant treatment during trial (4) |
| Change in ADHD treatment during trial (1) | Change in pain medication during trial (1) |
| Missed a large number of sessions (3) | Withdrew consent due to worsening of condition (2) |
| Withdrew consent due to AE – headache and fatigue (1) | Withdrew consent due to AE – headaches (1) |
| Withdrew consent due to AE – nausea and fatigue (1) | Withdrew consent due to AE – tinnitus (1) |
| Unable to contact participant (2) | Pregnancy (1) |
| | Did not receive any treatment and unable to reach participant (1) |
| | Unable to contact participant (1) |

tDCS, transcranial direct current stimulation. ADHD, Attention deficit hyperactivity disorder. AE, adverse event

### Table S5. Hamilton Depression Rating Scale estimated scores at each timepoint from baseline to week 10

| Outcome | Active tDCS (n = 87) | Sham tDCS (n = 86) |
|---|---|---|
| **HDRS** | | |
| Baseline score | 19.4 ± 3.42 | 19.1 ± 3.46 |
| Week 1 score | 15.4 ± 4.97 | 14.6 ± 5.04 |
| Week 4 score | 11.8 ± 5.50 | 13.4 ± 5.48 |
| Week 7 score | 10.9 ± 5.96 | 12.2 ± 5.91 |
| Week 10 score | 9.6 ± 6.02 | 11.7 ± 5.96 |

HDRS, Hamilton Depression Rating Scale,[3] Mean values are presented with '±' standard deviation values. HDRS scores range from 0 to 52 (minimal clinically significant difference = 3 points), with higher scores indicating more depression.

### Table S6. Montgomery-Åsberg Depression Rating Scale estimated scores at each timepoint from baseline to week 10

| Outcome | Active tDCS (n = 87) | Sham tDCS (n = 86) |
|---|---|---|
| **MADRS** | | |
| Baseline score | 24.8 ± 6.45 | 24.0 ± 6.52 |
| Week 1 score | 20.3 ± 7.78 | 19.5 ± 7.90 |
| Week 4 score | 15.1 ± 8.26 | 17.8 ± 8.24 |
| Week 7 score | 13.9 ± 9.04 | 16.6 ± 9.00 |
| Week 10 score | 12.5 ± 9.40 | 15.3 ± 9.28 |

MADRS, Montgomery-Åsberg Depression Rating Scale,[13] Mean values are presented with '±' standard deviation values. MADRS scores range from 0 to 60, with higher scores indicating more depression.

### Table S7. Montgomery-Åsberg Depression Rating Scale-self report estimated scores at each timepoint from baseline to week 10

| Outcome | Active tDCS (n = 87) | Sham tDCS (n = 86) |
|---|---|---|
| **MADRS-s** | | |
| Baseline score | 27.4 ± 7.87 | 26.3 ± 7.95 |
| Week 1 score | 22.5 ± 8.94 | 22.9 ± 9.26 |
| Week 4 score | 18.9 ± 9.04 | 20.8 ± 9.14 |
| Week 7 score | 18.9 ± 9.44 | 19.9 ± 9.72 |
| Week 10 score | 16.6 ± 9.33 | 19.6 ± 9.62 |

MADRS-s, Montgomery-Åsberg Depression Rating Scale-self report,[14] Mean values are presented with '±' standard deviation values. MADRS-s scores range from 0 to 54, with higher scores indicating more depression.

**Supplementary Appendix**

Home-based transcranial direct current stimulation RCT in major depression

### Figure S2. Change in Montgomery-Åsberg Depression Rating Scale (MADRS) ratings over time

Shown are the estimated mean MADRS rating scores from baseline to week 10 in the modified intention-to-treat analysis sample (n=173) for the active tDCS and sham tDCS treatment arms. Error bars represent ± 1 standard error (SE). MADRS scores range from 0 to 60 with higher values indicating more severe depression. A significant improvement was observed in the change in MADRS ratings from baseline to week 10 in the active tDCS treatment arm, MADRS change 11.3 ± 8.81(SD) (mean week 10 MADRS 12.5 ± 1.1 (SE)) as compared to sham tDCS treatment arm, MADRS change 7.7 ± 8.47 (SD) (mean week 10 MADRS 15.3 ± 1.1 (SE)) (95% CI 1.1 to 6.1, *p* = 0.006). The difference in change scores was also significant at week 4 (*p* = 0.003) and week 7 (*p* = 0.005) with a greater score decrease in the active treatment arm. ** = *p* <0.01.

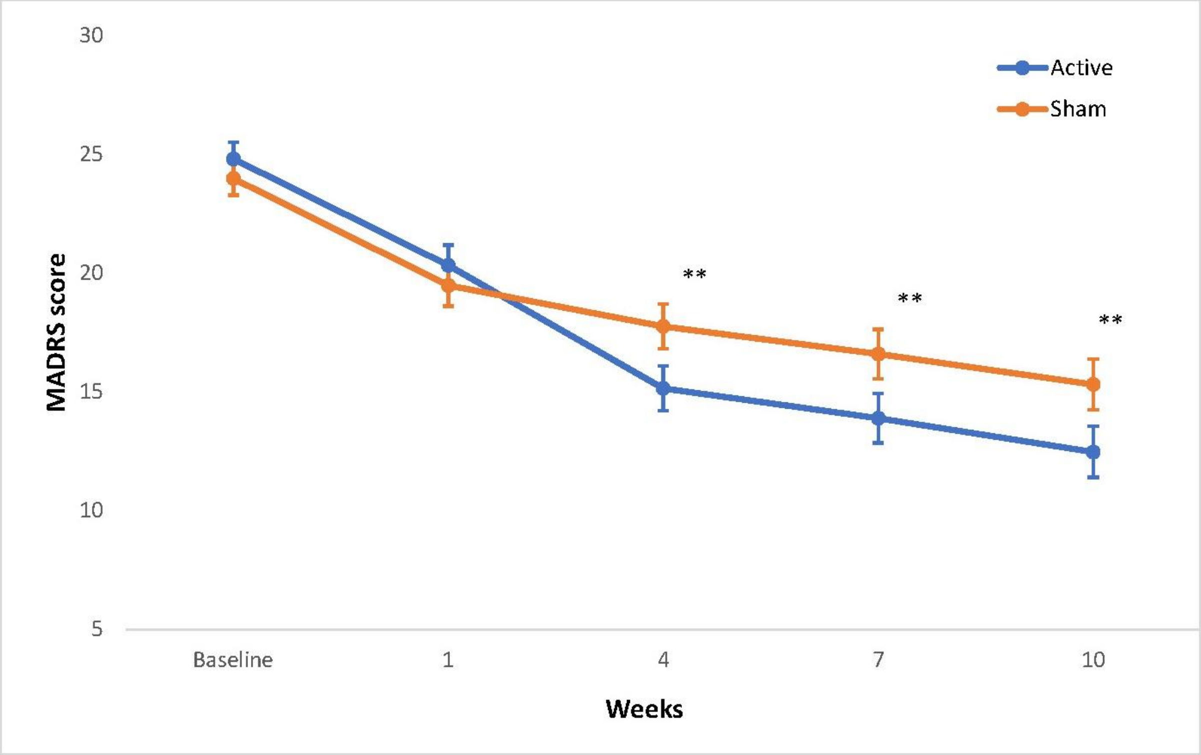

**Supplementary Appendix**

Home-based transcranial direct current stimulation RCT in major depression

### Figure S3. Change in Montgomery-Åsberg Depression Rating Scale-Self report (MADRS-s) ratings over time

Shown are the estimated mean MADRS-s rating scores from baseline to week 10 in the modified intention-to-treat analysis sample (n=173) for the active tDCS and sham tDCS treatment arms. Error bars represent + 1 standard error. MADRS-s scores range from 0 to 60 with higher values indicating more severe depression. A significant improvement was observed in the change in MADRS-s ratings from baseline to week 10 in the active tDCS treatment arm, MADRS-s change 9.9 + 8.94 (SD) (mean week 10 MADRS-s 16.6 + 1.2 (SE)) as compared to sham tDCS treatment arm, MADRS-s change 6.2 + 9.13 (SD) (mean week 10 MADRS-s 19.6 + 1.2 (SE)) (95% CI 0.9 to 6.4, *p* = 0.009). The difference in change scores was also significant at week 4 (*p* = 0.03) with a greater score decrease in the active treatment arm. * = *p* <0.05, ** = *p* <0.01.

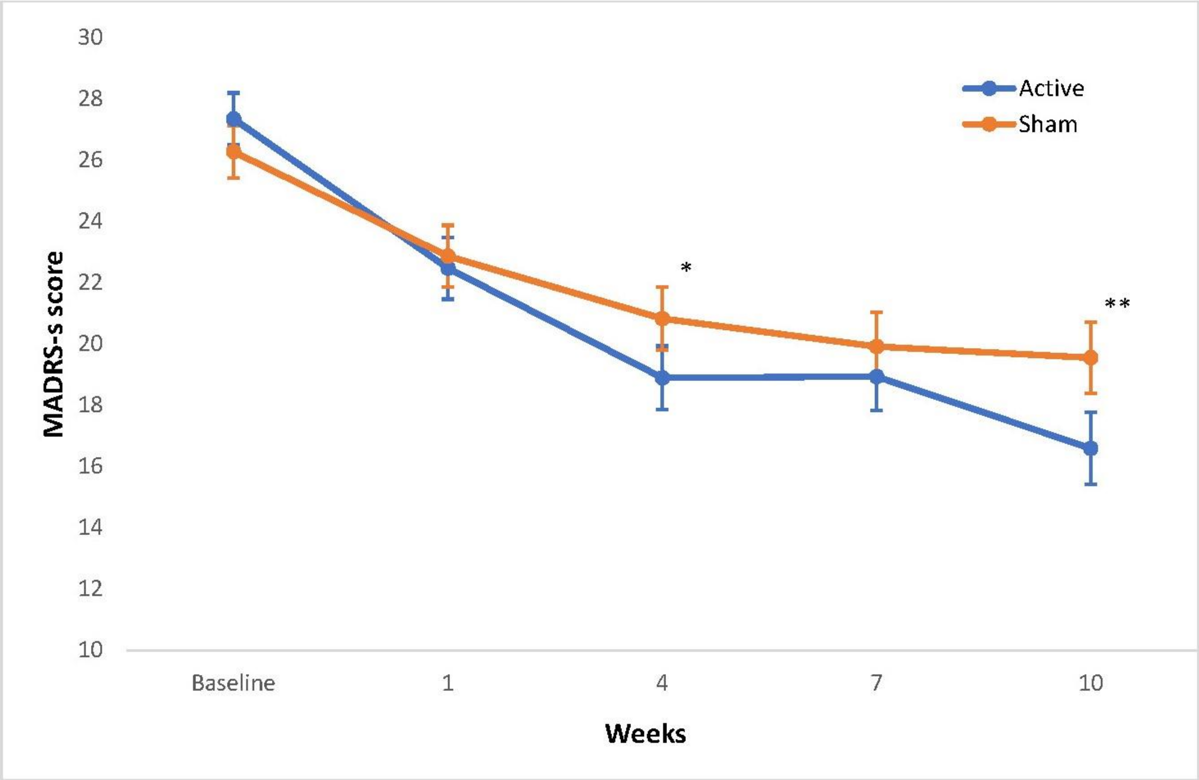

### Table S8. Multiplicity Adjustment

Given that the primary endpoint has been met, the secondary endpoints can be tested. As specified in the protocol, a Hochberg[11,12] approach is used for controlling multiplicity. The 2-sided p-values (and conversion to 1-sided) are provided along with the corresponding Hochberg-adjusted p-values. After correcting for multiplicity, the secondary endpoints Week 10 HDRS-based remission and Week 10 HDRS-based response are statistically significant based on the covariate-adjusted multiple-imputation-based multiplicity-adjusted 1-sided p-values. The Week 10 EQ-5D-3L endpoint is not statistically significant before or after multiplicity correction.

| | 2-sided p-values | 1-sided p-values | 1-sided Hochberg adjusted p-values |
|---|---|---|---|
| Week 10 HDRS response | 0.001 | 0.0005 | 0.0015 |
| Week 10 HDRS remission | 0.004 | 0.0020 | 0.0040 |
| Week 10 EQ-5D-3L | 0.326 | 0.1630 | 0.1630 |

HDRS, Hamilton Depression Rating Scale[3]; EQ-5D-3L, quality of life measure[8–10]. HDRS scores range from 0 to 52 (minimal clinically significant difference = 3 points). Clinical response was defined as a decrease in the score (indicating less depressive severity) of 50% or more from baseline to week 10. Clinical remission was defined as: HDRS score of 7 or less

### Per Protocol (PP) analysis

The Per Protocol (PP) analysis set was comprised of the following:

- Participants in the mITT analysis set
- Participants with device failure within the 10-week follow-up period
- Participants with deviation from the clinical investigation plan is caused by the investigational device or by problems with respect to tolerability

Participant data which were excluded from the PP analysis set are as follows:

- Participants with major protocol violations that would be expected to confound clinical assessment during follow-up as determined by the PIs.
- Participants who took new pharmaceutical substances or treatments during the clinical investigation which are listed as an exclusion criteria.
- Participants who do not meet the inclusion criteria or exclusion criteria.
- Participants who have performed less than 10 sessions (300 minutes) during the first 3 weeks.

Participants in the PP population were analyzed in the group to which they were randomized.

#### Table S9. Per protocol analysis reasons for exclusion

| PP exclusion reason | Number of participants |
|---|---|
| Participants with major protocol violations that would be expected to confound clinical assessment during follow-up as determined by the PIs. | 15 |
| Participants who took new pharmaceutical substances or treatments during the clinical investigation which are listed as an exclusion criteria. | 14 |
| Participants who do not meet the inclusion criteria or exclusion criteria. | 16 |
| Participants who have performed less than 10 sessions (300 minutes) during the first 3 weeks. | 24 |

Participants could meet more than one exclusion criteria. 2 participants completed less than 10 sessions during the first 3 weeks but were included in the PP analysis due to device failure.

### Table S10. Per protocol analysis of changes in depressive severity as measured by HDRS, MADRS and MADRS-s and quality of life as measured by EQ-5D-3L following a 10-week course of active or sham tDCS sessions

| Outcome | Active tDCS (n = 61) | Sham tDCS (n = 70) | Difference or Odds Ratio (95% CI) | Cohen's D | P value |
|---|---|---|---|---|---|
| **Primary Outcome** | | | | | |
| Decrease in HDRS score | 10.4 ± 5.86 | 7.9 ± 5.85 | 2.5 (0.7 to 4.2) | 0.42 | 0.005 |
| **Secondary Outcomes** | | | | | |
| HDRS | | | | | |
| Clinical response | 33 (53.6%) | 19 (26.4%) | 3.23 (1.55 to 6.74) | - | 0.002 |
| Clinical remission | 28 (44.8%) | 15 (20.0%) | 3.25 (1.50 to 7.04) | - | 0.003 |
| MADRS | | | | | |
| Decrease in score | 12.1 ± 8.15 | 8.9 ± 8.15 | 3.2 (0.8 to 5.7) | 0.39 | 0.01 |
| Clinical response | 37 (60.9%) | 23 (31.1%) | 3.44 (1.64 to 7.22) | - | 0.001 |
| Clinical remission | 35 (58.6%) | 23 (30.0%) | 3.31 (1.54 to 7.13) | - | 0.002 |
| MADRS-s | | | | | |
| Decrease in score | 10.8 ± 9.43 | 6.9 ± 9.44 | 3.9 (0.9 to 6.9) | 0.41 | 0.01 |
| Clinical response | 25 (47.3%) | 13 (21.8%) | 3.22 (1.42 to 7.32) | - | 0.005 |
| Clinical remission | 28 (53.5%) | 16 (21.0%) | 4.33 (1.78 to 10.54) | - | 0.001 |
| EQ-5D-3L | | | | | |
| Change in score | 0.07 ± 0.15 | 0.06 ± 0.15 | 0.01 (-0.03 to 0.05) | 0.08 | 0.56 |

HDRS, Hamilton Depression Rating Scale[3]; MADRS, Montgomery-Åsberg Depression Rating Scale[13]; MADRS-s, Montgomery-Åsberg Depression Rating Scale-self report[14]. EQ-5D-3L, quality of life measure[8–10]; CI, confidence interval. Mean values are presented with '±' standard deviation values. HDRS, MADRS, MADRS-s change ratings are the change in total ratings from baseline to week 10. Between-group differences are shown for the changes in scores from baseline to week 10, and odds ratios are shown for the outcomes for clinical response and remission. Percentages for clinical response and remission outcomes are estimated based on odds ratios. HDRS scores range from 0 to 52 (minimal clinically significant difference = 3 points), MADRS scores range from 0 to 60; MADRS-s scores range from 0 to 54, with higher scores indicating more depression. Clinical response was defined as a decrease in the score (indicating less depressive severity) of 50% or more from baseline to week 10. Clinical remission was defined as: HDRS score of 7 or less; MADRS score of 10 or less; MADRS-s score of 12 or less.

### Table S11. Changes in depressive severity as measured by HDRS, MADRS and MADRS-s and quality of life as measured by EQ-5D-3L at 7 weeks

| Outcome | Active (n = 87) | Sham (n = 86) | Difference or Odds Ratio (95% CI) | Cohen's D or NNT | P value |
|---|---|---|---|---|---|
| **Primary Outcome** | | | | | |
| Decrease in HDRS score | 8.1 ± 6.27 | 6.6 ± 6.10 | 1.5 (-0.2 to 3.2) | 0.24 | 0.09 |
| **Secondary Outcomes** | | | | | |
| HDRS | | | | | |
| Clinical response | 35 (47.8%) | 22 (28.8%) | 2.27 (1.14 to 4.51) | 5 | 0.02 |
| Clinical remission | 27 (37.1%) | 15 (20.5%) | 2.30 (1.04 to 5.08) | 6 | 0.04 |
| MADRS | | | | | |
| Decrease in score | 9.9 ± 8.46 | 6.5 ± 8.27 | 3.4 (1.1 to 5.8) | 0.41 | 0.005 |
| Clinical response | 35 (47.1%) | 23 (29.0%) | 2.18 (1.09 to 4.37) | 6 | 0.03 |
| Clinical remission | 34 (47.7%) | 19 (21.8%) | 3.28 (1.49 to 7.20) | 4 | 0.002 |
| MADRS-s | | | | | |
| Decrease in score | 7.6 ± 9.10 | 5.9 ± 9.29 | 1.7 (-0.9 to 4.3) | 0.18 | 0.21 |
| Clinical response | 25 (34.8%) | 15 (23.6%) | 1.72 (0.82 to 3.62) | 9 | 0.16 |
| Clinical remission | 24 (33.2%) | 16 (21.5%) | 1.81 (0.83 to 3.97) | 9 | 0.14 |

HDRS, Hamilton Depression Rating Scale[3]; MADRS, Montgomery-Åsberg Depression Rating Scale[13]; MADRS-s, Montgomery-Åsberg Depression Rating Scale-self report[14]. EQ-5D-3L, quality of life measure[8–10]; CI, confidence interval; NNT, number needed to treat. Mean values are presented with '±' standard deviation values. HDRS, MADRS, MADRS-s change ratings are the change in total ratings from baseline to week 7. Between-group differences are shown for the changes in scores from baseline to week 7, and odds ratios are shown for the outcomes for clinical response and remission. Percentages for clinical response and remission outcomes are estimated based on odds ratios. HDRS scores range from 0 to 52 (minimal clinically significant difference = 3 points), MADRS scores range from 0 to 60; MADRS-s scores range from 0 to 54, with higher scores indicating more depression. Clinical response was defined as a decrease in the score (indicating less depressive severity) of 50% or more from baseline to week 10. Clinical remission was defined as: HDRS score of 7 or less; MADRS score of 10 or less; MADRS-s score of 12 or less.

**Supplementary Appendix**

Home-based transcranial direct current stimulation RCT in major depression

### Subgroup analyses

#### Table S12. Primary and secondary outcomes for participants taking antidepressant medication

| Outcome | Active tDCS (n = 56) | Sham tDCS (n = 53) | Difference or Odds Ratio (95% CI) | P value |
|---|---|---|---|---|
| **Primary Outcome** | | | | |
| Decrease in HDRS score | 10.6 + 6.26 | 7.8 + 5.82 | 2.8 (0. to 5.1) | 0.02 |
| **Secondary Outcomes** | | | | |
| HDRS | | | | |
| Clinical response | 29 (63.2%) | 15 (35.4%) | 3.14 (1.29 to 7.64) | 0.01 |
| Clinical remission | 25 (53.1%) | 11 (25.5%) | 3.33 (1.31 to 8.44) | 0.009 |
| MADRS | | | | |
| Decrease in score | 12.9 + 8.96 | 8.3 + 8.48 | 4.7 (1.2 to 8.1) | 0.009 |
| Clinical response | 33 (70.7%) | 18 (43.8%) | 3.11 (1.24 to 7.77) | 0.02 |
| Clinical remission | 30 (67.1%) | 17 (42.9%) | 2.72 (1.07 to 6.92) | 0.04 |
| MADRS-s | | | | |
| Decrease in score | 9.9 + 9.67 | 6.3 + 9.54 | 3.6 (-0.2 to 7.3) | 0.06 |
| Clinical response | 20 (49.8%) | 10 (26.8%) | 2.73 (1.04 to 7.14) | 0.04 |
| Clinical remission | 22 (57.1%) | 14 (31.2%) | 2.96 (0.97 to 9.01) | 0.05 |
| EQ-5D-3L | | | | |
| Change in score | 0.08 + 0.16 | 0.08 + 0.19 | 0.04 (-0.01 to 0.09) | 0.16 |

HDRS, Hamilton Depression Rating Scale[3]; MADRS, Montgomery-Åsberg Depression Rating Scale[13]; MADRS-s, Montgomery-Åsberg Depression Rating Scale-self report[14]. EQ-5D-3L, quality of life measure[8–10]; CI, confidence interval. Mean values are presented with '±' standard deviation values. HDRS, MADRS, MADRS-s change ratings are the change in total ratings from baseline to week 10. Between-group differences are shown for the changes in scores from baseline to week 10, and odds ratios are shown for the outcomes for clinical response and remission. Percentages for clinical response and remission outcomes are estimated based on odds ratios. HDRS scores range from 0 to 52 (minimal clinically significant difference = 3 points), MADRS scores range from 0 to 60; MADRS-s scores range from 0 to 54, with higher scores indicating more depression. Clinical response was defined as a decrease in the score (indicating less depressive severity) of 50% or more from baseline to week 10. Clinical remission was defined as: HDRS score of 7 or less; MADRS score of 10 or less; MADRS-s score of 12 or less.

**Supplementary Appendix**

Home-based transcranial direct current stimulation RCT in major depression

### Table S13. Primary and secondary outcomes for participants not taking antidepressant medication

| Outcome | Active tDCS (n = 31) | Sham tDCS (n = 33) | Difference or Odds Ratio (95% CI) | P value |
|---|---|---|---|---|
| **Primary Outcome** | | | | |
| Decrease in HDRS score | 7.9 ± 5.96 | 5.2 ± 6.93 | 2.7 (0.1 to 5.5) | 0.06 |
| **Secondary Outcomes** | | | | |
| HDRS | | | | |
| Clinical response | 12 (49.3%) | 6 (10.1%) | 8.73 (1.84 to 41.33) | 0.001 |
| Clinical remission | 9 (36.9%) | 6 (9.4%) | 5.70 (1.19 to 27.20) | 0.02 |
| MADRS | | | | |
| Decrease in score | 8.1 ± 8.76 | 5.7 ± 9.51 | 2.4 (-1.6 to 6.4) | 0.23 |
| Clinical response | 13 (52.3%) | 7 (14.0%) | 6.76 (1.69 to 26.93) | 0.002 |
| Clinical remission | 12 (48.6%) | 8 (11.3%) | 7.41 (1.61 to 34.19) | 0.005 |
| MADRS-s | | | | |
| Decrease in score | 10.2 ± 10.44 | 5.9 ± 10.47 | 4.3 (-1.1 to 9.6) | 0.19 |
| Clinical response | 10 (40.2%) | 4 (16.7%) | 5.05 (1.25 to 20.35) | 0.01 |
| Clinical remission | 10 (51.4%) | 4 (13.2%) | 7.02 (1.54 to 32.00) | 0.005 |
| EQ-5D-3L | | | | |
| Change in score | 0.05 ± 0.14 | 0.05 ± 0.14 | -0.01 (-0.07 to 0.04) | 0.62 |

HDRS, Hamilton Depression Rating Scale[3]; MADRS, Montgomery-Åsberg Depression Rating Scale[13]; MADRS-s, Montgomery-Åsberg Depression Rating Scale-self report[14]. EQ-5D-3L, quality of life measure[8–10]; CI, confidence interval. Mean values are presented with '±' standard deviation values. HDRS, MADRS, MADRS-s change ratings are the change in total ratings from baseline to week 10. Between-group differences are shown for the changes in scores from baseline to week 10, and odds ratios are shown for the outcomes for clinical response and remission. Percentages for clinical response and remission outcomes are estimated based on odds ratios. HDRS scores range from 0 to 52 (minimal clinically significant difference = 3 points), MADRS scores range from 0 to 60; MADRS-s scores range from 0 to 54, with higher scores indicating more depression. Clinical response was defined as a decrease in the score (indicating less depressive severity) of 50% or more from baseline to week 10. Clinical remission was defined as: HDRS score of 7 or less; MADRS score of 10 or less; MADRS-s score of 12 or less.

**Supplementary Appendix**

Home-based transcranial direct current stimulation RCT in major depression

### Table S14. Young Mania Rating Scale change scores from baseline to week 10 and scores at all time points

| Outcome | Active tDCS (n = 87) | Sham tDCS (n = 86) | Difference or Odds Ratio (95% CI) | P value |
|---|---|---|---|---|
| **YMRS** | | | | |
| Decrease in score week 10 | 0.8 ± 2.1 | 0.2 ± 1.9 | 0.7 (0.0 to 1.3) | 0.05 |
| Baseline score | 2.1 ± 1.7 | 1.9 ± 1.6 | 0.2 (-0.3 to 0.7) | 0.52 |
| Week 1 score | 1.6 ± 1.7 | 1.9 ± 1.6 | -0.2 (-0.8 to 0.3) | 0.34 |
| Week 4 score | 1.5 ± 1.6 | 1.7 ± 1.4 | -0.2 (-0.7 to 0.3) | 0.46 |
| Week 7 score | 1.6 ± 1.7 | 1.5 ± 1.6 | 0.0 (-0.5 to 0.6) | 0.92 |
| Week 10 score | 1.3 ± 1.4 | 1.8 ± 1.7 | -0.6 (-1.1 to -0.1) | 0.03 |

YMRS, Young Mania Rating Scale[4]; tDCS, transcranial direct current stimulation. Plus-minus values are mean ± standard deviation values.

**Supplementary Appendix**

Home-based transcranial direct current stimulation RCT in major depression

### Table S15. Hamilton Anxiety Rating Scale (HAMA) change scores from baseline to week 10 and scores at all time points.

| Outcome | Active tDCS (n = 87) | Sham tDCS (n = 86) | Difference or Odds Ratio (95% CI) | P value |
|---|---|---|---|---|
| **HAMA** | | | | |
| Decrease in score | 6.6 ± 6.1 | 4.9 ± 5.9 | 1.7 (0.2 to 3.7) | 0.08 |
| Baseline score | 15.4 ± 4.6 | 14.3 ± 4.6 | 1.1 (-0.2 to 2.5) | 0.11 |
| Week 10 score | 8.2 ± 5.7 | 9.3 ± 4.9 | -1.1 (-2.8 to 0.7) | 0.23 |

HAMA, Hamilton Anxiety Rating Scale;[15] tDCS, transcranial direct current stimulation. Plus-minus values are mean ± standard deviation values.

### Figure S4. Correlation between change from baseline to 10 weeks in MADRS and MADRS-s

The correlation between change from baseline to 10 weeks in MADRS and MADRS-s is an exploratory endpoint in the trial. There are 173 subjects randomized in the trial. Of the 173 subjects, 46 subjects had missing MADRS or MADRS-s data at week 10. Of the 46 subjects, 22 of the subjects had early withdrawal from the trial, 3 subjects were missing MADRS data at week 10 and 21 subjects were missing MADRS-s data at week 10. The remaining 127 subjects were included in the correlation analysis. Percent of sessions with stimulation was calculated as the observed stimulation time between weeks 1 through 10 divided by the total number of expected minutes based on protocol recommendations (1080 = 30*5*3+30*3*7). The scatter plot shows the change from baseline to 10 weeks in MADRS-s score (madrs_s_score_chg) versus the change from baseline to 10 weeks in MADRS score (madrs_score_chg).

With the 127 observations, there was an observed correlation of 0.49 between the percent of sessions with stimulation and the change from baseline to 10 weeks in MADRS score (Pearson correlation $p < 0.0001$). Therefore, there is a statistically significant correlation between the percent of sessions with stimulation and the change from baseline to 10 weeks in MADRS score. The correlation between the two measures would be considered moderate. From Fantino et al., the investigators observed a Pearson's correlation between MADRS and MADRS-s of r=0.54 in subjects with major depressive disorder. The observed correlation of 0.49 in the trial is well aligned with the results from the Fantino and Moore (2009) study.[16]

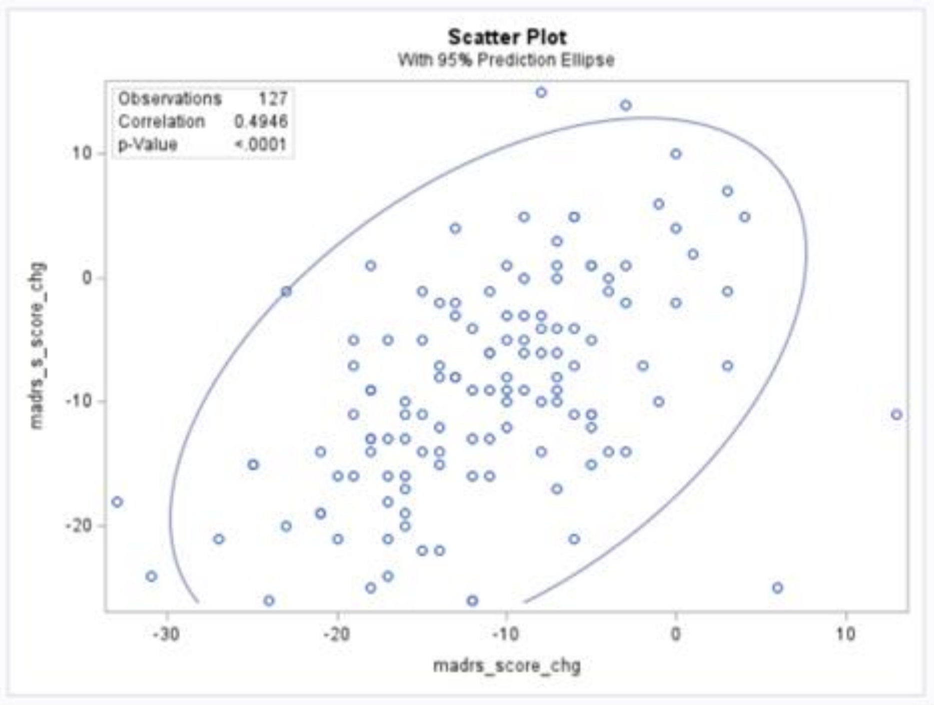

### Figure S5. Correlation between change from baseline to 10 weeks in HDRS and percent of sessions with stimulation in the active group

The correlation between change from baseline to 10 weeks in HDRS and percent of sessions with stimulation in the active group is an exploratory endpoint in the trial. There are 86 subjects randomized to the active group in the trial. Of the 86 subjects, 13 subjects had missing HDRS data at week 10. All 13 of the subjects had early withdrawal from the trial. The remaining 73 subjects were included in the correlation analysis. Percent of sessions with stimulation was calculated as the observed stimulation time between weeks 1 through 10 divided by the total number of expected minutes based on protocol recommendations (1080 = 30*5*3+30*3*7). The scatter plot shows the graphical representation of the percent of sessions with stimulation (pct_sessions) versus the change from baseline to 10 weeks in HDRS score (hdrs_17_score_chg).

With the 73 observations, there was an observed correlation of -0.093 between the percent of sessions with stimulation and the change from baseline to 10 weeks in HDRS score (Pearson correlation p=0.4362). Therefore, there is not a statistically significant correlation between the percent of sessions with stimulation and the change from baseline to 10 weeks in HDRS score. The correlation between the two measures would be considered weak.

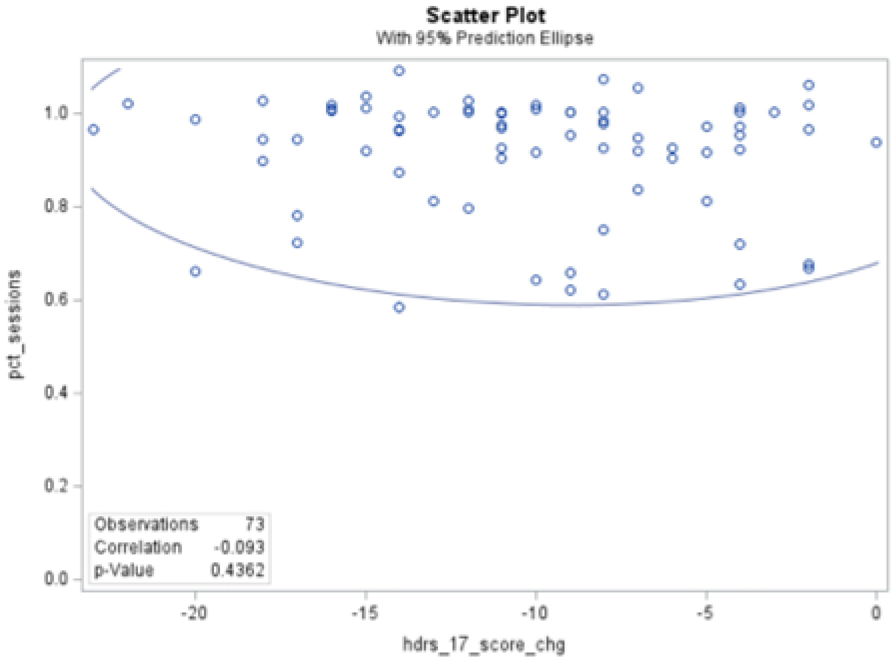

### Figure S6. Correlation between change from baseline to 10 weeks in MADRS and percent of sessions with stimulation in the active group

The correlation between change from baseline to 10 weeks in MADRS and percent of sessions with stimulation in the active group is an exploratory endpoint in the trial. There are 86 subjects randomized to the active group in the trial. Of the 86 subjects, 13 subjects had missing MADRS data at week 10. All 13 of the subjects had early withdrawal from the trial. The remaining 73 subjects were included in the correlation analysis. Percent of sessions with stimulation was calculated as the observed stimulation time between weeks 1 through 10 divided by the total number of expected minutes based on protocol recommendations (1080 = 30*5*3+30*3*7). The scatter plot shows the graphical representation of the percent of sessions with stimulation (pct_sessions) versus the change from baseline to 10 weeks in MADRS score (madrs_score_chg).

With the 73 observations, there was an observed correlation of -0.06 between the percent of sessions with stimulation and the change from baseline to 10 weeks in MADRS score (Pearson correlation p=0.6155). Therefore, there is not a statistically significant correlation between the percent of sessions with stimulation and the change from baseline to 10 weeks in MADRS score. The correlation between the two measures would be considered weak.

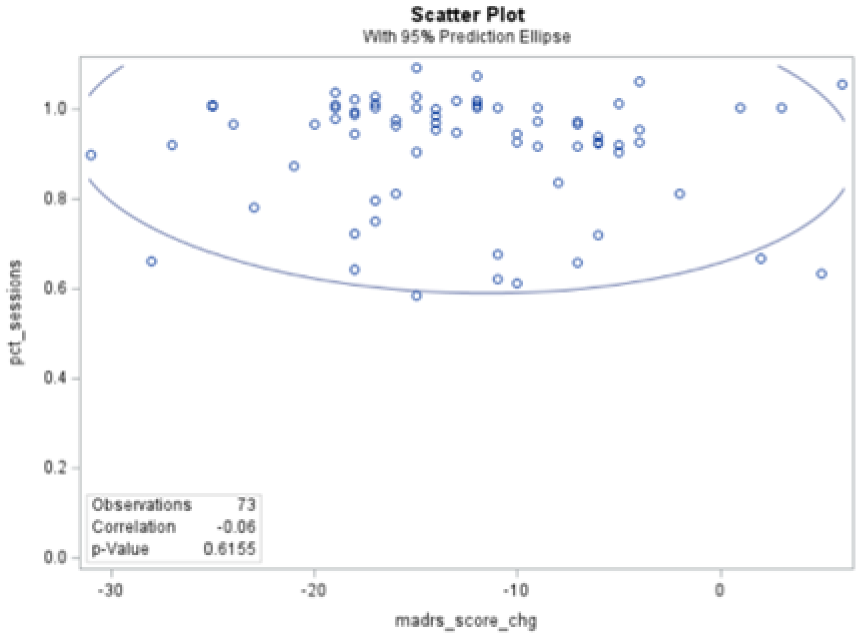

**Supplementary Appendix**

Home-based transcranial direct current stimulation RCT in major depression

### Neuropsychological measures

#### Table S16. Neuropsychological measure: verbal learning (Rey Auditory Verbal Learning Test (RAVLT) and information processing speed (Symbol-Digit Modalities Test (SDMT) change scores from baseline to week 10

| Outcome | Active tDCS (n = 87) | Sham tDCS (n = 86) | Difference or Odds Ratio (95% CI) | P value |
|---|---|---|---|---|
| **RAVLT** | | | | |
| Change in score | 3.1 ± 9.9 | 3.6 ± 9.1 | -0.5 (-3.6 to 2.5) | 0.73 |
| **SDMT** | | | | |
| Change in score | 2.5 ± 10.4 | 2.7 ± 7.5 | -0.2 (-3.3 to 2.8) | 0.89 |

SDMT, Symbol Digit Modalities Test[17]; RAVLT, Rey Auditory Verbal Learning Test[18]; tDCS, transcranial direct current stimulation. Plus-minus values are mean ± standard deviation values. RAVLT scores range from 0 to 75. SDMT scores range from 0 to 110.

#### Figure S7. Rey Auditory Verbal Learning Test (RAVLT) scores from baseline to week 10

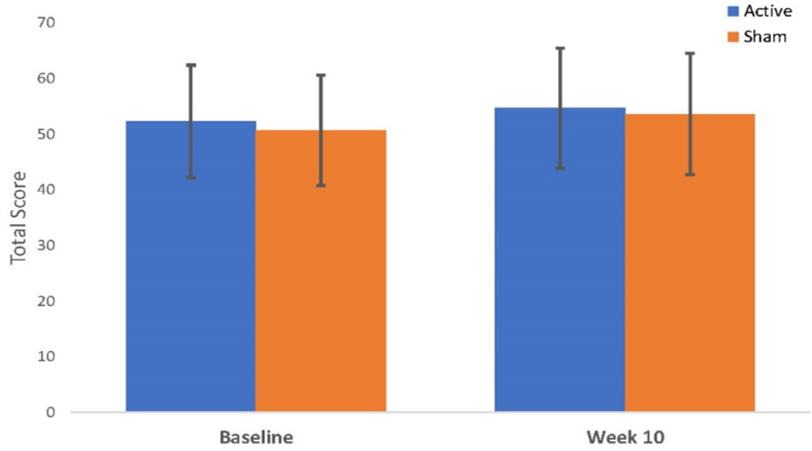

Error bars represent standard deviation.

#### Figure S8. Symbol-Digit Modalities Test (SDMT) scores from baseline to week 10

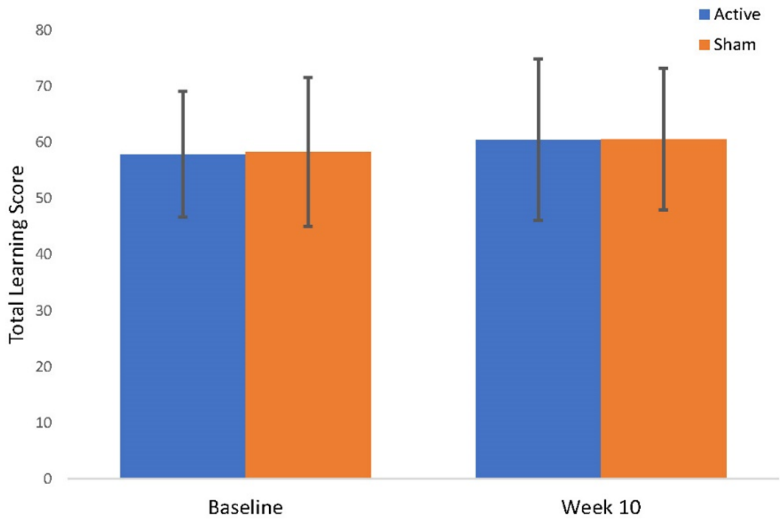

Error bars represent standard deviation.

### Table S17. Treatment acceptability questionnaire (TAQ) and responses at baseline (n = 174 (active = 87, sham = 87)), at the end of treatment (n = 151 (active = 75, sham = 76). Italics represent post-treatment phrasing

| | | | | Likert Ratings | | | | | | | | | | | | | |
|---|---|---|---|---|---|---|---|---|---|---|---|---|---|---|---|---|---|
| | Median (IQR) | | | 1 | | 2 | | 3 | | 4 | | 5 | | 6 | | 7 | |
| Question | Active tDCS | Sham tDCS | P Value | Active tDCS | Sham tDCS | Active tDCS | Sham tDCS | Active tDCS | Sham tDCS | Active tDCS | Sham tDCS | Active tDCS | Sham tDCS | Active tDCS | Sham tDCS | Active tDCS | Sham tDCS |
| **How acceptable do *(did)* you consider the tDCS sessions to be?** | | | | **Very unacceptable** | | **Quite unacceptable** | | **Unacceptable** | | **Neither** | | **Acceptable** | | **Quite acceptable** | | **Very acceptable** | |
| Baseline | 7 (1) | 6 (2) | 0.27 | 1 (1%) | 4 (5%) | 1 (1%) | 2 (2%) | 0 (0%) | 0 (0%) | 11 (13%) | 14 (16%) | 4 (5%) | 5 (6%) | 25 (29%) | 21 (24%) | 45 (52%) | 41 (47%) |
| After 10 weeks treatment | 7 (1) | 7 (1) | 0.83 | 0 (0%) | 0 (0%) | 0 (0%) | 4 (5%) | 0 (0%) | 2 (3%) | 2 (3%) | 3 (4%) | 3 (4%) | 4 (5%) | 24 (32%) | 14 (18%) | 46 (61%) | 49 (64%) |
| **How helpful do you think the tDCS sessions may be *(were)* for improving your depressive symptoms?** | | | | **Very unhelpful** | | **Quite unhelpful** | | **Bit unhelpful** | | **Neither** | | **Bit helpful** | | **Quite helpful** | | **Very helpful** | |
| Baseline | 6 (1) | 6 (1) | 0.94 | 0 (0%) | 0 (0%) | 1 (1%) | 1 (1%) | 2 (2%) | 1 (1%) | 8 (9%) | 8 (9%) | 26 (30%) | 30 (34%) | 36 (41%) | 31 (36%) | 14 (16%) | 16 (18%) |
| After 10 weeks treatment | 6 (2) | 5 (2) | 0.002 | 0 (0%) | 3 (4%) | 1 (1%) | 3 (4%) | 1 (1%) | 1 (1%) | 9 (12%) | 19 (25%) | 21 (28%) | 20 (26%) | 21 (28%) | 20 (26%) | 22 (29%) | 10 (13%) |
| **How likely do you think that there will be negative side effects from the tDCS sessions? */ How were you bothered by any negative side effects from the tDCS sessions?*** | | | | **Very unlikely/ Very much unaffected** | | **Quite unlikely/ Quite unaffected** | | **Bit unlikely/ Bit unaffected** | | **Neither** | | **Bit likely/ Bit affected** | | **Quite likely/ Quite affected** | | **Very likely/ Very affected** | |
| Baseline | 3 (2) | 2 (2) | 0.62 | 17 (20%) | 15 (17%) | 26 (30%) | 31 (36%) | 14 (16%) | 15 (17%) | 17 (20%) | 18 (21%) | 11 (13%) | 7 (8%) | 1 (1%) | 1 (1%) | 1 (1%) | 0 (0%) |
| After 10 weeks treatment | 2 (2) | 1 (1) | 0.21 | 29 (39%) | 39 (52%) | 26 (35%) | 19 (25%) | 4 (5%) | 2 (3%) | 1 (1%) | 4 (5%) | 14 (19%) | 10 (13%) | 1 (1%) | 1 (1%) | 0 (0%) | 1 (1%) |
| **How ethical do you think the tDCS sessions are?** | | | | **Very unethical** | | **Quite unethical** | | **Bit unethical** | | **Neither** | | **Bit ethical** | | **Quite ethical** | | **Very ethical** | |
| Baseline | 7 (1) | 7 (1) | 0.03 | 0 (0%) | 1 (1%) | 0 (0%) | 4 (5%) | 1 (1%) | 3 (3%) | 6 (7%) | 5 (6%) | 0 (0%) | 0 (0%) | 20 (23%) | 27 (31%) | 60 (69%) | 47 (54%) |
| After 10 weeks treatment | 7 (0) | 7 (0) | 0.02 | 0 (0%) | 0 (0%) | 0 (0%) | 0 (0%) | 0 (0%) | 0 (0%) | 2 (3%) | 5 (7%) | 0 (0%) | 1 (1%) | 4 (5%) | 10 (13%) | 69 (92%) | 60 (79%) |
| **How much effort do you think you need *(did you need)* to put in for the tDCS sessions?** | | | | **Very much more than usual** | | **Some more than usual** | | **Little bit more than usual** | | **Same as usual** | | **Bit less than usual** | | **Quite less than usual** | | **Very much less than usual** | |
| Baseline | 3 (2) | 3 (2) | 0.94 | 4 (5%) | 3 (3%) | 22 (25%) | 22 (25%) | 27 (31%) | 29 (33%) | 25 (29%) | 26 (30%) | 4 (5%) | 3 (3%) | 3 (3%) | 2 (2%) | 2 (2%) | 2 (2%) |
| After 10 weeks treatment | 4 (3) | 3 (1) | 0.06 | 0 (0%) | 2 (3%) | 5 (7%) | 7 (9%) | 27 (36%) | 33 (43%) | 19 (25%) | 18 (24%) | 2 (3%) | 3 (4%) | 13 (17%) | 8 (11%) | 9 (12%) | 5 (7%) |
| **Would you recommend the tDCS sessions to others?** | | | | **Would very strongly not recommend** | | **Would strongly not recommend** | | **Would not recommend** | | **Would not for or against** | | **Would recommend** | | **Would strongly recommend** | | **Would very strongly recommend** | |
| After 10 weeks treatment | 6 (2) | 5 (1.5) | 0.15 | 1 (1%) | 0 (0%) | 1 (1%) | 1 (1%) | 1 (1%) | 1 (1%) | 6 (8%) | 16 (21%) | 22 (30%) | 20 (27%) | 19 (25%) | 18 (24%) | 25 (33%) | 20 (26%) |

tDCS, transcranial direct current stimulation; IQR, inter-quartile range. Active and sham scores were compared using Mann-Whitney U tests.

A Mann-Whitney U test was conducted on each question at baseline and at week 10 to compare the distribution of scores between the active and sham treatment groups.

Participants were asked “How acceptable do you consider the tDCS sessions to be?” at baseline, and “How acceptable did you consider the tDCS sessions to be?” at week 10. At baseline, acceptability was endorsed as being “very acceptable” by the active tDCS group and “quite acceptable” by the sham group and there was no significant difference between the groups ($p$ =0.27). At week 10, both groups rated tDCS as “very acceptable” with no significant difference between group ($p$ = 0.83).

At baseline participants were asked “How helpful do you think the tDCS sessions may be for improving your depressive symptoms?”. Both groups thought that the treatment would be “quite helpful” at baseline with no significant difference in the distribution of scores ($p$ =0.94). When asked at week 10 “How helpful do you think the tDCS sessions were for improving your depressive symptoms?”, the active group thought that the treatment was more helpful than the sham group at week 10, rating the treatment as “quite helpful”, with the sham group rating that the treatment was “a bit helpful”. At baseline, there was a significant difference in the distribution of scores ($p$ = 0.002), with more participants in the sham group rating the treatment and “neither helpful or unhelpful” or one of the three “unhelpful” options.

Ethicality remained high at “very ethical” for both groups at both time points, however, the distribution of scores across groups was significantly different at both time points. Participants were asked “How ethical do you think the tDCS sessions are?” at both time points. At baseline participants used the full range of scores in the sham group and there was a significant difference in the distribution of scores between the groups ($p$ = 0.03). After receiving the treatment, no participants in either group rated the treatment as “unethical”, and 92% of participants in the active group rated the treatment at “very ethical”, the distribution of scores remained significantly different between groups ($p$ = 0.02).

At baseline participants were asked “How much effort do you think you need to put in for the tDCS sessions?” and both groups median response was a “little bit more than usual” with no significant difference between groups ($p$ = 0.94). This rating remained consistent for the sham group at week 10 when asked “How much effort did you need to put in for the tDCS sessions?”. The active group rated “some more effort than usual” at week 10 but there was no significant difference between the groups ($p$ = 0.06).

Participants were asked “How likely do you think that there will be negative side effects from the tDCS sessions?” at baseline. The active group thought it was “a bit unlikely” that they would be bothered by any side effects from the tDCS and the sham group thought it was “quite unlikely”, there was no significant difference between the groups ($p$ = 0.62) After treatment at week 10, participants were asked “How were you bothered by any negative side effects from the tDCS sessions?”. Both groups median scores reduced by one point with the active group reporting that they were “quite unaffected” by any negative side effects and the sham group reported that they were “very much unaffected” by the side effects. The difference between groups was not significant ($p$ = 0.21).

After receiving the treatment, both groups were asked “Would you recommend the tDCS sessions to others?” at week 10. The active group “would strongly recommend” the tDCS sessions and the sham group “would recommend” the tDCS sessions. The distribution of scores between the groups was not significant ($p$ = 0.15)

.

### Figure S9. Percentage of participants in active and sham groups who endorsed each response in the acceptability questionnaire at baseline and week 10

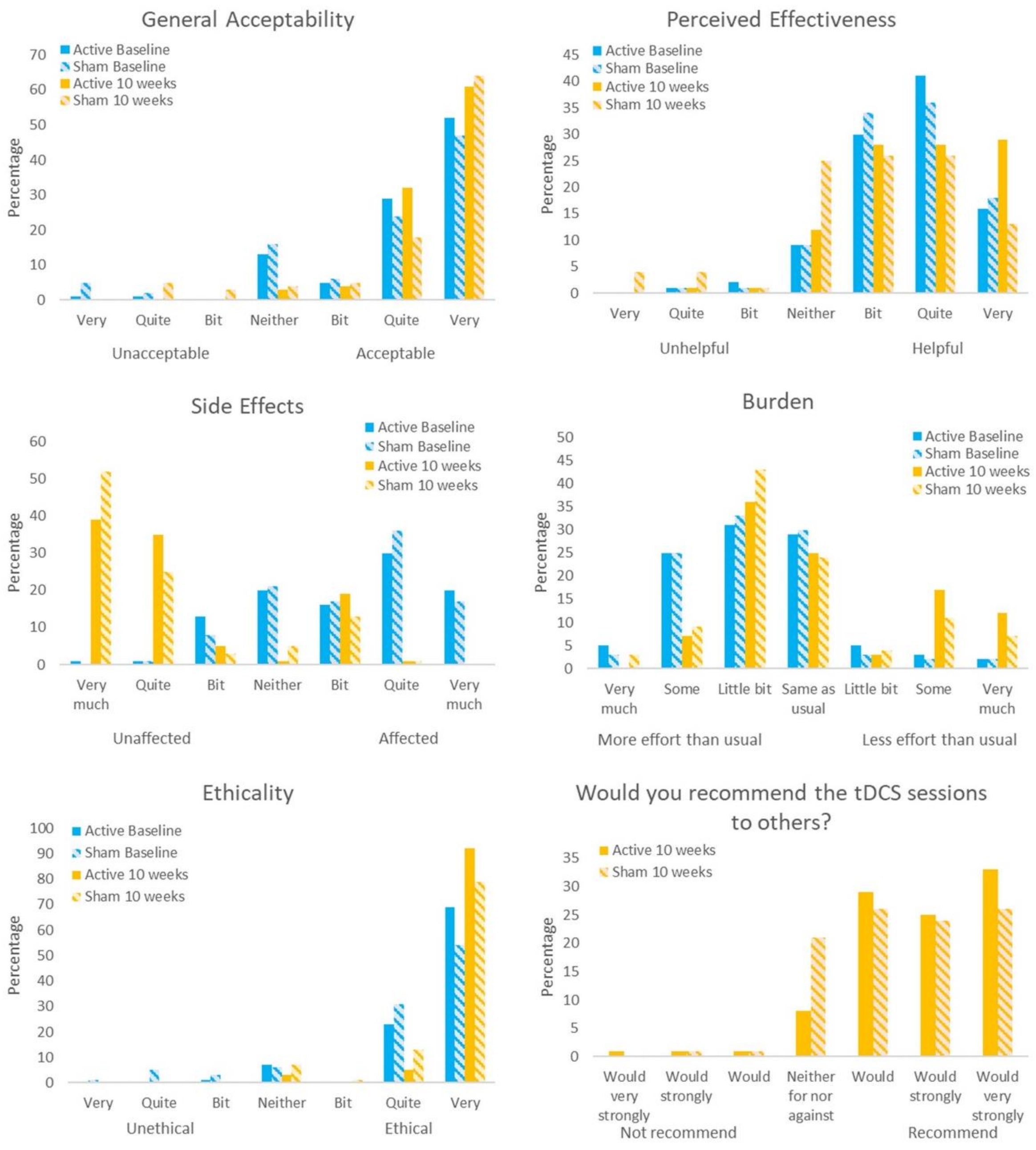

### Table S18. The number of participants who showed a clinically meaningful improvement at each time point (indicated by a HDRS scores of -3 from baseline)

The percentage of subjects for each arm that reached -3 points or more improvement at each time point from week 1 to week 10. -3 points is the smallest change in measurement on the HDRS that signifies a clinically meaningful improvement in and individual patient.

| Outcome | Active tDCS (n = 87) | Sham tDCS (n = 86) | Difference % (95% CI) | P value |
|---|---|---|---|---|
| **Clinically meaningful improvement** | | | | |
| Week 1 | 49 (59.0%) | 56 (66.7%) | -7.6 (-22.2 to 7.0) | 0.308 |
| Week 4 | 62 (80.5%) | 59 (77.6%) | 2.9 (-10.0 to 15.8) | 0.661 |
| Week 7 | 67 (89.3%) | 62 (82.7%) | 6.7 (-4.4 to 17.7) | 0.239 |
| Week 10 | 68 (91.9%) | 65 (86.7%) | 5.2 (-4.7 to 15.1) | 0.303 |

tDCS, transcranial direct current stimulation. CI, confidence interval.

**Supplementary Appendix**

Home-based transcranial direct current stimulation RCT in major depression

### Cumulative Proportion of Responders Analysis (CPRA) - Exploratory Analyses

#### HDRS

A Cumulative Proportion of Responders Analysis (CPRA) was performed per the Statistical Analysis Plan (SAP). The following figure shows:

- X-axis: The change from baseline to Week 10 in HDRS
- Y-axis: The percentage (%) of subjects experiencing a reduction greater than ‘k’ from the baseline in HDRS.
- The blue line is the Active group, and the red line is Sham.

Figure S10 shows the percentage of subjects experiencing a response as a function of the change in the HDRS from baseline. For this endpoint, the “more negative” primary endpoint change from baseline indicates a better outcome subject. The observed cumulative percent response in the treatment group is more negative than the control group for all observed HDRS from baseline.

The difference can be further summarized by the table above with cumulative response group difference by specified cut points. The difference between the groups is depicted by an initial separation of the curves with a maximum at 10 point difference in HDRS from baseline, which resulted in a 22.0% group difference in response.

Please note, for this analysis, the observed values are used and not imputed as in the primary endpoint analysis. Therefore, there may be very slight differences between the results in this analyses and other reported values. This was done for analytical simplicity and has no impact on the interpretation of the results.

In general, the cumulative response shows a better response to the change in HDRS for subjects in the treatment arm versus the control arm, regardless of the MCID (Minimal Clinically Important Difference) chosen.

#### Figure S10. HDRS cumulative proportion of responders analysis

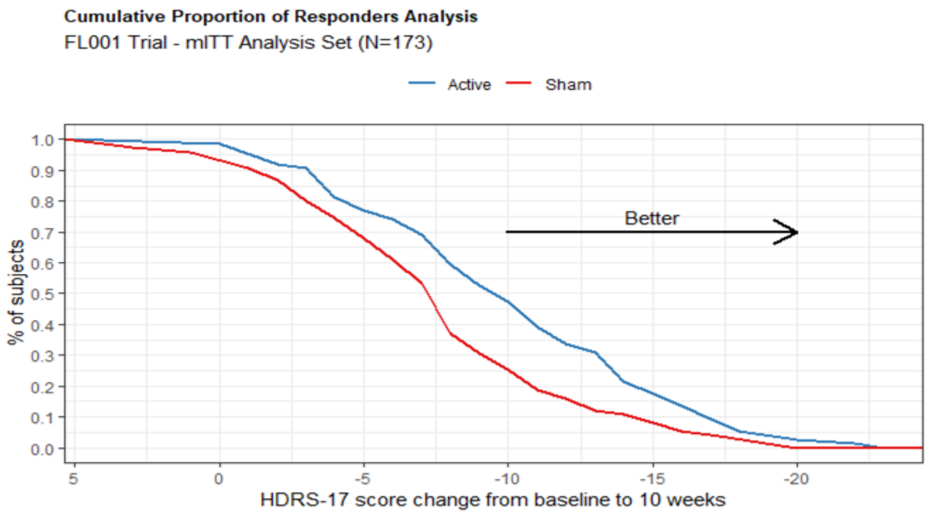

| *HDRS-17 Score Change* | -3 | -5 | -10 | -15 | -20 |
|---|---|---|---|---|---|
| *Group Difference* | 10.6% | 9.0% | 22.0% | 9.6% | 2.7% |

**Supplementary Appendix**

Home-based transcranial direct current stimulation RCT in major depression

### MADRS

A Cumulative Proportion of Responders Analysis (CPRA) was performed per the Statistical Analysis Plan (SAP). The following figure shows:

- X-axis: The change from baseline to Week 10 in MADRS
- Y-axis: The percentage (%) of subjects experiencing a reduction greater than 'k' from the baseline in MADRS.
- The blue line is the Active group, and the red line is Sham.

Figure S11 shows the percentage of subjects experiencing a response as a function of the change in the MADRS from baseline. For this endpoint, the "more negative" primary endpoint change from baseline indicates a better outcome subject. The observed cumulative percent response in the treatment group is more negative than the control group for all observed MADRS from baseline.

The difference can be further summarized by the table above with cumulative response group difference by specified cut points. The difference between the groups is depicted by an initial separation of the curves with a maximum at 10 point difference in MADRS from baseline, which resulted in a 24.9% group difference in response.

In general, the cumulative response shows a better response to the change in MADRS for subjects in the treatment arm versus the control arm, regardless of the MCID chosen.

**Figure S11. MADRS cumulative proportion of responders analysis**

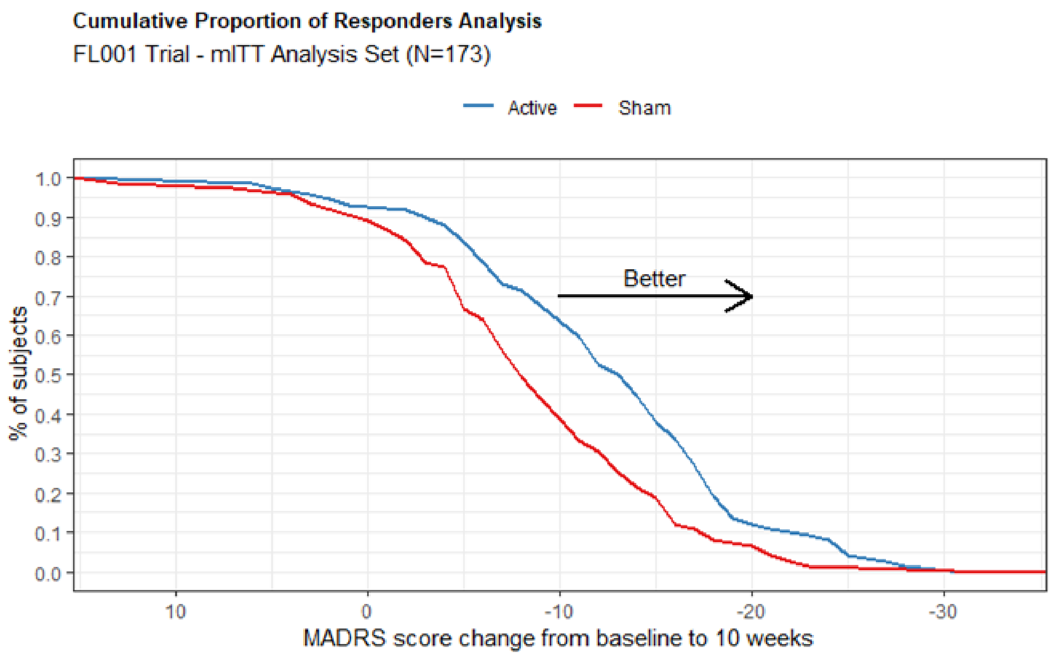

| *MADRS Score Change* | -5 | -10 | -15 | -20 |
|---|---|---|---|---|
| *Group Difference* | 17.1% | 24.9% | 19.2% | 5.5% |

### MADRS-s

A Cumulative Proportion of Responders Analysis (CPRA) was performed per the Statistical Analysis Plan (SAP). The following figure shows:

- X-axis: The change from baseline to Week 10 in MADRS-s
- Y-axis: The percentage (%) of subjects experiencing a reduction greater than ‘k’ from the baseline in MADRS-s.
- The blue line is the Active group, and the red line is Sham.

Figure S12 shows the percentage of subjects experiencing a response as a function of the change in the MADRS-s from baseline. For this endpoint, the “more negative” primary endpoint change from baseline indicates a better outcome subject. The observed cumulative percent response in the treatment group is more negative than the control group for all observed MADRS-s from baseline.

The difference can be further summarized by the table above with cumulative response group difference by specified cut points. The difference between the groups is depicted by an initial separation of the curves with a maximum at 10 point difference in MADRS-s from baseline, which resulted in a 23.0% group difference in response.

In general, the cumulative response shows a better response to the change in MADRS-s for subjects in the treatment arm versus the control arm, regardless of the MCID chosen.

**Figure S12. MADRS-s cumulative proportion of responders analysis**

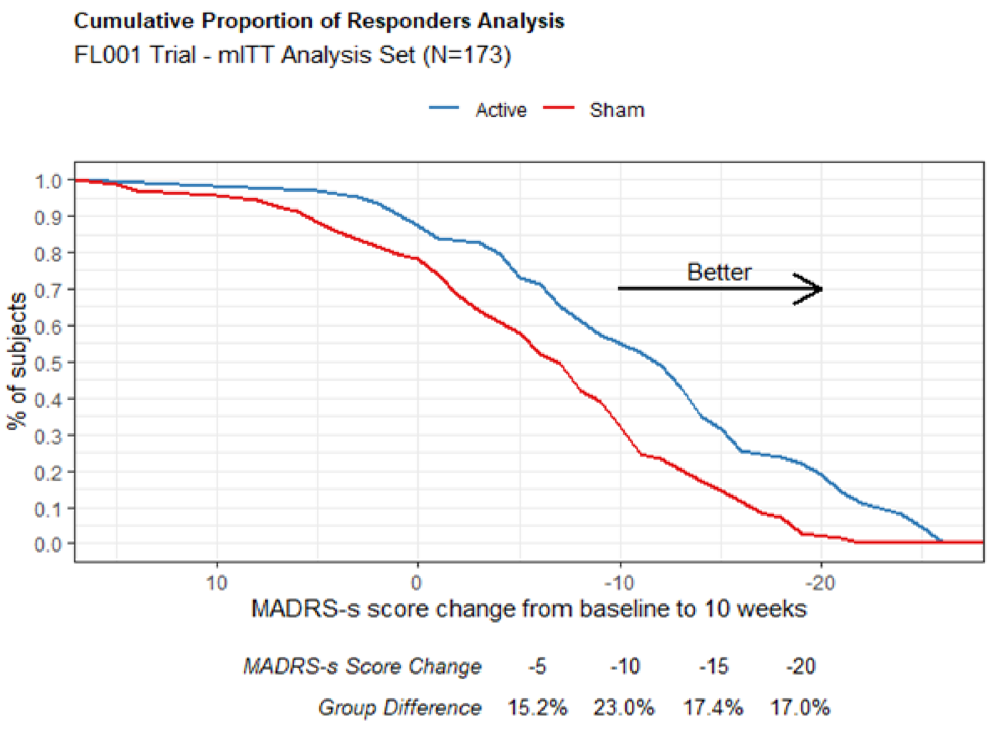

**Supplementary Appendix**

Home-based transcranial direct current stimulation RCT in major depression

### Table S19. All unanticipated device related adverse events

| | Active (N= 87) | | | Sham (N= 86) | | | Group Difference | | | |
|---|---|---|---|---|---|---|---|---|---|---|
| | Events | Subjs | % | Events | Subjs | % | Δ | LB | UB | P value |
| All Events | 48 | 30 | 34.5% | 27 | 21 | 24.4% | 10.1% | -3.7% | 23.8% | 0.18 |
| Tinnitus | 2 | 2 | 2.3% | 2 | 2 | 2.3% | 0.0% | -6.2% | 6.0% | 0.99 |
| Photopsia | 2 | 2 | 2.3% | 1 | 1 | 1.2% | 1.1% | -4.5% | 7.0% | 0.99 |
| Vision blurred | 2 | 1 | 1.1% | 0 | 0 | 0.0% | 1.1% | -3.3% | 6.4% | 0.99 |
| Nausea | 3 | 2 | 2.3% | 1 | 1 | 1.2% | 1.1% | -4.5% | 7.0% | 0.99 |
| Fatigue | 1 | 1 | 1.1% | 0 | 0 | 0.0% | 1.1% | -3.3% | 6.4% | 0.99 |
| Malaise | 1 | 1 | 1.1% | 0 | 0 | 0.0% | 1.1% | -3.3% | 6.4% | 0.99 |
| Pain | 1 | 1 | 1.1% | 2 | 2 | 2.3% | -1.2% | -7.2% | 4.4% | 0.62 |
| Nasopharyngitis | 1 | 1 | 1.1% | 0 | 0 | 0.0% | 1.1% | -3.3% | 6.4% | 0.99 |
| Sinusitis | 0 | 0 | 0.0% | 1 | 1 | 1.2% | -1.2% | -6.5% | 3.3% | 0.50 |
| Burns first degree | 2 | 2 | 2.3% | 0 | 0 | 0.0% | 2.3% | -2.2% | 8.1% | 0.50 |
| Burning sensation | 1 | 1 | 1.1% | 0 | 0 | 0.0% | 1.1% | -3.3% | 6.4% | 0.99 |
| Dizziness | 4 | 3 | 3.4% | 2 | 2 | 2.3% | 1.1% | -5.2% | 8.0% | 0.99 |
| Dysgeusia | 0 | 0 | 0.0% | 1 | 1 | 1.2% | -1.2% | -6.5% | 3.3% | 0.50 |
| Headache | 0 | 0 | 0.0% | 1 | 1 | 1.2% | -1.2% | -6.5% | 3.3% | 0.50 |
| Hyperaesthesia | 0 | 0 | 0.0% | 1 | 1 | 1.2% | -1.2% | -6.5% | 3.3% | 0.49 |
| Migraine | 1 | 1 | 1.1% | 0 | 0 | 0.0% | 1.1% | -3.3% | 6.4% | 0.99 |
| Ophthalmic migraine | 0 | 0 | 0.0% | 1 | 1 | 1.2% | -1.2% | -6.5% | 3.3% | 0.50 |
| Paraesthesia | 2 | 2 | 2.3% | 3 | 3 | 3.5% | -1.2% | -8.0% | 5.1% | 0.68 |
| Aggression | 0 | 0 | 0.0% | 1 | 1 | 1.2% | -1.2% | -6.5% | 3.3% | 0.50 |
| Anxiety | 1 | 1 | 1.1% | 1 | 1 | 1.2% | 0.0% | -5.5% | 5.3% | 0.99 |
| Depressed mood | 1 | 1 | 1.1% | 0 | 0 | 0.0% | 1.1% | -3.3% | 6.4% | 0.99 |
| Insomnia | 1 | 1 | 1.1% | 2 | 2 | 2.3% | -1.2% | -7.2% | 4.4% | 0.62 |
| Panic attack | 1 | 1 | 1.1% | 0 | 0 | 0.0% | 1.1% | -3.3% | 6.4% | 0.99 |
| Tension | 1 | 1 | 1.1% | 0 | 0 | 0.0% | 1.1% | -3.3% | 6.4% | 0.99 |
| Acne | 1 | 1 | 1.1% | 1 | 1 | 1.2% | 0.0% | -5.5% | 5.3% | 0.99 |
| Alopecia | 1 | 1 | 1.1% | 0 | 0 | 0.0% | 1.1% | -3.3% | 6.4% | 0.99 |
| Dry skin | 9 | 9 | 10.3% | 4 | 4 | 4.7% | 5.7% | -2.5% | 14.8% | 0.25 |
| Pruritus | 0 | 0 | 0.0% | 1 | 1 | 1.2% | -1.2% | -6.5% | 3.3% | 0.50 |
| Rash | 1 | 1 | 1.1% | 1 | 1 | 1.2% | 0.0% | -5.5% | 5.3% | 0.99 |
| Skin irritation | 7 | 6 | 6.9% | 0 | 0 | 0.0% | 6.9% | 1.9% | 14.5% | 0.03 |
| Hot flush | 1 | 1 | 1.1% | 0 | 0 | 0.0% | 1.1% | -3.3% | 6.4% | 0.99 |

An adverse event was present if the participant rated that it was at least possibly associated with the intervention. Participants rated the severity of the adverse events as mild, moderate, or severe, which were assessed by the investigator. P values represent difference between groups. LB, lower bound; UB, upper bound.

**Supplementary Appendix**

Home-based transcranial direct current stimulation RCT in major depression

### Table S20. All unanticipated device related adverse events rated as mild

| | Active (N = 87) | | | Sham (N = 86) | | | Group Difference | | | |
|---|---|---|---|---|---|---|---|---|---|---|
| | **Events** | **Subjs** | **%** | **Events** | **Subjs** | **%** | **Δ** | **LB** | **UB** | **p** |
| **All Events** | **29** | **21** | **24.1%** | **16** | **14** | **16.3%** | **7.9%** | **-4.5%** | **20.3%** | **0.26** |
| **Ear and labyrinth disorders** | **2** | **2** | **2.3%** | **1** | **1** | **1.2%** | **1.1%** | **-4.5%** | **7.0%** | **0.99** |
| Tinnitus | 2 | 2 | 2.3% | 1 | 1 | 1.2% | 1.1% | -4.5% | 7.0% | 0.99 |
| **Eye disorders** | **4** | **3** | **3.4%** | **1** | **1** | **1.2%** | **2.3%** | **-3.3%** | **8.9%** | **0.62** |
| Photopsia | 2 | 2 | 2.3% | 1 | 1 | 1.2% | 1.1% | -4.5% | 7.0% | 0.99 |
| Vision blurred | 2 | 1 | 1.1% | 0 | 0 | 0.0% | 1.1% | -3.3% | 6.4% | 0.99 |
| **Gastrointestinal disorders** | **3** | **2** | **2.3%** | **1** | **1** | **1.2%** | **1.1%** | **-4.5%** | **7.0%** | **0.99** |
| Nausea | 3 | 2 | 2.3% | 1 | 1 | 1.2% | 1.1% | -4.5% | 7.0% | 0.99 |
| **Injury, poisoning and procedural complications** | **1** | **1** | **1.1%** | **0** | **0** | **0.0%** | **1.1%** | **-3.3%** | **6.4%** | **0.99** |
| Burns first degree | 1 | 1 | 1.1% | 0 | 0 | 0.0% | 1.1% | -3.3% | 6.4% | 0.99 |
| **Nervous system disorders** | **6** | **5** | **5.7%** | **6** | **6** | **7.0%** | **-1.2%** | **-9.6%** | **6.8%** | **0.77** |
| Burning sensation | 1 | 1 | 1.1% | 0 | 0 | 0.0% | 1.1% | -3.3% | 6.4% | 0.99 |
| Dizziness | 4 | 3 | 3.4% | 0 | 0 | 0.0% | 3.4% | -1.0% | 9.8% | 0.25 |
| Dysgeusia | 0 | 0 | 0.0% | 1 | 1 | 1.2% | -1.2% | -6.5% | 3.3% | 0.50 |
| Headache | 0 | 0 | 0.0% | 1 | 1 | 1.2% | -1.2% | -6.5% | 3.3% | 0.50 |
| Hyperaesthesia | 0 | 0 | 0.0% | 1 | 1 | 1.2% | -1.2% | -6.5% | 3.3% | 0.50 |
| Ophthalmic migraine | 0 | 0 | 0.0% | 1 | 1 | 1.2% | -1.2% | -6.5% | 3.3% | 0.50 |
| Paraesthesia | 1 | 1 | 1.1% | 2 | 2 | 2.3% | -1.2% | -7.2% | 4.4% | 0.62 |
| **Psychiatric disorders** | **2** | **1** | **1.1%** | **2** | **2** | **2.3%** | **-1.2%** | **-7.2%** | **4.4%** | **0.62** |
| Aggression | 0 | 0 | 0.0% | 1 | 1 | 1.2% | -1.2% | -6.5% | 3.3% | 0.50 |
| Anxiety | 0 | 0 | 0.0% | 1 | 1 | 1.2% | -1.2% | -6.5% | 3.3% | 0.50 |
| Depressed mood | 1 | 1 | 1.1% | 0 | 0 | 0.0% | 1.1% | -3.3% | 6.4% | 0.99 |
| Tension | 1 | 1 | 1.1% | 0 | 0 | 0.0% | 1.1% | -3.3% | 6.4% | 0.99 |
| **Skin and subcutaneous tissue disorders** | **10** | **10** | **11.5%** | **5** | **5** | **5.8%** | **5.7%** | **-3.3%** | **15.0%** | **0.28** |
| Acne | 0 | 0 | 0.0% | 1 | 1 | 1.2% | -1.2% | -6.5% | 3.3% | 0.50 |
| Dry skin | 4 | 4 | 4.6% | 4 | 4 | 4.7% | -0.1% | -7.5% | 7.3% | 0.99 |
| Rash | 1 | 1 | 1.1% | 0 | 0 | 0.0% | 1.1% | -3.3% | 6.4% | 0.99 |
| Skin irritation | 5 | 5 | 5.7% | 0 | 0 | 0.0% | 5.7% | 1.0% | 12.9% | 0.06 |
| **Vascular disorders** | **1** | **1** | **1.1%** | **0** | **0** | **0.0%** | **1.1%** | **-3.3%** | **6.4%** | **0.99** |
| Hot flush | 1 | 1 | 1.1% | 0 | 0 | 0.0% | 1.1% | -3.3% | 6.4% | 0.99 |

An adverse event was present if the participant rated that it was at least possibly associated with the intervention. Participants rated the severity of the adverse events as mild, moderate, or severe, which were assessed by the investigator. P values represent difference between groups. LB, lower bound; UB, upper bound.

**Supplementary Appendix**

Home-based transcranial direct current stimulation RCT in major depression

### Table S21. All unanticipated device related adverse events rated as moderate

| | Active (N= 87) | | | Sham (N= 86) | | | Group Difference | | | |
|---|---|---|---|---|---|---|---|---|---|---|
| | Events | Subjs | % | Events | Subjs | % | Δ | LB | UB | p |
| **All Events** | **16** | **13** | **14.9%** | **10** | **8** | **9.3%** | **5.6%** | **-4.5%** | **16.1%** | **0.35** |
| **Ear and labyrinth disorders** | **0** | **0** | **0.0%** | **1** | **1** | **1.2%** | **-1.2%** | **-6.5%** | **3.3%** | **0.50** |
| Tinnitus | 0 | 0 | 0.0% | 1 | 1 | 1.2% | -1.2% | -6.5% | 3.3% | 0.50 |
| **General disorders and administration site conditions** | **3** | **3** | **3.4%** | **2** | **2** | **2.3%** | **1.1%** | **-5.2%** | **8.0%** | **0.99** |
| Fatigue | 1 | 1 | 1.1% | 0 | 0 | 0.0% | 1.1% | -3.3% | 6.4% | 0.99 |
| Malaise | 1 | 1 | 1.1% | 0 | 0 | 0.0% | 1.1% | -3.3% | 6.4% | 0.99 |
| Pain | 1 | 1 | 1.1% | 2 | 2 | 2.3% | -1.2% | -7.2% | 4.4% | 0.62 |
| **Infections and infestations** | **1** | **1** | **1.1%** | **1** | **1** | **1.2%** | **0.0%** | **-5.5%** | **5.3%** | **0.99** |
| Nasopharyngitis | 1 | 1 | 1.1% | 0 | 0 | 0.0% | 1.1% | -3.3% | 6.4% | 0.99 |
| Sinusitis | 0 | 0 | 0.0% | 1 | 1 | 1.2% | -1.2% | -6.5% | 3.3% | 0.50 |
| **Injury, poisoning and procedural complications** | **1** | **1** | **1.1%** | **0** | **0** | **0.0%** | **1.1%** | **-3.3%** | **6.4%** | **0.99** |
| Burns first degree | 1 | 1 | 1.1% | 0 | 0 | 0.0% | 1.1% | -3.3% | 6.4% | 0.99 |
| **Nervous system disorders** | **1** | **1** | **1.1%** | **2** | **2** | **2.3%** | **-1.2%** | **-7.2%** | **4.4%** | **0.62** |
| Dizziness | 0 | 0 | 0.0% | 1 | 1 | 1.2% | -1.2% | -6.5% | 3.3% | 0.50 |
| Paraesthesia | 1 | 1 | 1.1% | 1 | 1 | 1.2% | 0.0% | -5.5% | 5.3% | 0.99 |
| **Psychiatric disorders** | **3** | **3** | **3.4%** | **2** | **2** | **2.3%** | **1.1%** | **-5.2%** | **8.0%** | **0.99** |
| Anxiety | 1 | 1 | 1.1% | 0 | 0 | 0.0% | 1.1% | -3.3% | 6.4% | 0.99 |
| Insomnia | 1 | 1 | 1.1% | 2 | 2 | 2.3% | -1.2% | -7.2% | 4.4% | 0.62 |
| Panic attack | 1 | 1 | 1.1% | 0 | 0 | 0.0% | 1.1% | -3.3% | 6.4% | 0.99 |
| **Skin and subcutaneous tissue disorders** | **7** | **6** | **6.9%** | **2** | **2** | **2.3%** | **4.6%** | **-2.2%** | **12.5%** | **0.28** |
| Acne | 1 | 1 | 1.1% | 0 | 0 | 0.0% | 1.1% | -3.3% | 6.4% | 0.99 |
| Alopecia | 1 | 1 | 1.1% | 0 | 0 | 0.0% | 1.1% | -3.3% | 6.4% | 0.99 |
| Dry skin | 4 | 4 | 4.6% | 0 | 0 | 0.0% | 4.6% | 0.1% | 11.4% | 0.12 |
| Pruritus | 0 | 0 | 0.0% | 1 | 1 | 1.2% | -1.2% | -6.5% | 3.3% | 0.50 |
| Rash | 0 | 0 | 0.0% | 1 | 1 | 1.2% | -1.2% | -6.5% | 3.3% | 0.50 |
| Skin irritation | 1 | 1 | 1.1% | 0 | 0 | 0.0% | 1.1% | -3.3% | 6.4% | 0.99 |

An adverse event was present if the participant rated that it was at least possibly associated with the intervention. Participants rated the severity of the adverse events as mild, moderate, or severe, which were assessed by the investigator. P values represent difference between groups. LB, lower bound; UB, upper bound.

**Supplementary Appendix**

Home-based transcranial direct current stimulation RCT in major depression

### Table S22. All unanticipated device related adverse events rated as severe

| | Active (N= 87) | | | Sham (N= 86) | | | Group Difference | | | |
|---|---|---|---|---|---|---|---|---|---|---|
| | Events | Subjs | % | Events | Subjs | % | Δ | LB | UB | p |
| **All Events** | **3** | **3** | **3.4%** | **1** | **1** | **1.2%** | **2.3%** | **-3.3%** | **8.9%** | **0.62** |
| **Nervous system disorders** | **1** | **1** | **1.1%** | **1** | **1** | **1.2%** | **0.0%** | **-5.5%** | **5.3%** | **0.99** |
| Dizziness | 0 | 0 | 0.0% | 1 | 1 | 1.2% | -1.2% | -6.5% | 3.3% | 0.50 |
| Migraine | 1 | 1 | 1.1% | 0 | 0 | 0.0% | 1.1% | -3.3% | 6.4% | 0.99 |
| **Skin and subcutaneous tissue disorders** | **2** | **2** | **2.3%** | **0** | **0** | **0.0%** | **2.3%** | **-2.2%** | **8.1%** | **0.50** |
| Dry skin | 1 | 1 | 1.1% | 0 | 0 | 0.0% | 1.1% | -3.3% | 6.4% | 0.99 |
| Skin irritation | 1 | 1 | 1.1% | 0 | 0 | 0.0% | 1.1% | -3.3% | 6.4% | 0.99 |

An adverse event was present if the participant rated that it was at least possibly associated with the intervention. Participants rated the severity of the adverse events as mild, moderate, or severe, which were assessed by the investigator. P values represent difference between groups. LB, lower bound; UB, upper bound.

**Supplementary Appendix**

Home-based transcranial direct current stimulation RCT in major depression

### Table S23. All unanticipated device related adverse events by days post randomisation

| | Days Post-randomization | | | | | | | | | | | | | | | | | | | |
|---|---|---|---|---|---|---|---|---|---|---|---|---|---|---|---|---|---|---|---|---|
| | Missing | | <0 | | 0-7 | | 7-14 | | 14-28 | | 28-42 | | 42-56 | | 56-70 | | >70 | | Total | |
| | A | S | A | S | A | S | A | S | A | S | A | S | A | S | A | S | A | S | A | S |
| **All Events** | **0** | **1** | **0** | **0** | **18** | **7** | **3** | **2** | **9** | **6** | **8** | **1** | **4** | **6** | **4** | **2** | **2** | **2** | **48** | **27** |
| **Ear and labyrinth disorders** | **0** | **0** | **0** | **0** | **0** | **0** | **0** | **0** | **0** | **2** | **0** | **0** | **0** | **0** | **1** | **0** | **1** | **0** | **2** | **2** |
| Tinnitus | 0 | 0 | 0 | 0 | 0 | 0 | 0 | 0 | 0 | 2 | 0 | 0 | 0 | 0 | 1 | 0 | 1 | 0 | 2 | 2 |
| **Eye disorders** | **0** | **1** | **0** | **0** | **1** | **0** | **1** | **0** | **1** | **0** | **1** | **0** | **0** | **0** | **0** | **0** | **0** | **0** | **4** | **1** |
| Photopsia | 0 | 1 | 0 | 0 | 1 | 0 | 0 | 0 | 0 | 0 | 1 | 0 | 0 | 0 | 0 | 0 | 0 | 0 | 2 | 1 |
| Vision blurred | 0 | 0 | 0 | 0 | 0 | 0 | 1 | 0 | 1 | 0 | 0 | 0 | 0 | 0 | 0 | 0 | 0 | 0 | 2 | 0 |
| **Gastrointestinal disorders** | **0** | **0** | **0** | **0** | **1** | **0** | **0** | **0** | **2** | **0** | **0** | **0** | **0** | **1** | **0** | **0** | **0** | **0** | **3** | **1** |
| Nausea | 0 | 0 | 0 | 0 | 1 | 0 | 0 | 0 | 2 | 0 | 0 | 0 | 0 | 1 | 0 | 0 | 0 | 0 | 3 | 1 |
| **General disorders and administration site conditions** | **0** | **0** | **0** | **0** | **2** | **0** | **0** | **0** | **0** | **1** | **0** | **0** | **0** | **1** | **0** | **0** | **1** | **0** | **3** | **2** |
| Fatigue | 0 | 0 | 0 | 0 | 1 | 0 | 0 | 0 | 0 | 0 | 0 | 0 | 0 | 0 | 0 | 0 | 0 | 0 | 1 | 0 |
| Malaise | 0 | 0 | 0 | 0 | 0 | 0 | 0 | 0 | 0 | 0 | 0 | 0 | 0 | 0 | 0 | 0 | 1 | 0 | 1 | 0 |
| Pain | 0 | 0 | 0 | 0 | 1 | 0 | 0 | 0 | 0 | 1 | 0 | 0 | 0 | 1 | 0 | 0 | 0 | 0 | 1 | 2 |
| **Infections and infestations** | **0** | **0** | **0** | **0** | **0** | **0** | **0** | **0** | **1** | **1** | **0** | **0** | **0** | **0** | **0** | **0** | **0** | **0** | **1** | **1** |
| Nasopharyngitis | 0 | 0 | 0 | 0 | 0 | 0 | 0 | 0 | 1 | 0 | 0 | 0 | 0 | 0 | 0 | 0 | 0 | 0 | 1 | 0 |
| Sinusitis | 0 | 0 | 0 | 0 | 0 | 0 | 0 | 0 | 0 | 1 | 0 | 0 | 0 | 0 | 0 | 0 | 0 | 0 | 0 | 1 |
| **Injury, poisoning and procedural complications** | **0** | **0** | **0** | **0** | **0** | **0** | **0** | **0** | **1** | **0** | **1** | **0** | **0** | **0** | **0** | **0** | **0** | **0** | **2** | **0** |
| Burns first degree | 0 | 0 | 0 | 0 | 0 | 0 | 0 | 0 | 1 | 0 | 1 | 0 | 0 | 0 | 0 | 0 | 0 | 0 | 2 | 0 |
| **Nervous system disorders** | **0** | **0** | **0** | **0** | **5** | **5** | **1** | **0** | **0** | **0** | **0** | **0** | **1** | **3** | **1** | **1** | **0** | **0** | **8** | **9** |
| Burning sensation | 0 | 0 | 0 | 0 | 0 | 0 | 0 | 0 | 0 | 0 | 0 | 0 | 1 | 0 | 0 | 0 | 0 | 0 | 1 | 0 |
| Dizziness | 0 | 0 | 0 | 0 | 3 | 1 | 0 | 0 | 0 | 0 | 0 | 0 | 0 | 1 | 1 | 0 | 0 | 0 | 4 | 2 |
| Dysgeusia | 0 | 0 | 0 | 0 | 0 | 1 | 0 | 0 | 0 | 0 | 0 | 0 | 0 | 0 | 0 | 0 | 0 | 0 | 0 | 1 |
| Headache | 0 | 0 | 0 | 0 | 0 | 0 | 0 | 0 | 0 | 0 | 0 | 0 | 0 | 0 | 0 | 1 | 0 | 0 | 0 | 1 |
| Hyperaesthesia | 0 | 0 | 0 | 0 | 0 | 0 | 0 | 0 | 0 | 0 | 0 | 0 | 0 | 1 | 0 | 0 | 0 | 0 | 0 | 1 |
| Migraine | 0 | 0 | 0 | 0 | 0 | 0 | 1 | 0 | 0 | 0 | 0 | 0 | 0 | 0 | 0 | 0 | 0 | 0 | 1 | 0 |
| Ophthalmic migraine | 0 | 0 | 0 | 0 | 0 | 1 | 0 | 0 | 0 | 0 | 0 | 0 | 0 | 0 | 0 | 0 | 0 | 0 | 0 | 1 |
| Paraesthesia | 0 | 0 | 0 | 0 | 2 | 2 | 0 | 0 | 0 | 0 | 0 | 0 | 0 | 1 | 0 | 0 | 0 | 0 | 2 | 3 |

| | | | | | | | | | | | | | | | | | | | | |
|---|---|---|---|---|---|---|---|---|---|---|---|---|---|---|---|---|---|---|---|---|
| **Psychiatric disorders** | **0** | **0** | **0** | **0** | **4** | **0** | **1** | **1** | **0** | **1** | **0** | **1** | **0** | **0** | **0** | **0** | **0** | **1** | **5** | **4** |
| Aggression | 0 | 0 | 0 | 0 | 0 | 0 | 0 | 0 | 0 | 1 | 0 | 0 | 0 | 0 | 0 | 0 | 0 | 0 | 0 | 1 |
| Anxiety | 0 | 0 | 0 | 0 | 1 | 0 | 0 | 1 | 0 | 0 | 0 | 0 | 0 | 0 | 0 | 0 | 0 | 0 | 1 | 1 |
| Depressed mood | 0 | 0 | 0 | 0 | 1 | 0 | 0 | 0 | 0 | 0 | 0 | 0 | 0 | 0 | 0 | 0 | 0 | 0 | 1 | 0 |
| Insomnia | 0 | 0 | 0 | 0 | 0 | 0 | 1 | 0 | 0 | 0 | 0 | 1 | 0 | 0 | 0 | 0 | 0 | 1 | 1 | 2 |
| Panic attack | 0 | 0 | 0 | 0 | 1 | 0 | 0 | 0 | 0 | 0 | 0 | 0 | 0 | 0 | 0 | 0 | 0 | 0 | 1 | 0 |
| Tension | 0 | 0 | 0 | 0 | 1 | 0 | 0 | 0 | 0 | 0 | 0 | 0 | 0 | 0 | 0 | 0 | 0 | 0 | 1 | 0 |
| **Skin and subcutaneous tissue disorders** | **0** | **0** | **0** | **0** | **5** | **2** | **0** | **1** | **3** | **1** | **6** | **0** | **3** | **1** | **2** | **1** | **0** | **1** | **19** | **7** |
| Acne | 0 | 0 | 0 | 0 | 0 | 1 | 0 | 0 | 0 | 0 | 0 | 0 | 1 | 0 | 0 | 0 | 0 | 0 | 1 | 1 |
| Alopecia | 0 | 0 | 0 | 0 | 0 | 0 | 0 | 0 | 0 | 0 | 0 | 0 | 1 | 0 | 0 | 0 | 0 | 0 | 1 | 0 |
| Dry skin | 0 | 0 | 0 | 0 | 1 | 1 | 0 | 0 | 2 | 0 | 5 | 0 | 0 | 1 | 1 | 1 | 0 | 1 | 9 | 4 |
| Pruritus | 0 | 0 | 0 | 0 | 0 | 0 | 0 | 1 | 0 | 0 | 0 | 0 | 0 | 0 | 0 | 0 | 0 | 0 | 0 | 1 |
| Rash | 0 | 0 | 0 | 0 | 0 | 0 | 0 | 0 | 0 | 1 | 1 | 0 | 0 | 0 | 0 | 0 | 0 | 0 | 1 | 1 |
| Skin irritation | 0 | 0 | 0 | 0 | 4 | 0 | 0 | 0 | 1 | 0 | 0 | 0 | 1 | 0 | 1 | 0 | 0 | 0 | 7 | 0 |
| **Vascular disorders** | **0** | **0** | **0** | **0** | **0** | **0** | **0** | **0** | **1** | **0** | **0** | **0** | **0** | **0** | **0** | **0** | **0** | **0** | **1** | **0** |
| Hot flush | 0 | 0 | 0 | 0 | 0 | 0 | 0 | 0 | 1 | 0 | 0 | 0 | 0 | 0 | 0 | 0 | 0 | 0 | 1 | 0 |

An adverse event was present if the participant rated that it was at least possibly associated with the intervention. Participants rated the severity of the adverse events as mild, moderate, or severe, which were assessed by the investigator. P values represent difference between groups. LB, lower bound; UB, upper bound.

**Supplementary Appendix**
Home-based transcranial direct current stimulation RCT in major depression

### Table S24. All unanticipated adverse events

| | Active (N= 87) | | | Sham (N= 86) | | | Group Difference | | | |
|---|---|---|---|---|---|---|---|---|---|---|
| | Events | Subjs | % | Events | Subjs | % | Δ | LB | UB | p |
| **All Events** | **90** | **53** | **60.9%** | **55** | **37** | **43.0%** | **17.9%** | **2.0%** | **32.5%** | **0.02** |
| **Blood and lymphatic system disorders** | **1** | **1** | **1.1%** | **0** | **0** | **0.0%** | **1.1%** | **-3.3%** | **6.4%** | **0.99** |
| Lymphadenopathy | 1 | 1 | 1.1% | 0 | 0 | 0.0% | 1.1% | -3.3% | 6.4% | 0.99 |
| **Ear and labyrinth disorders** | **2** | **2** | **2.3%** | **2** | **2** | **2.3%** | **0.0%** | **-6.2%** | **6.0%** | **0.99** |
| Tinnitus | 2 | 2 | 2.3% | 2 | 2 | 2.3% | 0.0% | -6.2% | 6.0% | 0.99 |
| **Eye disorders** | **4** | **3** | **3.4%** | **1** | **1** | **1.2%** | **2.3%** | **-3.3%** | **8.9%** | **0.62** |
| Photopsia | 2 | 2 | 2.3% | 1 | 1 | 1.2% | 1.1% | -4.5% | 7.0% | 0.99 |
| Vision blurred | 2 | 1 | 1.1% | 0 | 0 | 0.0% | 1.1% | -3.3% | 6.4% | 0.99 |
| **Gastrointestinal disorders** | **8** | **6** | **6.9%** | **1** | **1** | **1.2%** | **5.7%** | **-0.4%** | **13.6%** | **0.12** |
| Diarrhoea | 1 | 1 | 1.1% | 0 | 0 | 0.0% | 1.1% | -3.3% | 6.4% | 0.99 |
| Irritable bowel syndrome | 2 | 2 | 2.3% | 0 | 0 | 0.0% | 2.3% | -2.2% | 8.1% | 0.50 |
| Nausea | 3 | 2 | 2.3% | 1 | 1 | 1.2% | 1.1% | -4.5% | 7.0% | 0.99 |
| Vomiting | 2 | 1 | 1.1% | 0 | 0 | 0.0% | 1.1% | -3.3% | 6.4% | 0.99 |
| **General disorders and administration site conditions** | **3** | **3** | **3.4%** | **3** | **3** | **3.5%** | **0.0%** | **-6.8%** | **6.8%** | **0.99** |
| Chest pain | 0 | 0 | 0.0% | 1 | 1 | 1.2% | -1.2% | -6.5% | 3.3% | 0.50 |
| Fatigue | 1 | 1 | 1.1% | 0 | 0 | 0.0% | 1.1% | -3.3% | 6.4% | 0.99 |
| Malaise | 1 | 1 | 1.1% | 0 | 0 | 0.0% | 1.1% | -3.3% | 6.4% | 0.99 |
| Pain | 1 | 1 | 1.1% | 2 | 2 | 2.3% | -1.2% | -7.2% | 4.4% | 0.62 |
| **Infections and infestations** | **25** | **22** | **25.3%** | **25** | **19** | **22.1%** | **3.2%** | **-9.8%** | **16.1%** | **0.72** |
| Bacterial infection | 0 | 0 | 0.0% | 1 | 1 | 1.2% | -1.2% | -6.5% | 3.3% | 0.50 |
| COVID-19 | 6 | 6 | 6.9% | 5 | 5 | 5.8% | 1.1% | -7.0% | 9.3% | 0.99 |
| Cystitis | 1 | 1 | 1.1% | 0 | 0 | 0.0% | 1.1% | -3.3% | 6.4% | 0.99 |
| Gastroenteritis viral | 1 | 1 | 1.1% | 1 | 1 | 1.2% | 0.0% | -5.5% | 5.3% | 0.99 |
| Influenza | 4 | 3 | 3.4% | 6 | 6 | 7.0% | -3.5% | -11.5% | 3.7% | 0.33 |
| Lower respiratory tract infection | 1 | 1 | 1.1% | 1 | 1 | 1.2% | 0.0% | -5.5% | 5.3% | 0.99 |
| Nasopharyngitis | 12 | 11 | 12.6% | 8 | 7 | 8.1% | 4.5% | -5.1% | 14.5% | 0.46 |
| Pneumonia | 0 | 0 | 0.0% | 1 | 1 | 1.2% | -1.2% | -6.5% | 3.3% | 0.50 |

| | | | | | | | | | | |
|---|---|---|---|---|---|---|---|---|---|---|
| Sinusitis | 0 | 0 | 0.0% | 1 | 1 | 1.2% | -1.2% | -6.5% | 3.3% | 0.50 |
| Viral infection | 0 | 0 | 0.0% | 1 | 1 | 1.2% | -1.2% | -6.5% | 3.3% | 0.50 |
| **Injury, poisoning and procedural complications** | **4** | **4** | **4.6%** | **0** | **0** | **0.0%** | **4.6%** | **0.1%** | **11.4%** | **0.12** |
| Burns first degree | 2 | 2 | 2.3% | 0 | 0 | 0.0% | 2.3% | -2.2% | 8.1% | 0.50 |
| Head injury | 1 | 1 | 1.1% | 0 | 0 | 0.0% | 1.1% | -3.3% | 6.4% | 0.99 |
| Limb injury | 1 | 1 | 1.1% | 0 | 0 | 0.0% | 1.1% | -3.3% | 6.4% | 0.99 |
| **Metabolism and nutrition disorders** | **1** | **1** | **1.1%** | **0** | **0** | **0.0%** | **1.1%** | **-3.3%** | **6.4%** | **0.99** |
| Gout | 1 | 1 | 1.1% | 0 | 0 | 0.0% | 1.1% | -3.3% | 6.4% | 0.99 |
| **Neoplasms benign, malignant and unspecified (incl cysts and polyps)** | **1** | **1** | **1.1%** | **0** | **0** | **0.0%** | **1.1%** | **-3.3%** | **6.4%** | **0.99** |
| Acrochordon | 1 | 1 | 1.1% | 0 | 0 | 0.0% | 1.1% | -3.3% | 6.4% | 0.99 |
| **Nervous system disorders** | **13** | **9** | **10.3%** | **9** | **8** | **9.3%** | **1.0%** | **-8.5%** | **10.6%** | **0.99** |
| Burning sensation | 1 | 1 | 1.1% | 0 | 0 | 0.0% | 1.1% | -3.3% | 6.4% | 0.99 |
| Dizziness | 5 | 3 | 3.4% | 2 | 2 | 2.3% | 1.1% | -5.2% | 8.0% | 0.99 |
| Dysgeusia | 0 | 0 | 0.0% | 1 | 1 | 1.2% | -1.2% | -6.5% | 3.3% | 0.50 |
| Headache | 0 | 0 | 0.0% | 1 | 1 | 1.2% | -1.2% | -6.5% | 3.3% | 0.50 |
| Hyperaesthesia | 0 | 0 | 0.0% | 1 | 1 | 1.2% | -1.2% | -6.5% | 3.3% | 0.50 |
| Migraine | 4 | 3 | 3.4% | 0 | 0 | 0.0% | 3.4% | -1.0% | 9.8% | 0.25 |
| Migraine with aura | 1 | 1 | 1.1% | 0 | 0 | 0.0% | 1.1% | -3.3% | 6.4% | 0.99 |
| Ophthalmic migraine | 0 | 0 | 0.0% | 1 | 1 | 1.2% | -1.2% | -6.5% | 3.3% | 0.50 |
| Paraesthesia | 2 | 2 | 2.3% | 3 | 3 | 3.5% | -1.2% | -8.0% | 5.1% | 0.68 |
| **Psychiatric disorders** | **5** | **4** | **4.6%** | **4** | **4** | **4.7%** | **-0.1%** | **-7.5%** | **7.3%** | **0.99** |
| Aggression | 0 | 0 | 0.0% | 1 | 1 | 1.2% | -1.2% | -6.5% | 3.3% | 0.50 |
| Anxiety | 1 | 1 | 1.1% | 1 | 1 | 1.2% | 0.0% | -5.5% | 5.3% | 0.99 |
| Depressed mood | 1 | 1 | 1.1% | 0 | 0 | 0.0% | 1.1% | -3.3% | 6.4% | 0.99 |
| Insomnia | 1 | 1 | 1.1% | 2 | 2 | 2.3% | -1.2% | -7.2% | 4.4% | 0.62 |
| Panic attack | 1 | 1 | 1.1% | 0 | 0 | 0.0% | 1.1% | -3.3% | 6.4% | 0.99 |
| Tension | 1 | 1 | 1.1% | 0 | 0 | 0.0% | 1.1% | -3.3% | 6.4% | 0.99 |
| **Reproductive system and breast disorders** | **0** | **0** | **0.0%** | **1** | **1** | **1.2%** | **-1.2%** | **-6.5%** | **3.3%** | **0.50** |
| Heavy menstrual bleeding | 0 | 0 | 0.0% | 1 | 1 | 1.2% | -1.2% | -6.5% | 3.3% | 0.50 |
| **Respiratory, thoracic and mediastinal disorders** | **1** | **1** | **1.1%** | **2** | **1** | **1.2%** | **0.0%** | **-5.5%** | **5.3%** | **0.99** |
| Asthma | 0 | 0 | 0.0% | 1 | 1 | 1.2% | -1.2% | -6.5% | 3.3% | 0.50 |
| Dyspnoea | 0 | 0 | 0.0% | 1 | 1 | 1.2% | -1.2% | -6.5% | 3.3% | 0.50 |

| | | | | | | | | | | |
|---|---|---|---|---|---|---|---|---|---|---|
| Oropharyngeal pain | 1 | 1 | 1.1% | 0 | 0 | 0.0% | 1.1% | -3.3% | 6.4% | 0.99 |
| **Skin and subcutaneous tissue disorders** | **19** | **17** | **19.5%** | **7** | **7** | **8.1%** | **11.4%** | **1.0%** | **22.3%** | **0.05** |
| Acne | 1 | 1 | 1.1% | 1 | 1 | 1.2% | 0.0% | -5.5% | 5.3% | 0.99 |
| Alopecia | 1 | 1 | 1.1% | 0 | 0 | 0.0% | 1.1% | -3.3% | 6.4% | 0.99 |
| Dry skin | 9 | 9 | 10.3% | 4 | 4 | 4.7% | 5.7% | -2.5% | 14.8% | 0.25 |
| Pruritus | 0 | 0 | 0.0% | 1 | 1 | 1.2% | -1.2% | -6.5% | 3.3% | 0.50 |
| Rash | 1 | 1 | 1.1% | 1 | 1 | 1.2% | 0.0% | -5.5% | 5.3% | 0.99 |
| Skin irritation | 7 | 6 | 6.9% | 0 | 0 | 0.0% | 6.9% | 1.9% | 14.5% | 0.03 |
| **Vascular disorders** | **3** | **2** | **2.3%** | **0** | **0** | **0.0%** | **2.3%** | **-2.2%** | **8.1%** | **0.50** |
| Hot flush | 1 | 1 | 1.1% | 0 | 0 | 0.0% | 1.1% | -3.3% | 6.4% | 0.99 |
| Hypertension | 2 | 1 | 1.1% | 0 | 0 | 0.0% | 1.1% | -3.3% | 6.4% | 0.99 |

All unanticipated adverse events that were reported up to week 10 including those that weren't associated with the intervention. Participants rated the severity of the adverse events as mild, moderate, or severe, which were assessed by the investigator. P values represent difference between groups. LB, lower bound; UB, upper bound.

**Supplementary Appendix**

Home-based transcranial direct current stimulation RCT in major depression

### Table S25. Response and remission rates at all time points

| | Week 1 | | | | Week 4 | | | | Week 7 | | | | Week 10 | | | |
|---|---|---|---|---|---|---|---|---|---|---|---|---|---|---|---|---|
| Outcome – n (%) | Active tDCS (N=87) | Sham tDCS (N=86) | Odds Ratio (95% CI) | P Value | Active tDCS (N=87) | Sham tDCS (N=86) | Odds Ratio (95% CI) | P Value | Active tDCS (N=87) | Sham tDCS (N=86) | Odds Ratio (95% CI) | P Value | Active tDCS (N=87) | Sham tDCS (N=86) | Odds Ratio (95% CI) | P Value |
| **HDRS** | | | | | | | | | | | | | | | | |
| response | 9 (10.9) | 12 (13.2) | 0.80 (0.31 to 2.06) | 0.67 | 27 (35.1) | 13 (16.2) | 2.80 (1.30 to 6.02) | 0.008 | 35 (47.8) | 22 (28.8) | 2.27 (1.14 to 4.51) | 0.02 | 41 (54.4%) | 21 (26.9%) | 3.25 (1.57 to 6.74) | 0.001 |
| remission | 4 (4.7) | 6 (6.5) | 0.70 (0.18 to 2.73) | 0.66 | 18 (23.8) | 11 (13.6) | 1.99 (0.85 to 4.66) | 0.13 | 27 (37.1) | 15 (20.5) | 2.30 (1.05 to 5.08) | 0.04 | 34 (44.9%) | 17 (21.8%) | 2.93 (1.41 to 6.09) | 0.004 |
| **MADRS** | | | | | | | | | | | | | | | | |
| response | 10 (11.8) | 7 (7.6) | 1.63 (0.57 to 4.62) | 0.39 | 30 (39.0) | 13 (16.1) | 3.35 (1.55 to 7.22) | 0.002 | 35 (47.1) | 23 (29.0) | 2.18 (1.09 to 4.47) | 0.03 | 46 (63.0%) | 25 (31.6%) | 3.70 (1.82 to 7.52) | <0.001 |
| remission | 7 (4.4) | 9 (3.7) | 1.21 (0.36 to 4.08) | 0.81 | 25 (32.7) | 11 (12.5) | 3.42 (1.45 to 8.09) | 0.004 | 34 (47.7) | 19 (21.8) | 3.28 (1.49 to 7.20) | 0.002 | 42 (57.5%) | 25 (29.4%) | 3.26 (1.53 to 6.94) | 0.002 |
| **MADRS-s** | | | | | | | | | | | | | | | | |
| response | 8 (9.7) | 6 (7.4) | 1.35 (0.43 to 4.27) | 0.62 | 22 (27.8) | 11 (15.2) | 2.16 (0.95 to 4.93) | 0.07 | 25 (34.8) | 15 (23.5) | 1.72 (0.82 to 3.62) | 0.16 | 30 (49.1%) | 14 (24.0%) | 3.06 (1.43 to 6.56) | 0.004 |
| remission | 11 (8.6) | 9 (5.7) | 1.59 (0.51 to 4.94) | 0.52 | 23 (29.3) | 13 (16.2) | 2.15 (0.95 to 4.89) | 0.07 | 24 (33.2) | 16 (21.5) | 1.81 (0.83 to 3.97) | 0.14 | 32 (53.8%) | 18 (23.4%) | 3.83 (1.61 to 9.13) | 0.002 |

Response was defined as a 50% decrease (indicating less depression) in total scores from baseline. Remission was defined as a score of 7 or fewer points on the HDRS or as 10 or fewer points on the MADRS or as 12 or fewer points of the MADRS-s at 10 weeks. Percentages are estimated based on odds ratios. tDCS, transcranial direct current stimulation. CI, confidence interval. HDRS, Hamilton Depression Rating Scale[3]; MADRS, Montgomery-Åsberg Depression Rating Scale[13]; MADRS-s, Montgomery-Åsberg Depression Rating Scale-self report[14]. HDRS scores range from 0 to 52 (minimal clinically significant difference = 3 points), MADRS scores range from 0 to 60; MADRS-s scores range from 0 to 54, with higher scores indicating more depression. P values represent difference between groups.

### Blinding Analysis

#### Table S26. Blinding evaluation mITT sample

| | Active (n = 85) | | Sham (n = 81) | | P value |
|---|---|---|---|---|---|
| | n | % | n | % | |
| **Correct arm assumed** | | | | | |
| No | 19 | 22% | 48 | 59% | 0.012 |
| Yes | 66 | 78% | 33 | 41% | |

n = 166. Week 10 completers and early termination sample. 8 participants did not complete a blinding analysis. P value compares how many participants guessed active or sham in both groups.

Participants were asked to guess which treatment they thought they had been receiving at the week 10 study visit or early termination visit. Of participants who were receiving active tDCS, 78% correctly speculated that they were receiving active tDCS and 22% that they were receiving inactive tDCS. Of participants who were receiving inactive tDCS, 59% speculated that they were receiving at active tDCS, and 41% that they were receiving inactive tDCS. There was a significant difference between the proportion of correct guesses about group allocation.

#### Table S27. Blinding evaluation of guess certainty

| | | Active (n = 85) | | Sham (n = 81) | |
|---|---|---|---|---|---|
| **Correct assumed arm** | **Certain (4 or 5 out of 1-5)** | **n** | **%** | **n** | **%** |
| No | No | 11 | 13% | 28 | 35% |
| No | Yes | 8 | 9% | 20 | 25% |
| Yes | No | 28 | 32% | 23 | 28% |
| Yes | Yes | 38 | 45% | 10 | 12% |

n = 166. Week 10 completers and early termination sample.

#### Table S28. Changes in depressive severity as measured by HDRS, MADRS and MADRS-s and quality of life as measured by EQ-5D-3L for participants who guessed they were receiving the active treatment at 10 weeks

| Outcome | Active (n = 66) | Sham (n = 48) | Difference or Odds Ratio (95% CI) | P value |
|---|---|---|---|---|
| **Primary Outcome** | | | | |
| Decrease in HDRS score | 10.3 ± 5.94 | 8.0 ± 5.76 | 2.3 (0.2 to 4.4) | 0.03 |
| **Secondary Outcomes** | | | | |
| HDRS | | | | |
| Clinical response | 35 (61.1%) | 16 (37.1%) | 2.66 (1.14 to 6.19) | 0.02 |
| Clinical remission | 29 (51.2%) | 12 (26.5%) | 2.91 (1.21 to 7.00) | 0.01 |
| MADRS | | | | |
| Decrease in score | 12.5 ± 8.59 | 8.7 ± 8.22 | 3.9 (0.8 to 6.9) | 0.01 |
| Clinical response | 40 (70.2%) | 18 (41.0%) | 3.40 (1.35 to 8.56) | 0.009 |
| Clinical remission | 36 (63.4%) | 17 (35.7%) | 3.10 (1.24 to 7.75) | 0.01 |
| MADRS-s | | | | |
| Decrease in score | 11.1 ± 8.47 | 8.6 ± 8.46 | 2.5 (-0.7 to 5.7) | 0.13 |
| Clinical response | 25 (51.9%) | 9 (25.7%) | 3.12 (1.20 to 8.11) | 0.01 |
| Clinical remission | 28 (60.2%) | 13 (28.7%) | 3.78 (1.30 to 11.02) | 0.009 |
| EQ-5D-3L | 0.08 ± 0.16 | 0.09 ± 0.15 | 0.02 (-0.03 to 0.07) | 0.38 |

HDRS, Hamilton Depression Rating Scale[3]; MADRS, Montgomery-Åsberg Depression Rating Scale[13]; MADRS-s, Montgomery-Åsberg Depression Rating Scale-self report[14]. EQ-5D-3L, quality of life measure[8–10]; CI, confidence interval. Mean values are presented with '±' standard deviation values. HDRS, MADRS, MADRS-s change ratings are the change in total ratings from baseline to week 10. Between-group differences are shown for the changes in scores from baseline to week 10, and odds ratios are shown for the outcomes for clinical response and remission. Percentages for clinical response and remission outcomes are estimated based on odds ratios. HDRS scores range from 0 to 52 (minimal clinically significant difference = 3 points), MADRS scores range from 0 to 60; MADRS-s scores range from 0 to 54, with higher scores indicating more depression. Clinical response was defined as a decrease in the score (indicating less depressive severity) of 50% or more from baseline to week 10. Clinical remission was defined as: HDRS score of 7 or less; MADRS score of 10 or less; MADRS-s score of 12 or less.

An analysis comparing only participants who believed that they were receiving active tDCS was completed to assess the active treatment effect over the placebo-sham effect. Group decreases and significant differences between groups are consistent with the overall trial results, with greater improvements in depressive symptoms in the active group compared to the sham group for HDRS and MADRS decrease in score at week 10, and for HDRS, MADRS and MADRS-s response and remission rates at week 10. These results indicate that participant beliefs alone about which treatment they were receiving cannot explain the results and suggests that there was a treatment effect. The MADRS-s decease in score was not significant between groups, which could suggest a greater placebo effect in self-reported depressive symptoms.

### Table 29. Changes in depressive severity as measured by HDRS, MADRS and MADRS-s and quality of life as measured by EQ-5D-3L for participants at the UK site at 10 weeks

| Outcome | Active (n = 58) | Sham (n = 57) | Difference or Odds Ratio (95% CI) | P value |
|---|---|---|---|---|
| **Primary Outcome** | | | | |
| Decrease in HDRS score | 8.0 ± 5.56 | 5.9 ± 5.49 | 2.1 (0.2 to 3.9) | 0.03 |
| **Secondary Outcomes** | | | | |
| HDRS | | | | |
| Clinical response | 24 (47.2%) | 11 (20.0%) | 3.57 (1.46 to 8.71) | 0.004 |
| Clinical remission | 18 (34.8%) | 7 (12.5%) | 3.73 (1.34 to 10.34) | 0.01 |
| MADRS | | | | |
| Decrease in score | 11.1 ± 8.91 | 7.6 ± 8.61 | 3.5 (0.5 to 6.5) | 0.02 |
| Clinical response | 28 (54.9%) | 16 (31.6%) | 2.63 (1.14 to 6.08) | 0.02 |
| Clinical remission | 25 (48.5%) | 14 (26.1%) | 2.67 (1.13 to 6.34) | 0.03 |
| MADRS-s | | | | |
| Decrease in score | 9.1 ± 8.87 | 5.6 ± 9.46 | 3.5 (0.3 to 6.8) | 0.04 |
| Clinical response | 16 (41.2%) | 7 (14.7%) | 4.06 (1.45 to 11.42) | 0.006 |
| Clinical remission | 17 (43.2%) | 10 (15.3%) | 4.22 (1.44 to 12.39) | 0.008 |
| EQ-5D-3L | 0.07 ± 0.17 | 0.05 ± 0.15 | 0.04 (0.00 to 0.09) | 0.08 |

HDRS, Hamilton Depression Rating Scale[3]; MADRS, Montgomery-Åsberg Depression Rating Scale[13]; MADRS-s, Montgomery-Åsberg Depression Rating Scale-self report[14]. EQ-5D-3L, quality of life measure[8–10]; CI, confidence interval. Mean values are presented with '±' standard deviation values. HDRS, MADRS, MADRS-s change ratings are the change in total ratings from baseline to week 10. Between-group differences are shown for the changes in scores from baseline to week 10, and odds ratios are shown for the outcomes for clinical response and remission. Percentages for clinical response and remission outcomes are estimated based on odds ratios. HDRS scores range from 0 to 52 (minimal clinically significant difference = 3 points), MADRS scores range from 0 to 60; MADRS-s scores range from 0 to 54, with higher scores indicating more depression. Clinical response was defined as a decrease in the score (indicating less depressive severity) of 50% or more from baseline to week 10. Clinical remission was defined as: HDRS score of 7 or less; MADRS score of 10 or less; MADRS-s score of 12 or less.

### Table 30. Changes in depressive severity as measured by HDRS, MADRS and MADRS-s and quality of life as measured by EQ-5D-3L for participants at the US site at 10 weeks

| Outcome | Active (n = 29) | Sham (n = 29) | Difference or Odds Ratio (95% CI) | P value |
|---|---|---|---|---|
| **Primary Outcome** | | | | |
| Decrease in HDRS score | 11.7 ± 7.54 | 9.5 ± 8.54 | 2.2 (2.0 to 6.5) | 0.30 |
| **Secondary Outcomes** | | | | |
| HDRS | | | | |
| Clinical response | 17 (67.5%) | 10 (44.8%) | 2.57 (0.73 to 9.04) | 0.13 |
| Clinical remission | 16 (67.7%) | 10 (43.6%) | 2.72 (0.75 to 9.86) | 0.11 |
| MADRS | | | | |
| Decrease in score | 11.6 ± 9.27 | 8.2 ± 9.72 | 3.4 (-1.5 to 8.2) | 0.17 |
| Clinical response | 18 (77.3%) | 9 (37.5%) | 5.79 (1.26 to 26.62) | 0.01 |
| Clinical remission | 17 (76.0%) | 11 (41.4%) | 4.58 (0.88 to 23.80) | 0.05 |
| MADRS-s | | | | |
| Decrease in score | 11.9 ± 11.18 | 8.8 ± 9.92 | 3.1 (-2.9 to 9.1) | 0.31 |
| Clinical response | 14 (65.7%) | 7 (43.2%) | 2.54 (0.70 to 9.27) | 0.16 |
| Clinical remission | 15 (73.0%) | 8 (42.5%) | 3.69 (0.84 to 16.19) | 0.07 |
| EQ-5D-3L | 0.07 ± 0.12 | 0.11 ± 0.19 | -0.03 (-0.10 to 0.04) | 0.38 |

HDRS, Hamilton Depression Rating Scale[3]; MADRS, Montgomery-Åsberg Depression Rating Scale[13]; MADRS-s, Montgomery-Åsberg Depression Rating Scale-self report[14]. EQ-5D-3L, quality of life measure[8–10]; CI, confidence interval. Mean values are presented with '±' standard deviation values. HDRS, MADRS, MADRS-s change ratings are the change in total ratings from baseline to week 10. Between-group differences are shown for the changes in scores from baseline to week 10, and odds ratios are shown for the outcomes for clinical response and remission. Percentages for clinical response and remission outcomes are estimated based on odds ratios. HDRS scores range from 0 to 52 (minimal clinically significant difference = 3 points), MADRS scores range from 0 to 60; MADRS-s scores range from 0 to 54, with higher scores indicating more depression. Clinical response was defined as a decrease in the score (indicating less depressive severity) of 50% or more from baseline to week 10. Clinical remission was defined as: HDRS score of 7 or less; MADRS score of 10 or less; MADRS-s score of 12 or less.

### Table 31. Changes in depressive severity as measured by HDRS, MADRS and MADRS-s and quality of life as measured by EQ-5D-3L for participants who correctly guessed their treatment group

| Outcome | Active (n = 66) | Sham (n = 33) | Difference or Odds Ratio (95% CI) | P value |
|---|---|---|---|---|
| **Primary Outcome** | | | | |
| Decrease in HDRS score | 10.0 ± 6.15 | 5.9 ± 6.05 | 4.1 (1.9 to 6.4) | <0.001 |
| **Secondary Outcomes** | | | | |
| HDRS | | | | |
| Clinical response | 35 (60.6%) | 5 (12.6%) | 10.77 (3.25 to 35.64) | <0.001 |
| Clinical remission | 29 (49.5%) | 5 (13.2%) | 6.45 (2.06 to 20.18) | <0.001 |
| MADRS | | | | |
| Decrease in score | 12.4 ± 9.59 | 6.5 ± 9.24 | 5.9 (2.4 to 9.5) | 0.001 |
| Clinical response | 40 (69.7%) | 7 (18.6%) | 10.08 (3.35 to 30.28) | <0.001 |
| Clinical remission | 36 (62.2%) | 8 (19.9%) | 6.63 (2.23 to 19.76) | <0.001 |
| MADRS-s | | | | |
| Decrease in score | 11.2 ± 9.03 | 3.1 ± 9.02 | 8.1 (4.5 to 11.7) | <0.001 |
| Clinical response | 25 (50.4%) | 5 (17.8%) | 4.71 (1.51 to 14.66) | 0.002 |
| Clinical remission | 28 (56.6%) | 5 (15.1%) | 7.35 (2.15 to 25.09) | <0.001 |
| EQ-5D-3L | 0.08 ± 0.16 | 0.04 ± 0.19 | 0.05 (-0.01 to 0.11) | 0.08 |

HDRS, Hamilton Depression Rating Scale[3]; MADRS, Montgomery-Åsberg Depression Rating Scale[13]; MADRS-s, Montgomery-Åsberg Depression Rating Scale-self report[14]. EQ-5D-3L, quality of life measure[8–10]; CI, confidence interval. Mean values are presented with '±' standard deviation values. HDRS, MADRS, MADRS-s change ratings are the change in total ratings from baseline to week 10. Between-group differences are shown for the changes in scores from baseline to week 10, and odds ratios are shown for the outcomes for clinical response and remission. Percentages for clinical response and remission outcomes are estimated based on odds ratios. HDRS scores range from 0 to 52 (minimal clinically significant difference = 3 points), MADRS scores range from 0 to 60; MADRS-s scores range from 0 to 54, with higher scores indicating more depression. Clinical response was defined as a decrease in the score (indicating less depressive severity) of 50% or more from baseline to week 10. Clinical remission was defined as: HDRS score of 7 or less; MADRS score of 10 or less; MADRS-s score of 12 or less.

#### Table 32. Changes in depressive severity as measured by HDRS, MADRS and MADRS-s and quality of life as measured by EQ-5D-3L for participants who incorrectly guessed their treatment group

| Outcome | Active (n = 19) | Sham (n = 48) | Difference or Odds Ratio (95% CI) | P value |
|---|---|---|---|---|
| **Primary Outcome** | | | | |
| Decrease in HDRS score | 7.1 ± 6.99 | 9.4 ± 6.90 | 2.4 (-5.8 to 1.1) | 0.18 |
| **Secondary Outcomes** | | | | |
| HDRS | | | | |
| Clinical response | 6 (40.6%) | 16 (41.7%) | 0.94 (0.28 to 3.22) | 0.94 |
| Clinical remission | 5 (34.3%) | 12 (30.9%) | 1.17 (0.31 to 4.38) | 0.83 |
| MADRS | | | | |
| Decrease in score | 7.9 ± 8.81 | 9.2 ± 8.27 | 1.3 (-5.7 to 3.0) | 0.54 |
| Clinical response | 6 (34.1%) | 18 (37.6%) | 0.86 (0.22 to 3.26) | 0.84 |
| Clinical remission | 6 (37.3%) | 17 (35.5%) | 1.08 (0.28 to 4.09) | 0.91 |
| MADRS-s | | | | |
| Decrease in score | 8.9 ± 9.95 | 9.6 ± 9.42 | 0.7 (-6.7 to 5.3) | 0.81 |
| Clinical response | 5 (55.0%) | 9 (29.0%) | 3.00 (0.67 to 13.40) | 0.19 |
| Clinical remission | 4 (46.5%) | 13 (34.4%) | 1.66 (0.30 to 9.08) | 0.59 |
| EQ-5D-3L | 0.05 ± 0.14 | 0.09 ± 0.15 | -0.03 (-0.09 to 0.03) | 0.37 |

HDRS, Hamilton Depression Rating Scale[3]; MADRS, Montgomery-Åsberg Depression Rating Scale[13]; MADRS-s, Montgomery-Åsberg Depression Rating Scale-self report[14]. EQ-5D-3L, quality of life measure[8–10]; CI, confidence interval. Mean values are presented with '±' standard deviation values. HDRS, MADRS, MADRS-s change ratings are the change in total ratings from baseline to week 10. Between-group differences are shown for the changes in scores from baseline to week 10, and odds ratios are shown for the outcomes for clinical response and remission. Percentages for clinical response and remission outcomes are estimated based on odds ratios. HDRS scores range from 0 to 52 (minimal clinically significant difference = 3 points), MADRS scores range from 0 to 60; MADRS-s scores range from 0 to 54, with higher scores indicating more depression. Clinical response was defined as a decrease in the score (indicating less depressive severity) of 50% or more from baseline to week 10. Clinical remission was defined as: HDRS score of 7 or less; MADRS score of 10 or less; MADRS-s score of 12 or less.

### Site heterogeneity

The pre-specified site heterogeneity analysis was performed. The P value for the group x site interaction was p = 0.09. The stratified site analysis shows an estimated change from baseline to Week 10 in HDRS-17 is -2.2 for the US and -2.1 for the UK. Therefore, while statistically observed, the finding is not understood to have clinical interpretation implications, and the sites declared pooled as reported in the manuscript.

### Open label phase analysis

#### Table S33. Demographic and clinical characteristics of week 20 completer patients at baseline

| Characteristic | Active (N=67) | Sham (N=71) |
|---|---|---|
| Age | 37.5 + 10.6 | 37.6 + 10.4 |
| Gender | | |
| Female | 42 (63) | 54 (76) |
| Race | | |
| Asian | 5 (7) | 2 (3) |
| Black or African American | 3 (4) | 1 (1) |
| Native Hawaiian or Other | 0 (0) | 0 (0) |
| White | 56 (84) | 60 (85) |
| Other | 3 (4) | 8 (11) |
| Missing | 0 (0) | 0 (0) |
| Ethnicity | | |
| Hispanic or Latino | 6 (9) | 8 (11) |
| Not Hispanic or Latino | 59 (88) | 59 (83) |
| Not reported | 2 (3) | 3 (4) |
| Missing | 0 (0) | 1 (1) |
| Educational Level | | |
| Less than High School/Secondary School | 1 (1) | 0 (0) |
| Some College | 15 (22) | 15 (21) |
| Diploma | 5 (7) | 6 (8) |
| Bachelor's or Professional Degree | 28 (42) | 32 (45) |
| Master's or Doctoral Degree | 18 (27) | 17 (24) |
| Prefer not to answer/Missing | 0 (0) | 1 (1) |
| Clinical ratings | | |
| HDRS | 19.0 + 2.7 | 18.8 + 2.6 |
| MADRS | 24.5 + 4.3 | 23.9 + 5.2 |
| MADRS-s | 26.2 + 6.6 | 25.6 + 6.3 |
| HAMA | 15.2 + 4.4 | 14.4 + 4.7 |
| YMRS | 2.1 + 1.7 | 2.1 + 1.6 |
| RAVLT | 57.5 + 11.3 | 57.0 + 12.6 |
| SDMT | 51.4 + 9.9 | 51.0 + 10.3 |

Categorical variables are presented as number of participants with percentage in parentheses for gender, race, ethnicity and educational level. Mean values are presented with '+' standard deviation values. HDRS, Hamilton Depression Rating Scale; MADRS, Montgomery-Åsberg Depression Rating Scale; MADRS-s, Montgomery–Åsberg Depression Rating Scale-self report; HAMA, Hamilton Anxiety Rating Scale; YMRS, Young Mania Rating Scale; RAVLT, Rey Auditory Verbal Learning Test; SDMT, Symbol Digit Modalities Test. SDMT active, n=65, SDMT sham, n=65. HDRS scores range from 0 to 52, MADRS scores range from 0 to 60, MADRS-s scores range from 0 to 54, with higher scores indicating more depression. RAVLT scores range from 0 to 75. SDMT scores range from 0 to 110.

### Transition analysis

A post hoc transition analysis was completed for all participants who completed the blinded phase of the trial and continued in the open label phase of the trial.

Active tDCS treatment arm
- > 95% of participants maintained clinical response from Week 10 to Week 20
- ~40% of participants who did not show a clinical response at Week 10 showed a clinical response at Week 20

Sham tDCS treatment arm
- ~90% of participants maintained clinical response from Week 10 to Week 20
- 50% of participants who did not show a clinical response at Week 10 showed a clinical response at Week 20
-

The sample size is not large enough to parse out the impact of the amount of simulation time.

#### Table S34. Transition analysis of week 10 and week 20 response rates for patients who completed >=660 minutes of stimulation in the open label phase

| Active Group | | | | Sham Group | | | |
|---|---|---|---|---|---|---|---|
| Week_10 | Week_20 | | | Week_10 | Week_20 | | |
| N<br>Row %<br>Column % | Clinical Response | No clinical response | Total | N<br>Row %<br>Column % | Clinical Response | No clinical response | Total |
| Clinical Response | 17<br>100%<br>85% | 0<br>0%<br>0% | 17 | Clinical Response | 8<br>88.89%<br>28.57% | 1<br>11.11%<br>4.76% | 9 |
| No clinical response | 3<br>37.5%<br>15% | 5<br>62.5%<br>100% | 8 | No clinical response | 20<br>50.00%<br>71.43% | 20<br>50.00<br>95.24% | 40 |
| Total | 20<br>80% | 5<br>20% | 25<br>100% | Total | 28<br>57.14% | 21<br>42.86% | 49<br>100% |
| Frequency Missing = 4 | | | | Frequency Missing = 4 | | | |

Clinical response, >=50% reduction from baseline in Hamilton depression rating scale score.

Table S34 shows response rates at week 10 and week 20 for all participants in the active and sham groups who completed a minimum 22 sessions in the blinded phase of the trial and a minimum of 22 sessions (660 minutes of active stimulation) in the open label phase of the trial.

In the active group, all participants who responded to treatment in the blinded phase, continued to be responders at week 20. Of the 8 participants who did not respond to treatment during the blinded phase, 3 participants were responders at week 20.

In the sham group, 8 of the 9 participants who responded to treatment in the blinded phase continued to be responders at week 20 after receiving the active treatment. Of the 40 participants who did not respond during the blinded phase, 20 participants were responders at week 20 after receiving the active treatment.

### Table S35. Transition analysis of week 10 and week 20 response rates for patients who completed >=330 minutes of stimulation in the open label phase

| Active Group | | | | Sham Group | | | |
|---|---|---|---|---|---|---|---|
| Week_10 | Week_20 | | | Week_10 | Week_20 | | |
| N<br>Row %<br>Column % | Clinical Response | No clinical response | Total | N<br>Row %<br>Column % | Clinical Response | No clinical response | Total |
| Clinical Response | 26<br>96.3%<br>78.79% | 1<br>3.7%<br>11.11% | 27 | Clinical Response | 9<br>81.82%<br>30% | 2<br>18.18%<br>9.09% | 11 |
| No clinical response | 7<br>46.67%<br>21.21% | 8<br>53.33%<br>88.89% | 15 | No clinical response | 21<br>51.22%<br>70% | 20<br>48.78%<br>90.91% | 41 |
| Total | 33<br>78.57% | 9<br>21.43% | 42<br>100% | Total | 30<br>57.69% | 22<br>42.31% | 52<br>100% |
| Frequency Missing = 9 | | | | Frequency Missing = 5 | | | |

Clinical response, >=50% reduction from baseline in Hamilton depression rating scale score.

Table S35 shows response rates at week 10 and week 20 for all participants in the active and sham groups who completed a minimum 22 sessions in the blinded phase of the trial and a minimum of 11 sessions (330 minutes of active stimulation) in the open label phase of the trial.

In the active group 26 of the 27 participants who responded to treatment in the blinded phase continued to be responders at week 20. Of the 15 participants who did not respond during the blinded phase, 7 participants were responders at week 20.

In the sham group 9 of the 11 participants who responded to treatment in the blinded phase continued to be responders at week 20. Of the 41 participants who did not respond during the blinded phase, 21 participants were responders at week 20.

### Table S36. Transition analysis of week 10 and week 20 response rates for patients who completed >=65 minutes of stimulation in the open label phase

| Active Group | | | | Sham Group | | | |
|---|---|---|---|---|---|---|---|
| Week_10 | Week_20 | | | Week_10 | Week_20 | | |
| N<br>Row %<br>Column % | Clinical Response | No clinical response | Total | N<br>Row %<br>Column % | Clinical Response | No clinical response | Total |
| **Clinical Response** | 28<br>96.55%<br>75.68% | 1<br>3.45%<br>8.33% | 29 | **Clinical Response** | 9<br>81.82%<br>30% | 2<br>18.18%<br>8.70% | 11 |
| **No clinical response** | 9<br>45%<br>24.32% | 11<br>55%<br>91.67% | 20 | **No clinical response** | 21<br>50%<br>70% | 21<br>50%<br>91.30% | 42 |
| **Total** | 37<br>75.51% | 12<br>24.49% | 49<br>100% | **Total** | 30<br>56.60% | 23<br>43.40% | 53<br>100% |
| **Frequency Missing = 10** | | | | **Frequency Missing = 10** | | | |

Clinical response, >=50% reduction from baseline in Hamilton depression rating scale score.

Table S36 shows response rates at week 10 and week 20 for all participants in the active and sham groups who completed a minimum 22 sessions in the blinded phase of the trial and a minimum of 165 minutes of active stimulation in the open label phase of the trial.

In the active group 28 of the 29 participants who responded to treatment in the blinded phase continued to be responders at week 20. Of the 20 participants who did not respond during the blinded phase, 9 participants were responders at week 20.

In the sham group 9 of the 11 participants who responded to treatment in the blinded phase continued to be responders at week 20. Of the 42 participants who did not respond during the blinded phase, 21 participants were responders at week 20.

### Table S37. Transition analysis of week 10 and week 20 response rates for patients who completed >=80 minutes of stimulation in the open label phase

| Active Group | | | | Sham Group | | | |
|---|---|---|---|---|---|---|---|
| Week_10 | Week_20 | | | Week_10 | Week_20 | | |
| N<br>Row %<br>Column % | Clinical Response | No clinical response | Total | N<br>Row %<br>Column % | Clinical Response | No clinical response | Total |
| Clinical Response | 29<br>96.67%<br>76.32% | 1<br>3.33%<br>8.33% | 30 | Clinical Response | 9<br>81.82%<br>30% | 2<br>18.18%<br>8.70% | 11 |
| No clinical response | 9<br>45%<br>23.68% | 11<br>55%<br>91.67% | 20 | No clinical response | 21<br>50%<br>70% | 21<br>50%<br>91.30% | 42 |
| Total | 38<br>76% | 12<br>24% | 50<br>100% | Total | 30<br>56% | 23<br>43.40% | 53<br>100% |
| Frequency Missing = 11 | | | | Frequency Missing = 13 | | | |

Clinical response, >=50% reduction from baseline in Hamilton depression rating scale score.

Table S37 shows response rates at week 10 and week 20 for all participants in the active and sham groups who completed a minimum 22 sessions in the blinded phase of the trial and a minimum of 80 minutes of active stimulation in the open label phase of the trial.

In the active group 29 of the 30 participants who responded to treatment in the blinded phase continued to be responders at week 20. Of the 20 participants who did not respond during the blinded phase, 9 participants were responders at week 20.

In the sham group 9 of the 11 participants who responded to treatment in the blinded phase continued to be responders at week 20. Of the 42 participants who did not respond during the blinded phase, 21 participants were responders at week 20.

### Table S38. Transition analysis of week 10 and week 20 response rates for patients who completed >=0 minutes of stimulation in the open label phase

| Active Group | | | | Sham Group | | | |
|---|---|---|---|---|---|---|---|
| **Week_10** | **Week_20** | | | **Week_10** | **Week_20** | | |
| **N<br>Row %<br>Column %** | **Clinical Response** | **No clinical response** | **Total** | **N<br>Row %<br>Column %** | **Clinical Response** | **No clinical response** | **Total** |
| **Clinical Response** | 30<br>96.77%<br>76.92% | 1<br>3.23%<br>6.25% | 31 | **Clinical Response** | 10<br>83.33%<br>31.25% | 2<br>16.67%<br>8.33% | 12 |
| **No clinical response** | 9<br>37.50%<br>23.08% | 15<br>62.50%<br>93.75% | 24 | **No clinical response** | 22<br>50%<br>68.75% | 22<br>50%<br>91.67% | 44 |
| **Total** | 39<br>70.91% | 16<br>29.09% | 55<br>100% | **Total** | 32<br>57.14% | 24<br>42.86% | 56<br>100% |
| **Frequency Missing = 32** | | | | **Frequency Missing = 30** | | | |

Clinical response, <=50% reduction from baseline in Hamilton depression rating scale score.

Table S38 shows response rates at week 10 and week 20 for all participants in the active and sham groups who completed a minimum 22 sessions in the blinded phase of the trial and a >=0 minutes of active stimulation in the open label phase of the trial.

In the active group 30 of the 31 participants who responded to treatment in the blinded phase continued to be responders at week 20. Of the 24 participants who did not respond during the blinded phase, 9 participants were responders at week 20.

In the sham group 10 of the 12 participants who responded to treatment in the blinded phase continued to be responders at week 20. Of the 44 participants who did not respond during the blinded phase, 22 participants were responders at week 20.

**Supplementary Appendix**

Home-based transcranial direct current stimulation RCT in major depression

### Table S39. All unanticipated device related adverse events in the open label phase

| | Active (N= 67) | | | Sham (N= 71) | | | Group Difference | | | |
|---|---|---|---|---|---|---|---|---|---|---|
| | Events | Subjs | % | Events | Subjs | % | Δ | LB | UB | p |
| **All Events** | **3** | **3** | **4.5%** | **6** | **6** | **8.5%** | **4.0%** | **-13.5%** | **5.2%** | **0.50** |
| **Ear and labyrinth disorders** | **1** | **1** | **1.5%** | **1** | **1** | **1.4%** | **0.1%** | **-6.3%** | **6.9%** | **0.99** |
| Tinnitus | 1 | 1 | 1.5% | 1 | 1 | 1.4% | 0.1% | -6.3% | 6.9% | 0.99 |
| **Gastrointestinal disorders** | **0** | **0** | **0.0%** | **1** | **1** | **1.4%** | **1.4%** | **-7.7%** | **4.4%** | **0.99** |
| Nausea | 0 | 0 | 0.0% | 1 | 1 | 1.4% | 1.4% | -7.7% | 4.4% | 0.99 |
| **Nervous system disorders** | **1** | **1** | **1.5%** | **0** | **0** | **0.0%** | **1.5%** | **-3.8%** | **8.2%** | **0.49** |
| Headache | 1 | 1 | 1.5% | 0 | 0 | 0.0% | 1.5% | -3.8% | 8.2% | 0.49 |
| **Psychiatric disorders** | **1** | **1** | **1.5%** | **0** | **0** | **0.0%** | **1.5%** | **-3.8%** | **8.2%** | **0.49** |
| Mood Altered | 1 | 1 | 1.5% | 0 | 0 | 0.0% | 1.5% | -3.8% | 8.2% | 0.49 |
| **Skin and subcutaneous tissue disorders** | **0** | **0** | **0.0%** | **4** | **4** | **5.6%** | **5.6%** | **-13.8%** | **0.2%** | **0.12** |
| Dry skin | 0 | 0 | 0.0% | 4 | 4 | 5.6% | 5.6% | -13.8% | 0.2% | 0.12 |

Adverse events are reported for participants that completed >0 minutes of stimulation during the open label phase. An adverse event was present if the participant rated that it was at least possibly associated with the intervention. Participants rated the severity of the adverse events as mild, moderate, or severe, which were assessed by the investigator. P values represent difference between groups. LB, lower bound; UB, upper bound.

**Supplementary Appendix**

Home-based transcranial direct current stimulation RCT in major depression

### Table S40. All unanticipated adverse events in the open label phase

| | Active (N= 67) | | | Sham (N= 71) | | | Group Difference | | | |
|---|---|---|---|---|---|---|---|---|---|---|
| | Events | Subjs | % | Events | Subjs | % | Δ | LB | UB | p |
| **All Events** | **6** | **6** | **9.0%** | **14** | **12** | **16.9%** | **7.9%** | **20.1%** | **3.7%** | **0.21** |
| **Blood and lymphatic system disorders** | **0** | **0** | **0.0%** | **1** | **1** | **1.4%** | **1.4%** | **-7.7%** | **4.4%** | **0.99** |
| Anaemia | 0 | 0 | 0.0% | 1 | 1 | 1.4% | 1.4% | -7.7% | 4.4% | 0.99 |
| **Ear and labyrinth disorders** | **1** | **1** | **1.5%** | **1** | **1** | **1.4%** | **0.1%** | **-6.3%** | **6.9%** | **0.99** |
| Tinnitus | 1 | 1 | 1.5% | 1 | 1 | 1.4% | 0.1% | -6.3% | 6.9% | 0.99 |
| **Gastrointestinal disorders** | **0** | **0** | **0.0%** | **1** | **1** | **1.4%** | **1.4%** | **-7.7%** | **4.4%** | **0.99** |
| Nausea | 0 | 0 | 0.0% | 1 | 1 | 1.4% | 1.4% | -7.7% | 4.4% | 0.99 |
| **General disorders and administration site conditions** | **0** | **0** | **0.0%** | **2** | **2** | **2.8%** | **2.8%** | **10.0%** | **3.1%** | **0.50** |
| Asthenia | 0 | 0 | 0.0% | 1 | 1 | 1.4% | 1.4% | -7.7% | 4.4% | 0.99 |
| Illness | 0 | 0 | 0.0% | 1 | 1 | 1.4% | 1.4% | -7.7% | 4.4% | 0.99 |
| **Immune system disorders** | **1** | **1** | **1.5%** | **0** | **0** | **0.0%** | **1.5%** | **-3.8%** | **8.2%** | **0.49** |
| Seasonal allergy | 1 | 1 | 1.5% | 0 | 0 | 0.0% | 1.5% | -3.8% | 8.2% | 0.49 |
| **Infections and infestations** | **1** | **1** | **1.5%** | **4** | **4** | **5.6%** | **4.1%** | **12.5%** | **3.1%** | **0.37** |
| COVID-19 | 0 | 0 | 0.0% | 2 | 2 | 2.8% | 2.8% | 10.0% | 3.1% | 0.50 |
| Lower respiratory tract infection | 0 | 0 | 0.0% | 2 | 2 | 2.8% | 2.8% | 10.0% | 3.1% | 0.50 |
| Sinusitis | 1 | 1 | 1.5% | 0 | 0 | 0.0% | 1.5% | -3.8% | 8.2% | 0.49 |
| **Musculoskeletal and connective tissue disorders** | **1** | **1** | **1.5%** | **0** | **0** | **0.0%** | **1.5%** | **-3.8%** | **8.2%** | **0.49** |
| Pain in extremity | 1 | 1 | 1.5% | 0 | 0 | 0.0% | 1.5% | -3.8% | 8.2% | 0.49 |
| **Nervous system disorders** | **1** | **1** | **1.5%** | **0** | **0** | **0.0%** | **1.5%** | **-3.8%** | **8.2%** | **0.49** |
| Headache | 1 | 1 | 1.5% | 0 | 0 | 0.0% | 1.5% | -3.8% | 8.2% | 0.49 |
| **Psychiatric disorders** | **1** | **1** | **1.5%** | **0** | **0** | **0.0%** | **1.5%** | **-3.8%** | **8.2%** | **0.49** |
| Mood Altered | 1 | 1 | 1.5% | 0 | 0 | 0.0% | 1.5% | -3.8% | 8.2% | 0.49 |
| **Reproductive system and breast disorders** | **0** | **0** | **0.0%** | **1** | **1** | **1.4%** | **1.4%** | **-7.7%** | **4.4%** | **0.99** |
| Pelvic pain | 0 | 0 | 0.0% | 1 | 1 | 1.4% | 1.4% | -7.7% | 4.4% | 0.99 |
| **Skin and subcutaneous tissue disorders** | **0** | **0** | **0.0%** | **4** | **4** | **5.6%** | **5.6%** | **13.8%** | **0.2%** | **0.12** |
| Dry skin | 0 | 0 | 0.0% | 4 | 4 | 5.6% | 5.6% | 13.8% | 0.2% | 0.12 |

All unanticipated adverse events that were reported between week 10 and week 20 including those that weren't associated with the intervention for participants that completed >0 minutes of stimulation during the open label phase. Participants rated the severity of the adverse events as mild, moderate, or severe, which were assessed by the investigator. P values represent difference between groups. LB, lower bound; UB, upper bound.

### Table S41. All anticipated adverse events in the open label phase as measured by the tDCS Adverse Events Questionnaire.[19]

| | Active (N=56) | | | | Sham (N=57) | | | | |
|---|---|---|---|---|---|---|---|---|---|
| **Adverse event category** | **Total** | **Mild** | **Moderate** | **Severe** | **Total** | **Mild** | **Moderate** | **Severe** | **P Value** |
| Headache | 12 (21) | 8 (14) | 3 (5) | 1 (2) | 10 (18) | 7 (12) | 3 (5) | 0 (0) | 0.64 |
| Neck Pain | 1 (2) | 1 (2) | 0 (0) | 0 (0) | 2 (4) | 0 (0) | 2 (4) | 0 (0) | 1.00 |
| Scalp pain | 10 (18) | 8 (14) | 2 (4) | 0 (0) | 14 (25) | 10 (18) | 4 (7) | 0 (0) | 0.49 |
| Itching | 26 (46) | 19 (34) | 6 (11) | 1 (2) | 23 (40) | 19 (33) | 3 (5) | 1 (2) | 0.57 |
| Burning sensation | 22 (39) | 20 (36) | 2 (4) | 0 (0) | 26 (46) | 20 (35) | 6 (11) | 0 (0) | 0.57 |
| Skin redness | 34 (61) | 28 (50) | 6 (11) | 0 (0) | 30 (53) | 30 (53) | 0 (0) | 0 (0) | 0.45 |
| Sleepiness | 6 (11) | 4 (7) | 2 (4) | 0 (0) | 9 (16) | 8 (14) | 1 (2) | 0 (0) | 0.58 |
| Trouble concentrating | 5 (9) | 2 (4) | 3 (5) | 0 (0) | 2 (4) | 2 (4) | 0 (0) | 0 (0) | 0.27 |
| Acute mood change | 1 (2) | 1 (2) | 0 (0) | 0 (0) | 0 (0) | 0 (0) | 0 (0) | 0 (0) | 0.50 |

Values are number of participants with percentage in parentheses. An adverse event was present if the participant rated that it was at least remotely possible that it was associated with the intervention. Participants rated the severity of the adverse events as mild, moderate, or severe. P values represent group differences of the total number of events per event category.

### Sample representativeness

#### Table S42. Background information regarding socio-demographic characteristics of patients with Major Depressive Disorder and the representativeness of participants

| **Category** | |
|---|---|
| Disease, problem, or condition under investigation | Major Depressive Disorder (MDD) |
| Special considerations related to | |
| Sex and gender | MDD affects women more than men (ratio of 3:2)[20] |
| Age | Prevalence of depression rates vary by age and peak in older adulthood (55-74 years).[20] |
| Race or ethnic group | MDD affects White and Hispanic racial groups the most in the United States, with lower levels of depression in the African American population.[21] Multiracial adults have higher rates of depression,[21] but make up a smaller number of the population.[22] In the United Kingdom the majority of MDD patients are White due to a largely White population.[23] Black, Mixed and Asian populations experience higher rates of depression (24-29%) compared to White populations (16-21%).[24] |
| Other considerations | Anxiety disorders are common comorbid disorders in MDD[25, 26] and therefore were only included as an exclusion criterion if they were considered to be the primary disorder. |
| Overall representativeness of this trial | The participants in the present trial demonstrated the expected ratio of women to men, with larger proportion of female participants (69%) relative to male participants (31%). Options for gender were male and female. Participants were asked to report their race and their ethnicity. The response options for race were Asian, Black or African American, Native Hawaiian or Other, White and Other. The response options for ethnicity were Hispanic or Latino, Not Hispanic or Latino or Not Reported. White participants make up the majority of the study sample (83%), which reflects the general demographic population of the United Kingdom[23] and United States[22]. Hispanic and Latino participants made up 7% of the study sample and 31% of the sample from the United Stated. People with a Hispanic and Latino ethnicity represent 19% of the United States population.[22] Hispanic or Latino is not a common ethnicity un the United Kingdom and represented 3% of the United Kingdom sample. Black and African American populations were underrepresented in our sample. 2% of the study sample were Black or African American. This is lower than the number of Black or African American people both in the United States (14%)[22] and the United Kingdom (4%)[23], where rates of depression are higher among Black populations.[24] |
